# A cross-disorder dosage sensitivity map of the human genome

**DOI:** 10.1101/2021.01.26.21250098

**Authors:** Ryan L. Collins, Joseph T. Glessner, Eleonora Porcu, Lisa-Marie Niestroj, Jacob Ulirsch, Georgios Kellaris, Daniel P. Howrigan, Selin Everett, Kiana Mohajeri, Xander Nuttle, Chelsea Lowther, Jack Fu, Philip M. Boone, Farid Ullah, Kaitlin E. Samocha, Konrad Karczewski, Diane Lucente, Epi25 Consortium, James F. Gusella, Hilary Finucane, Ludmilla Matyakhina, Swaroop Aradhya, Jeanne Meck, Dennis Lal, Benjamin M. Neale, Jennelle C. Hodge, Alexandre Reymond, Zoltan Kutalik, Nicholas Katsanis, Erica E. Davis, Hakon Hakonarson, Shamil Sunyaev, Harrison Brand, Michael E. Talkowski

## Abstract

Rare deletions and duplications of genomic segments, collectively known as rare copy number variants (rCNVs), contribute to a broad spectrum of human diseases. To date, most disease-association studies of rCNVs have focused on recognized genomic disorders or on the impact of haploinsufficiency caused by deletions. By comparison, our understanding of duplications in disease remains rudimentary as very few individual genes are known to be triplosensitive (*i*.*e*., duplication intolerant). In this study, we meta-analyzed rCNVs from 753,994 individuals across 30 primarily neurological disease phenotypes to create a genome-wide catalog of rCNV association statistics across disorders. We discovered 114 rCNV-disease associations at 52 distinct loci surpassing genome-wide significance (P=3.72×10^−6^), 42% of which involve duplications. Using Bayesian fine-mapping methods, we further prioritized 38 novel triplosensitive disease genes (*e*.*g*., *GMEB2* in brain abnormalities), including three known haploinsufficient genes that we now reveal as bidirectionally dosage sensitive (*e*.*g*., *ANKRD11* in growth abnormalities). By integrating our results with prior literature, we found that disease-associated rCNV segments were enriched for genes constrained against damaging coding variation and identified likely dominant driver genes for about one-third (32%) of rCNV segments based on *de novo* mutations from exome sequencing studies of developmental disorders. However, while the presence of constrained driver genes was a common feature of many pathogenic large rCNVs across disorders, most of the rCNVs showing genome-wide significant association were incompletely penetrant (mean odds ratio=11.6) and we also identified two examples of noncoding disease-associated rCNVs (*e*.*g*., intronic *CADM2* deletions in behavioral disorders). Finally, we developed a statistical model to predict dosage sensitivity for all genes, which defined 3,006 haploinsufficient and 295 triplosensitive genes where the effect sizes of rCNVs were comparable to deletions of genes constrained against truncating mutations. These dosage sensitivity scores classified disease genes across molecular mechanisms, prioritized pathogenic *de novo* rCNVs in children with autism, and revealed features that distinguished haploinsufficient and triplosensitive genes, such as insulation from other genes and local *cis*-regulatory complexity. Collectively, the cross-disorder rCNV maps and metrics derived in this study provide the most comprehensive assessment of dosage sensitive genomic segments and genes in disease to date and set the foundation for future studies of dosage sensitivity throughout the human genome.

## Introduction

Natural selection maintains nearly all mammalian genomes as diploid (*i*.*e*., two copies of each chromosome).^1^ While deletions and duplications of genomic segments, collectively known as copy-number variants (CNVs), have been recognized as important mechanisms of evolutionary adaptation for over 50 years,^2^ there are relatively few examples of CNVs that provide adaptive advantages in humans.^3^ Instead, most large CNVs (≥100 kilobases) typically experience strong purifying selection and are held at low frequencies in the global population.^4^ These rare (frequency <1%) CNVs (rCNVs) have been widely associated with Mendelian and complex diseases.^5^ Although deletions are usually more damaging than duplications,^6-8^ both have been causally implicated in disease, such as duplications of *APP* in early-onset Alzheimer’s disease and deletions of *NRXN1* in a range of neuropsychiatric disorders.^9-11^ While the effects of deletions are usually assumed to be mediated by the loss of one or more genes or functional elements, the potential molecular consequences of duplications are more variable and context-dependent.^12^ Clearly, the impact of CNVs on human traits and diseases is widespread and complex.

A subset of disease-associated rCNVs, known as “genomic disorders” (GDs), have been prominent in the Mendelian and complex disease literature for decades.^13-15^ GDs are sites of recurrent rCNVs, often formed by non-allelic homologous recombination (NAHR) between tracts of nearly identical sequence flanking specific regions of the genome.^16^ Several dozen GDs have been reported to date, including “reciprocal” GDs, such as 16p11.2 and 22q11.2, where deletions and duplications of the same locus have been independently associated with disease.^17^ Many GDs manifest with variable phenotypes, but collectively comprise one of the most common genetic causes of abnormal neurodevelopment.^6^ Recent population-scale biobank studies have shown that GD-associated rCNVs also have subtle effects on traits in the general population, like height and blood pressure, even in the absence of disease.^18-20^ Some reciprocal GDs have been linked to “mirror” phenotypes, wherein decreased DNA dosage leads to one phenotype (*e*.*g*., obesity and macrocephaly in 16p11.2 deletions) while increased dosage leads to the opposite (*e*.*g*., underweight and microcephaly in 16p11.2 duplications).^21,22^ The existence of mirror phenotypes for reciprocal GDs suggests that one or more genes or elements within these large rCNVs may be dosage sensitive “drivers” of some aspects of their associated phenotypes.^23^ Indeed, sensitivity to decreased DNA dosage (*i*.*e*., haploinsufficiency) or increased DNA dosage (*i*.*e*., triplosensitivity) has already been clinically documented for individual genes,^24,25^ although the genome-wide patterns and properties of dosage sensitivity are largely opaque.

Despite the morbidity attributable to large rCNVs, our understanding of their pathogenic mechanisms remains limited for several reasons. Many disease-associated rCNVs exhibit incomplete penetrance and variable expressivity of complex syndromic phenotypes.^6,26^ Most large rCNVs exist at vanishingly low frequencies in the population, often being ascertained in a single individual.^27-29^ Many large rCNVs encompass dozens of genes, confounding the identification of the critical driver(s) underlying disease, while other rCNVs are restricted entirely to noncoding sequence. Furthermore, large rCNVs can have myriad indirect consequences, including regulatory, polygenic, or epistatic effects.^30^ Finally, CNVs have a lower (≥100-fold) mutational density than short variants (<50bp) in the human genome,^31^ which means that genome-wide studies of large rCNVs have required comparatively greater sample sizes to attain the statistical power necessary to detect disease associations.^32-35^ As a consequence, the existing lists of dosage-sensitive genomic segments and genes that exceed genome-wide significance thresholds or meet robust guidelines for clinical interpretation are limited: for example, the ClinGen Genome Dosage Map includes just 15 triplosensitive genes.^25^ While haploinsufficient genes can be revealed by analyses of either protein-truncating short variants or deletions,^36^ triplosensitive genes are only weakly predicted by both truncating and missense variants and thus require dedicated analyses of duplications for their confident identification.^27^ Our ability to interpret rCNVs—especially duplications—outside of established GDs therefore lags behind analyses of single nucleotide variants and large cohorts will be required to build comprehensive maps of dosage sensitivity across disorders.

In this study, we evaluated the impact of rCNVs in 30 human disease phenotypes (**Figure 1**). We harmonized large rCNV data from 753,994 individuals, including 293,235 cases and 460,759 controls, and conducted meta-analyses to create a genome-wide catalog of rCNV association statistics across disorders, which included over four dozen loci at strict genome-wide significance. Finally, we integrated these rCNVs with a compendium of genome annotations to computationally predict haploinsufficiency and triplosensitivity for all protein-coding genes, allowing us to define >3,000 high-confidence dosage sensitive genes and to investigate the general properties of dosage sensitivity throughout the genome. We provide all maps and metrics derived in this study as an open resource for the community and anticipate that the insights revealed here will have broad utility for population genomics and medical genetics.

**Figure 1.**
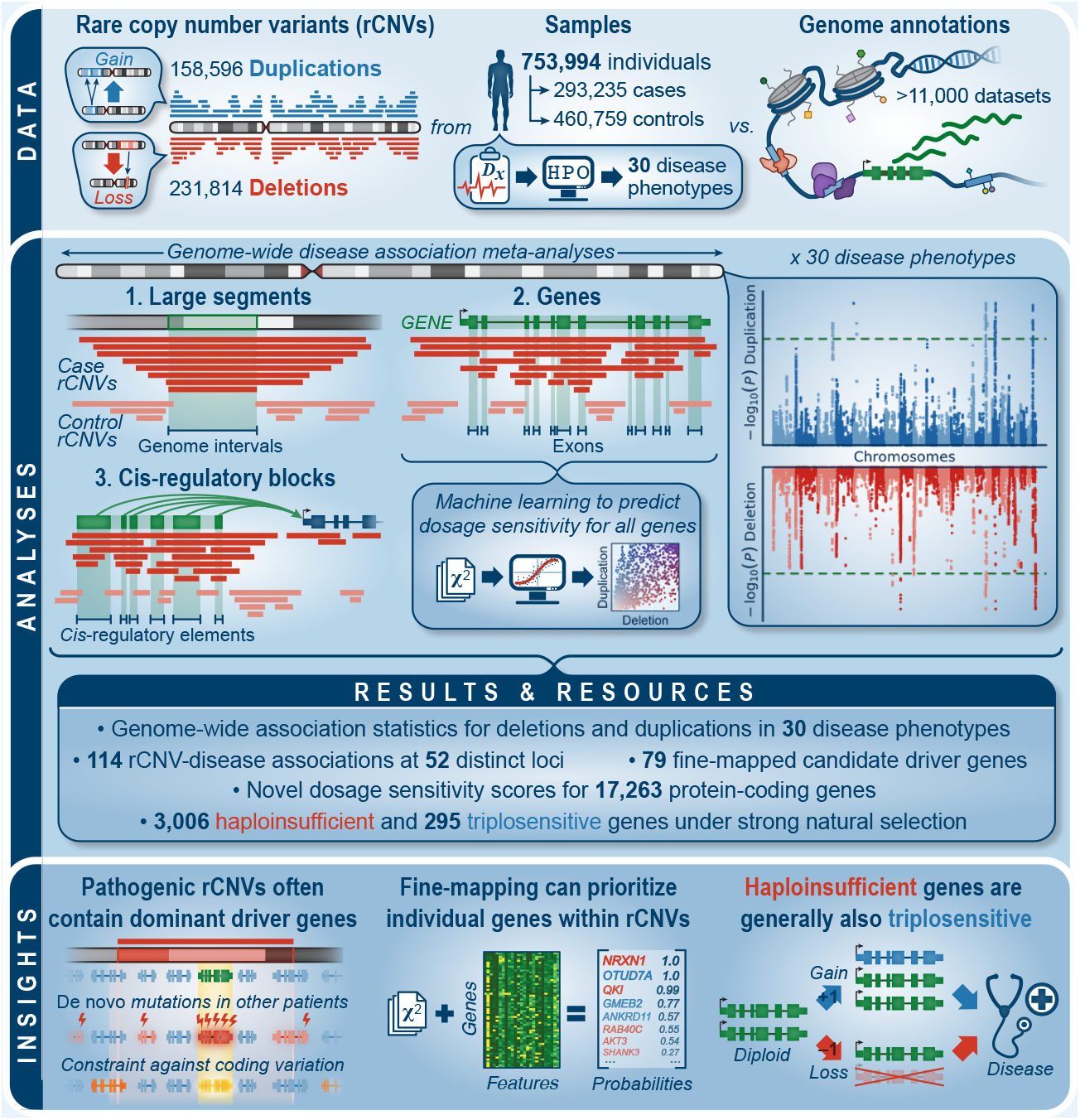
Study overview. Summary of data, analyses, results, and insights from this study.

## RESULTS

### Creating a cross-disorder catalog of rCNV associations

We aggregated rCNVs ascertained with microarrays from 13 sources, ranging from diagnostic laboratories to large-scale population genetic studies and national biobanks (**Table S1**). To account for heterogeneity in study designs and technical details across sources, we developed a harmonization procedure that retained large (≥100kb), focal (≤20Mb) CNVs observed in mostly non-repetitive genomic regions and at <1% frequency across every source in our dataset and in every global population documented by three genome sequencing-derived reference maps of CNVs.^27,37,38^ This procedure reduced variability in average CNV sizes, frequencies, and carrier rates by nearly two orders of magnitude across sources (**Figure S1**), with the final harmonized dataset including a total of 390,410 rCNVs in 753,994 individuals, or an average of one large rCNV observed in every 1.9 genomes. Finally, to control for residual heterogeneity, we further grouped sources into four independent cohorts based on their technical similarities, such as microarray platform and sample recruitment strategy.

The extent of phenotypic data varied between sources, ranging from the presence or absence of a single primary phenotype ascertained for disease association studies to deep phenome-wide metadata collected as part of population-scale biobanks. Therefore, we consolidated these disparate data into a smaller set of standardized phenotypes represented across cohorts in our analyses. To accomplish this, we first mapped all available phenotype data per sample onto the structured Human Phenotype Ontology (HPO) with a keyword-matching approach and performed recursive hierarchical clustering to define a minimal set of non-redundant primary phenotypes represented across cohorts.^39^ We required each phenotype to include a minimum of >500 samples in at least two independent cohorts, >2,000 samples in total across all cohorts, and to have less than 50% sample overlap with any other final phenotype. This process yielded a total of 30 disease phenotypes, including 16 neurological, 12 non-neurological, and two general catch-all phenotypes (**Figure S2**; **Table S2**). While imperfect, this principled approach partitioned our dataset into 293,235 samples matching one or more primary disease phenotype (*i*.*e*., “cases”) and 460,759 samples not matching any of the 30 disease phenotypes (*i*.*e*., “controls”). In this curated dataset, the phenotype terms encompassing the most samples typically represented high-level organ system disorders (*e*.*g*., nervous system abnormalities, N=161,891 cases) whereas terms with fewer samples represented more specific phenotypes (*e*.*g*., morphological brain abnormalities, N=2,634 cases).

Building on decades of seminal studies of CNV in disease,^18,32,35,40-43^ we leveraged the comparatively larger sample size of our aggregated dataset to identify loci where rCNVs were enriched in cases over controls. However, our rCNVs were large (interquartile range=132-324kb) and frequently overlapped multiple genes, which confounded most conventional genome-wide association methods. Therefore, we implemented three complementary approaches to detect rCNV-disease associations (**Figure S3**). We first searched for disease-associated large rCNV segments by dividing all 22 autosomes into 200kb sliding windows in 10kb steps. For each window, we performed an association meta-analysis of rCNVs per phenotype while controlling our false discovery rate (FDR) at genome-wide significance (P=3.72×10^−6^; **Note S1**) and further requiring nominal evidence (P<0.05) in at least two independent cohorts. The resulting test statistics appeared generally well calibrated across disorders (median genomic inflation, =0.99; **Figure S4**) and have been provided as a reference catalog for future studies (**File S1**). We next refined each significant association to the minimal region expected to contain the causal element(s) by implementing a Bayesian algorithm to define the 99% credible interval(s) per associated locus.^44^ In total, this approach discovered 102 rCNV-phenotype associations corresponding to 49 distinct large rCNV segments after collapsing across phenotypes (median size=450kb; 28 deletions & 21 duplications; **Figure 2**; **Tables S3-4**). We cross-examined these 49 significant rCNV segments with two orthogonal datasets to assess whether we had captured *bona fide* disease associations, finding an 8.2-fold overrepresentation among a curated list of 114 GDs from six literature-based surveys (**Figure S5A**; P<10^−5^, one-tailed 100,000-fold permutation test controlled for segment size) and a 1.8-fold enrichment for phenotype-matched disease genes (**Figure S5B**; **Table S5**; P<10^−5^, one-tailed permutation test controlled for number of genes per segment).^6,19,45-49^ Conversely, 18 of our 49 genome-wide significant rCNV segments had not been statistically associated with disease in prior studies and may therefore represent new discoveries (**Figure S6**).

**Figure 2.**
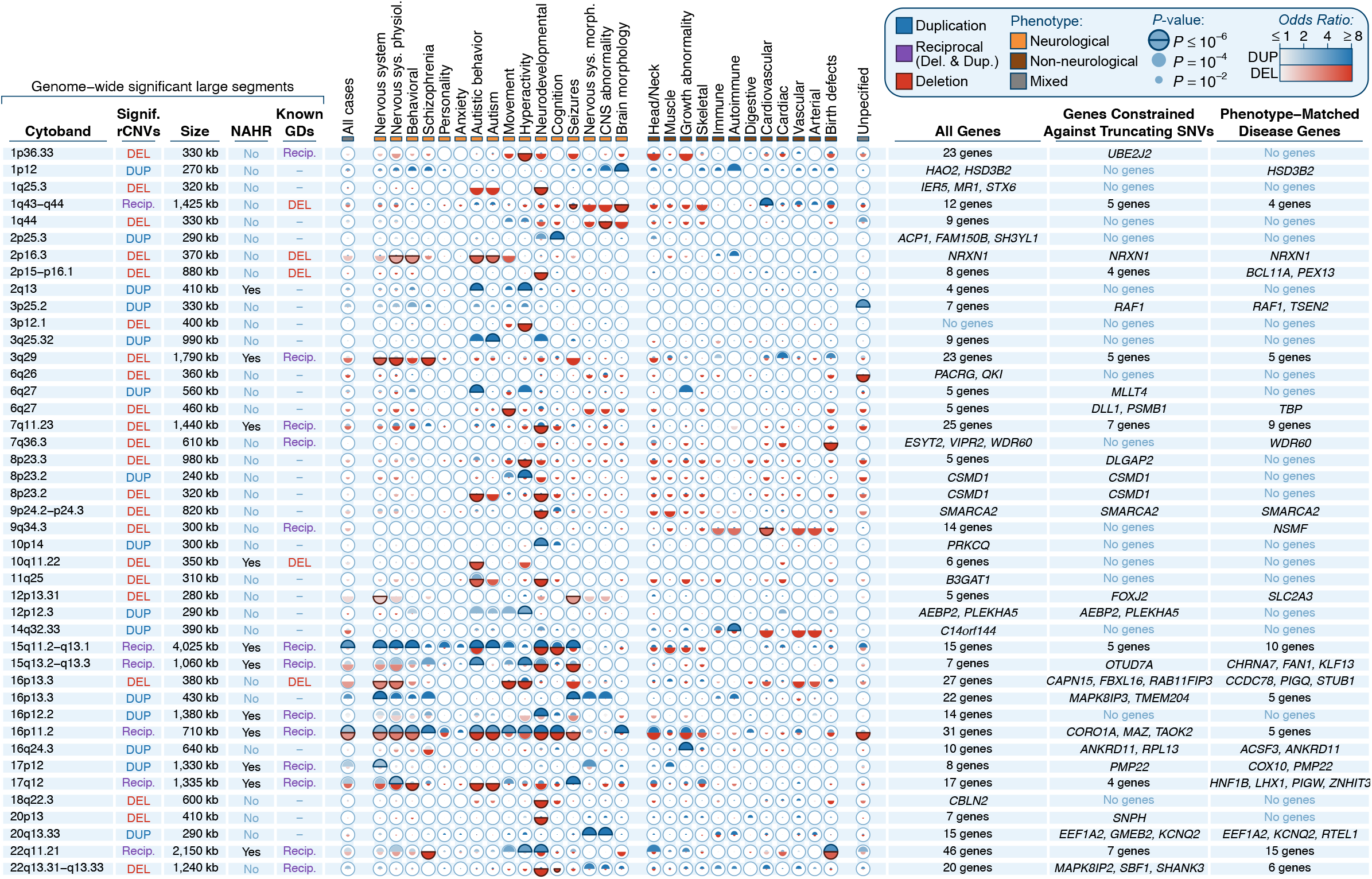
Disease-associated large rCNV segments at genome-wide significance. We identified 102 phenotype-rCNV associations at genome-wide significance (P≤3.72×10^−6^), which localized to 49 unique rCNV segments after collapsing across phenotypes. Details for each of the 49 segments are summarized here and are provided in **Tables S3-4**. Overlapping segments have been merged into single rows for clarity. For each locus, we provide segment size, predicted NAHR-mediated mechanism, overlap with genomic disorders (GDs) reported by at least one of six sources,^6,19,45-48^ meta-analysis association statistics for 30 phenotypes, and genic content, further partitioned by constraint against truncating point mutations and previously reported associations with the same disease.^49,50^ Association statistics are represented as one semicircle each for duplication (blue) and deletion (red) shaded by the effect size estimate with radii scaled proportional to the −log_10_ P-value. Sample sizes vary per phenotype; see **Table S2** and **Figure S2** for details.

### Characteristics of genomic disorder loci

Following our meta-analyses to identify rCNV segments associated with disease at strict genome-wide significance (**Figure 3A**), we next sought to characterize the features contributing to dosage sensitivity across the genome. To accomplish this, we compiled a comprehensive set of likely disease-relevant large rCNV segments by integrating our 49 genome-wide significant segments with the curated list of 114 GDs previously reported in the literature. While 80% (91/114) of these literature-based GDs lacked sufficient evidence across cohorts and phenotypes in our dataset to meet our likely conservative criteria for genome-wide significance, we nevertheless found that that 79 of the GDs below genome-wide significance were at least nominally significant (P<0.05) for at least one phenotype in our meta-analyses (**Figure 3B**; **Figure S5C-D**; **Figure S7**). Thus, we combined our 49 genome-wide significant segments with the 79 nominally significant GDs to build a consensus set of 128 disease-relevant rCNV segments for subsequent analyses, which we also provide as a resource for future studies (**Table S6**).

**Figure 3.**
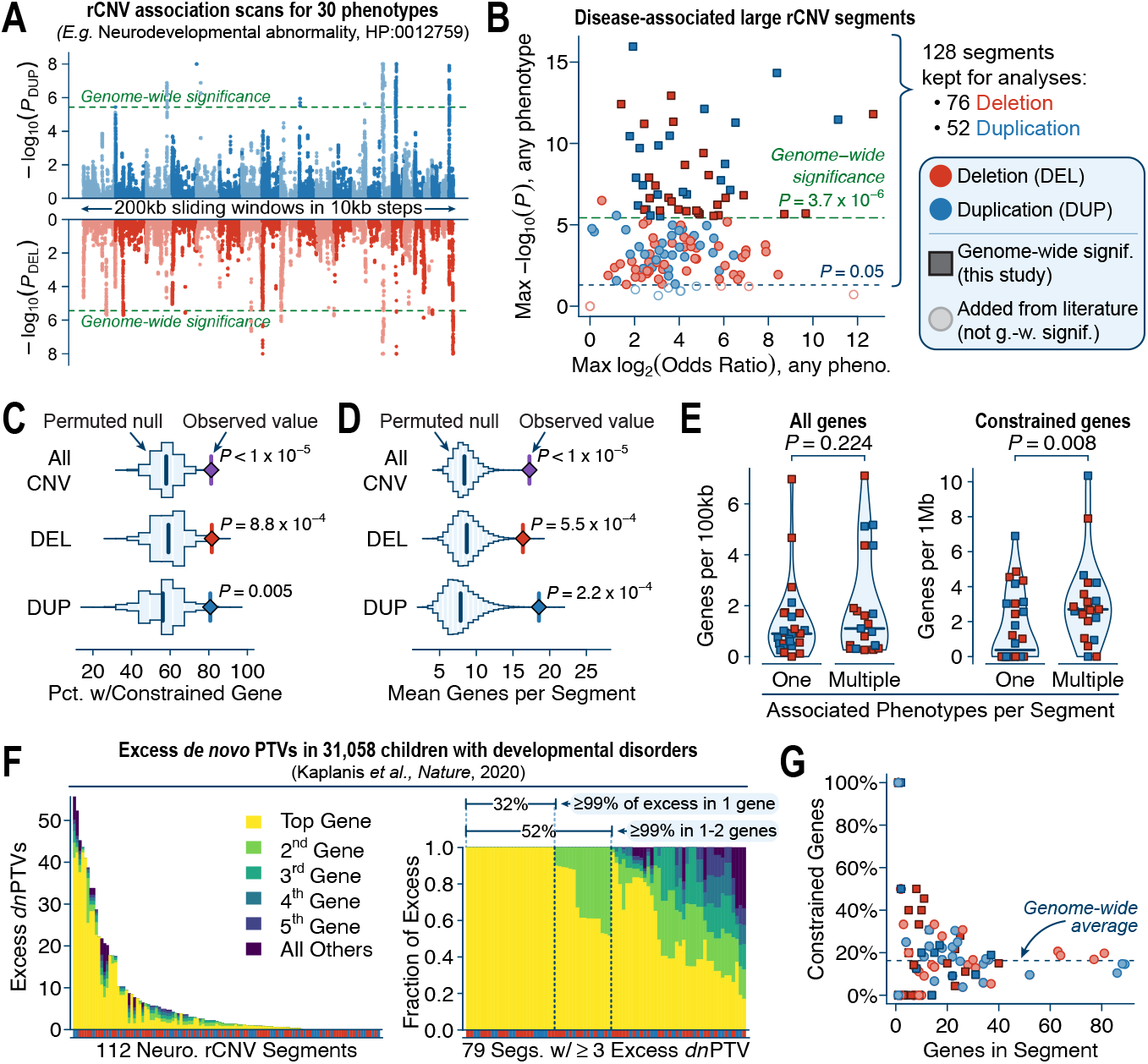
Characteristics of disease-associated large rCNV segments. (**A**) Miami plot of genome-wide rCNV association statistics for one example phenotype, neurodevelopmental abnormalities. (**B**) Relationship between effect size and strength of association for the 49 genome-wide significant segments (dark squares) and 91 reportedly disease-associated rCNV segments curated from the literature (light circles). For subsequent analyses, we retained all 49 genome-wide significant segments plus the 79 literature-based segments that were nominally associated (P<0.05; solid circles) with at least one phenotype in our dataset (total N=128 segments). (**C**) rCNV segments were more likely than expected to overlap at least one mutationally constrained gene based on 100,000 size-matched sets of randomly permuted segments.^50^ (**D**) rCNV segments also overlapped 2.1-fold more total genes on average than expected from 100,000 permutations. (**E**) Genome-wide significant rCNV segments associated with multiple phenotypes (i.e., pleiotropic rCNVs) did not have a greater average density of genes after adjusting for segment size (left) but had increased densities of constrained genes (right) (one-tailed Wilcoxon tests). (**F**) The distributions of de novo protein-truncating variants (PTVs) in 31,058 children with developmental disorders were non-uniform across the genes within most rCNV segments associated with neurological phenotypes.^53^ Shown here is the excess of de novo protein-truncating variants (PTVs) per segment after accounting for the number of genes per segment and gene-specific mutation rates. (**G**) The proportion of constrained genes per rCNV segment was inversely related to the total number of genes in the segment.

A combined analysis of these 128 large rCNV segments revealed that the presence of individual dosage sensitive genes was a common feature that distinguished many of them from the rest of the genome. Over 80% of all segments (104/128) encompassed at least one gene constrained against truncating mutations in the general population,^50^ which was significantly more than the 58% expected by chance based on 100,000 size-matched sets of randomly permuted segments (**Figure 3C**; P<10^−5^, one-tailed permutation test). However, these rCNV segments were also typically gene-dense, overlapping 2.1-fold more genes on average than expected by chance (**Figure 3D**; **Figure S5E**; median=13 genes per segment; P<10^−5^, one-tailed permutation test). Even after accounting for this relatively greater number of genes per segment, we still found that these segments were 1.2-fold more likely than 50 expected to include at least one constrained gene (**Figure S5F**; P=4.8×10^−4^, one-tailed permutation test matched on number of genes per segment) but did not overlap a greater total number of constrained genes per segment (**Figure S5G**; P=0.394, one-tailed permutation test). We also discovered that rCNV segments associated with at least two phenotypes at genome-wide significance (*i*.*e*., pleiotropic rCNVs; N=21) did not exhibit greater densities of genes in general than segments associated with just one phenotype (N=28) (P=0.224, one-tailed Wilcoxon test), but had double the densities of constrained genes (2.0-fold; P=0.008, one-tailed Wilcoxon test) and known disease genes (2.0-fold; P=0.005, one-tailed Wilcoxon test) ^49^ (**Figure 3E**), which may suggest that a greater total number of “driver” genes per segment could be related to the broad phenotypic spectra associated with some GDs.^6^

If the presence of at least one dosage sensitive gene was a common feature of many GDs, we reasoned that this trend should be confirmed by the patterns of damaging protein-coding mutations observed in other patients with related phenotypes, as has been previously proposed.^51,52^ We cross-examined the 113 rCNV segments associated with at least one neurological phenotype versus two datasets of damaging *de novo* mutations (DNMs) from exome sequencing studies of developmental disorders and autism.^52,53^ In both studies, we found that the genes in these 113 segments contained more damaging DNMs in affected individuals than expected from 100,000 permutations adjusted gene-specific mutation rates (**Figure S8**; all P≤0.008, one-tailed permutation tests), which did not clearly differ between protein-truncating and missense DNMs in deletion versus duplication segments. We observed no significant DNM enrichments in the unaffected siblings of autistic children (all P≥0.175). Furthermore, the distributions of damaging DNMs in affected individuals were non-uniform within most rCNV segments (**Figure S9A-D**). For example, when restricting to the 79/113 segments with at least three more protein-truncating DNMs than expected in 31,058 children with developmental disorders,^53^ we found that one-third (32%; 25/79) of such segments had their excess DNMs completely (>99%) concentrated in just a single gene, and half (52%; 41/79) could be explained by no more than two genes (**Figure 3F**). At minimum, this analysis prioritized 25 rCNV segments with orthogonal evidence implicating an individual dosage sensitive gene as a driver of some aspects of their associated neurodevelopmental phenotypes. More generally, these trends were broadly concordant for both truncating and missense DNMs and both deletion and duplication segments, even for many of the largest segments containing dozens of genes (**Figure S9E-F**). Nevertheless, while these distributions of DNMs nominated possible driver genes for up to half of all rCNV segments, the full genetic architecture of most segments is likely to be more complex given the known examples of multiple gene-phenotype correlations within the same segment,^54,55^ gene-gene interactions,^56,57^ and variable penetrance or expressivity due to secondary variants and polygenic background.^6,26,58,59^

Intriguingly, both the proportion of constrained genes and the enrichment of damaging DNMs per segment were inversely related to the total number of genes, as smaller segments showed stronger enrichments for these key features (**Figure 3G**; **Figure S10A**-**B**). We examined whether this pattern might be explained by CNV mechanism: while some GDs formed via non-homologous mechanisms have been focally refined to individual dominant driver genes,^60,61^ NAHR-mediated GDs—which are typically larger and disrupt identical sets of genes in nearly every patient—might be more likely to involve multiple genes with weaker individual effects. To test this hypothesis, we classified the 128 rCNV segments based on predicted mechanism and compared the properties of NAHR-mediated segments (N=60) versus other segments not mediated by NAHR (N=68). As expected, we found that NAHR-mediated segments were 1.4-fold larger on average (**Figure S10C**; P=7.44×10^−7^, two-sided Wilcoxon test), but after correction for size we found no significant differences in densities of constrained genes or known disease genes, enrichments of damaging DNMs in developmental disorders or autism, or average gene expression levels after accounting for multiple comparisons (**Figure S10D**-**H**; N=16 tests; all unadjusted P≥0.048, two-sided Wilcoxon tests). Likewise, we found that damaging DNMs were no more uniformly distributed across the genes in NAHR-mediated segments than in other segments (**Figure S10I**-**J**; all P>0.185, two-sided Wilcoxon tests). Based on the lack of any clear differences, we concluded that NAHR-mediated and other disease-associated rCNVs appear equally likely to contain dominant dosage-sensitive gene(s), and that smaller rCNV segments showed greater enrichments for such genes simply due to their narrow critical regions more precisely pinpointing the underlying driver(s).

### Fine-mapping individual dosage sensitive disease genes within large rCNVs

Our analyses of large rCNV segments indicated that the presence of at least one dominant driver gene was a common feature of many rCNV-disease associations. However, we anticipated that these findings captured just one tail of the total distribution of genic effects across disease-associated rCNVs, ranging from highly penetrant drivers of Mendelian disorders to genes that contribute more modest risk for common and complex diseases.^62^ We thus sought to identify genes enriched for rCNVs in cases over controls by conducting exome-wide rCNV association tests similar to our sliding window analyses. For each autosomal protein-coding gene (N=17,263), we meta-analyzed exonic rCNVs in cases and controls per phenotype while controlling FDR at exome-wide significance (P=2.90×10^−6^) and further requiring nominal evidence (P<0.05) in at least two cohorts (**Figure S11A-B**). In these analyses, we restricted to predicted protein-truncating deletions (≥10% coding sequence [CDS] overlap) and approximately-whole-gene duplications (≥75% CDS overlap). These meta-analyses identified a total of 847 gene-phenotype associations (**File S2**); however, given that many large rCNVs simultaneously deleted or duplicated multiple adjacent genes, we expected that most of these associated genes were not causal but simply carried to significance due to their proximity to true causal genes, analogous to the effects of linkage disequilibrium in conventional genome-wide association studies (GWAS).^62^ To address this problem, we clustered all proximal (±1Mb) significant genes per phenotype and applied a Bayesian fine-mapping algorithm to define the 95% credible set of genes for each association while also prioritizing the most likely causal gene(s) based on their association statistics and 129 gene-level annotations (**Figure 4A**; **Figure S11C-D**; **Figure S12**; **Table S7**).^63,64^

**Figure 4.**
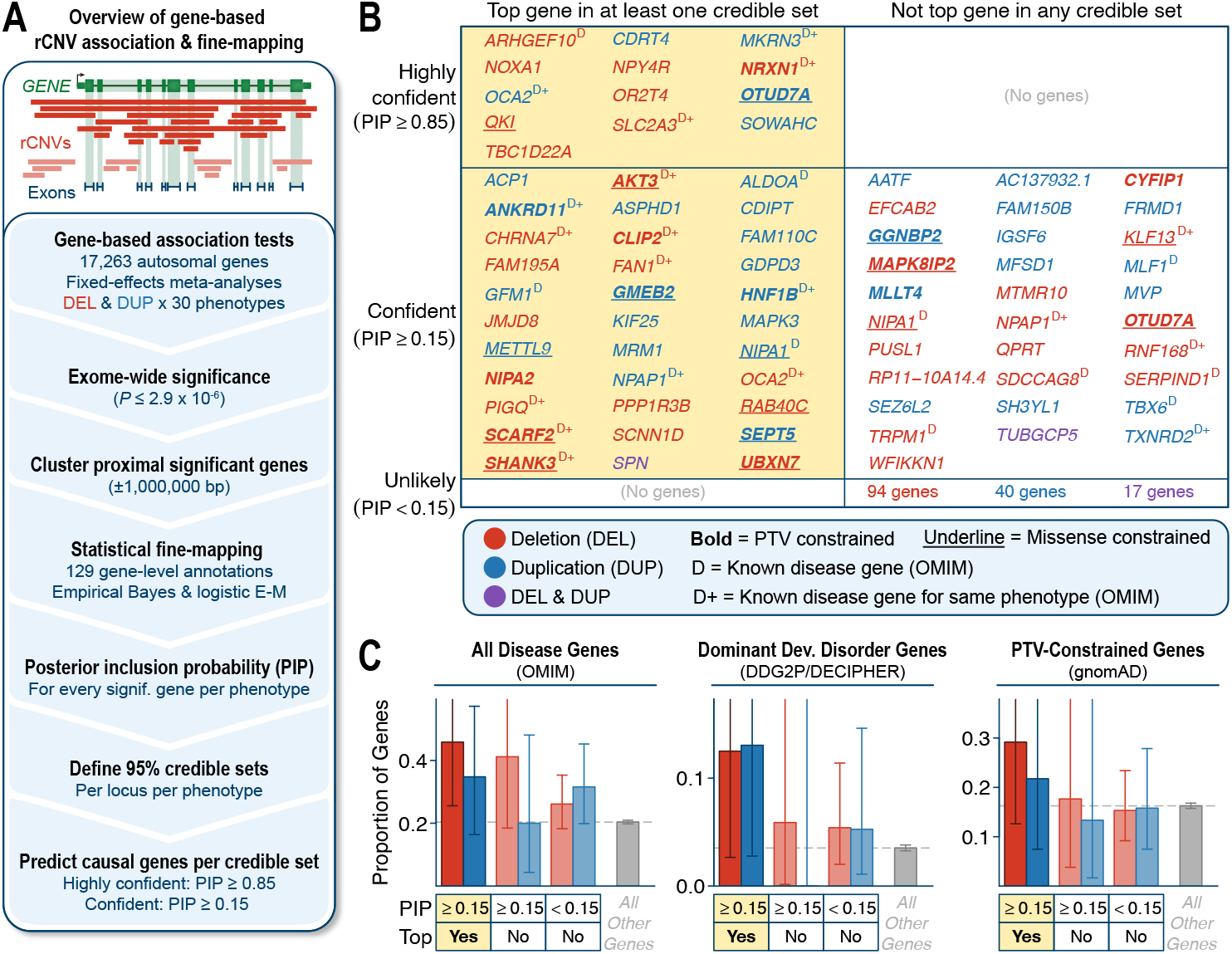
Fine-mapping prioritizes individual genes within large rCNVs. (**A**) Gene-based rCNV disease association & fine-mapping workflow. (**B**) Summary of fine-mapped genes stratified by PIP and whether the gene had the highest PIP (i.e., “top gene”) among all genes in at least one credible set. Gene symbols are colored by CNV association type and are bolded or underlined if they are constrained against PTVs or missense variants, respectively.^50^ Superscript “D” indicates preexisting disease association reports in OMIM,^49^ with “D+” further indicating that the association in OMIM corresponds to at least one of the same phenotypes associated with the gene in this study. (**C**) Proportions of gene groups from (B) that also had at least one reported disease association in OMIM, were reported in the DDG2P/ DECIPHER database as the cause of a Mendelian dominant developmental disorder,^24^ or were constrained against PTVs. Bars indicate binomial 95% confidence intervals. For each panel, the background average of all autosomal genes not contained in any credible set is provided in grey.

Fine-mapping reduced the average number of genes per association by 21%, resulting in a total of 85 credible sets averaging 7.8 genes each (range: 1-25 genes) (**Table S8**). These credible sets typically had strong effect sizes (mean odds ratio=10.4) but varied considerably by locus and phenotype (odds ratio range=1.5-213). Across all credible sets, we identified at least one disease association for 212 unique genes and prioritized 73 “confident” and 13 “highly confident” genes with fine-mapped posterior inclusion probabilities (PIPs) ≥0.15 and ≥0.85, respectively, in at least one credible set (**Figure 4B**; **Figure S11E**; **Table S9**). Most (62%; 45/73) of the confident fine-mapped genes were also the top-ranked gene in at least one credible set and these 45 top-ranked confident genes were enriched for known disease genes (odds ratio [OR]=2.6; P=0.003, two-sided Fisher’s exact test),^49^ especially dominant developmental disorder genes (OR=4.2; P=0.005),^24^ and trended towards stronger constraint against truncating mutations in the general population (OR=1.9; P=0.07) (**Figure 4C**).^50^ These enrichments confirmed that our fine-mapping approach successfully prioritized plausible driver genes, although we did not expect that all true causal genes must match these criteria, such as those responsible for incompletely penetrant rCNV associations with relatively modest effect sizes.

Essentially all (96.5%; 82/85) of the exome-wide significant credible sets overlapped large rCNV segments already identified by our previous sliding window analyses. However, fine-mapping allowed us to nominate candidate genes for 61.2% (30/49) of genome-wide significant rCNV segments (**Figure S13**). These prioritized genes included 27 with documented roles in disease,^49^ like *SHANK3* (PIP=0.27), the cause of Phelan-McDermid syndrome,^65^ which was the top-ranked gene among 19 genes in a 1.2Mb deletion segment associated with neurodevelopmental disorders (OR=64.1; 95% confidence interval [CI]=19.3-214.9). These analyses also prioritized 18 genes that were mutationally constrained but had no known roles in disease, including novel candidate triplosensitive genes like *GMEB2* in nervous system abnormalities (PIP=0.77; OR=21.5; 95% CI=8.2-56.8; **Figure 5A**). However, gene prioritization by fine-mapping was not limited to individual constrained drivers of potentially monogenic phenotypes. For example, we found several associations with modest effect sizes but existing biological evidence supporting possible roles for the candidate gene in their associated phenotypes, including deletions of *SLC2A3* (PIP=1.0) in seizures (OR=2.7; 95% CI=2.1-3.3) and deletions of *NOXA1* (PIP=0.89) in cardiovascular disease (OR=5.9; 95% CI=4.2-8.2). Both *SLC2A3* and *NOXA1* are relatively tolerant of coding variants in the general population,^50^ but deletions of *SLC2A3* have been proposed as a neurodegenerative risk factor and produce abnormal neuronal activity in mice,^66,67^ while common variation near the *NOXA1* locus has been associated with vascular traits like blood pressure and the NOXA1 protein activates NAPDH oxidase, a key enzyme in many cardiovascular diseases.^68,69^ Lastly, our analyses prioritized three genes within duplication associations that had previously been shown to be dominant genetic causes of diseases via haploinsufficiency, like *ANKRD11* (PIP=0.57) in a five-gene credible set associated with growth abnormalities (OR=16.6; 95% CI=11.6-24.1) (**Figure 5B**). Haploinsufficiency of *ANKRD11* causes Cornelia de Lange and KBG syndromes but the duplications in this study suggest that *ANKRD11* is not only haploinsufficient but is bidirectionally dosage sensitive.^70,71^ Collectively, these results demonstrated that fine-mapping algorithms originally designed for GWAS can be readily reconfigured for rCNV association frameworks to refine large rCNV segments and prioritize candidate genes across a range of phenotypes and effect sizes.

**Figure 5.**
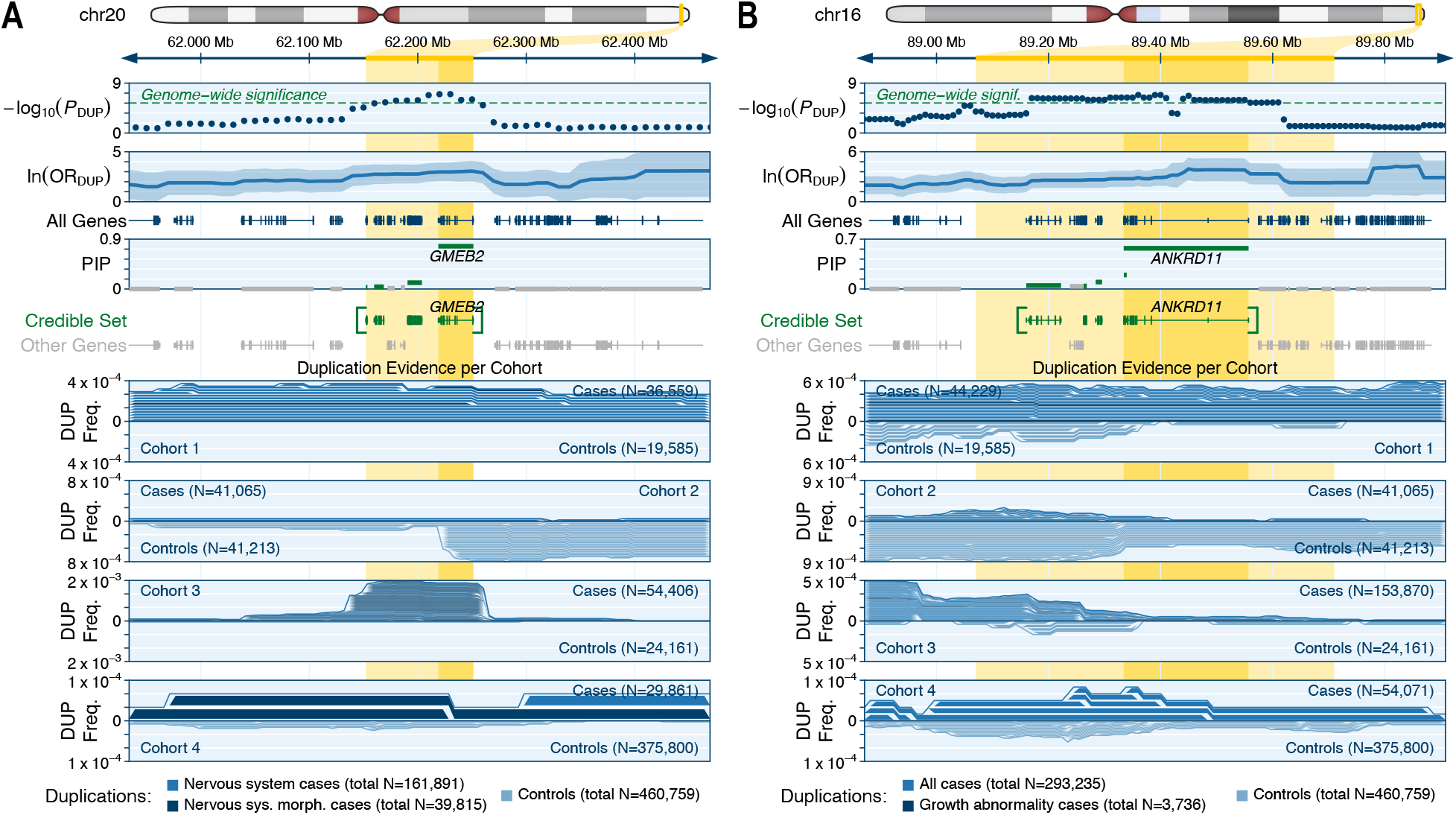
Novel candidate triplosensitive disease genes revealed by fine-mapping. (**A**) We identified a 95% credible set of four genes on chromosome 20 where rare duplications were associated with nervous system abnormalities (OR=21.5; 95% CI=8.2-56.8). Fine-mapping prioritized GMEB2 as the likely driver gene for this association (PIP=0.77), which to our knowledge represents the first association between rare genetic variation in GMEB2 and disease. Meta-analysis P-values and ORs are provided for the more specific (smaller N) of the two phenotypes listed at the bottom of the panel, and ORs are also provided with a 95% confidence interval in lighter shading. (**B**) We identified a 95% credible set of five genes on chromosome 16 where rare duplications were associated with growth abnormalities (OR=16.6; 95% CI=11.6-24.1), and fine-mapping prioritized ANKRD11 as the likely driver gene for this association (PIP=0.57). ANKRD11 is an established haploinsufficient gene and a cause of autosomal dominant developmental disorders,^70,71^ but the duplication association identified here suggests that ANKRD11 is likely also triplosensitive.

### Discovering dosage sensitive noncoding regulatory loci

Although most penetrant variants are expected to act via direct alteration of coding sequences,^72,73^ there is a growing list of examples where noncoding rare variants—especially rCNVs—exert strong effects in disease.^74^ Therefore, as a third complementary approach, we scanned the genome for enrichments of rCNVs in cases over controls after depleting our dataset of strong effects attributable to direct alterations of known coding sequences (**Note S2**; **Figure S14A**). Given our limited prior knowledge of which noncoding genomic features were most likely to be dosage sensitive,^75^ we designed an association framework to empirically evaluate 11,612 genome annotation classes (**Figure S14B-C**; **Table S10**). First, we computed the genome-wide burden of rCNVs in cases versus controls per annotation class, which prioritized 397 classes with nominal (P<0.05) enrichments of noncoding rCNVs in cases over controls (**Figure S14D**; **Table S11**). Consistent with previous predictions, the classes most enriched for rare deletions in cases were generally activating regulatory elements, like enhancers, which are typically more constrained against deletions than other classes of noncoding elements (**Figure S14E**).^27^ Next, we clustered the elements from these 397 classes to build 15,497 non-overlapping *cis*-regulatory blocks (CRBs). The median CRB spanned 23.4kb and involved 52 individual elements (**Figure S14F**). Finally, we conducted genome-wide rCNV association meta-analyses for all phenotypes per CRBs using a similar framework as our gene-based association tests while assessing significance at a genome-wide threshold (P=3.23×10^−6^; corrected for 15,497 total CRBs) and requiring nominal evidence of association (P<0.05) in at least two cohorts.

This approach detected just two independent genome-wide significant associations, both of which overlapped loci already identified in our large segment analyses and were therefore robust to the technical details of our noncoding association test. The interpretation of these two noncoding rCNV associations was more challenging than our previous gene-based analyses. For example, we discovered an association between noncoding deletions and hyperactivity (OR=42.1; 95% CI=9.2-191.6) within the very large (∼550kb) first intron of *CADM2*. These noncoding deletions directly overlapped a validated recursive splice site that controls *CADM2* isoform switching in the human brain (**Figure 6A**).^76^ *CADM2* is constrained against truncating variation,^50^ encodes a synaptic cell adhesion protein involved in early postnatal neurodevelopment via direct interactions with neurexins 1-3,^77^ and has been implicated in multiple behavioral phenotypes by common variant GWAS.^78,79^ While our data indicate that intronic *CADM2* deletions increase risk for hyperactivity, experimental validation will be required to clarify the precise mechanism. As a second example, we identified an intergenic locus bookended by the coding genes *PRKCQ* and *SFMBT2* where duplications were associated with neurodevelopmental disorders (OR=5.8; 95% CI=5.0-6.6) (**Figure 6B**). This locus encompassed three lincRNAs, none of which had known roles in neurodevelopment, and several enhancers that were active in embryonic stem cells but largely silent in adult brain tissue; however, we were unable to identify any genes within ±1Mb known to be involved in developmental disorders. While the molecular mechanisms remain uncertain for both of the noncoding rCNV-disease associations detected here, they provide hints at the possible ways by which noncoding rCNVs might contribute disease risk.

**Figure 6.**
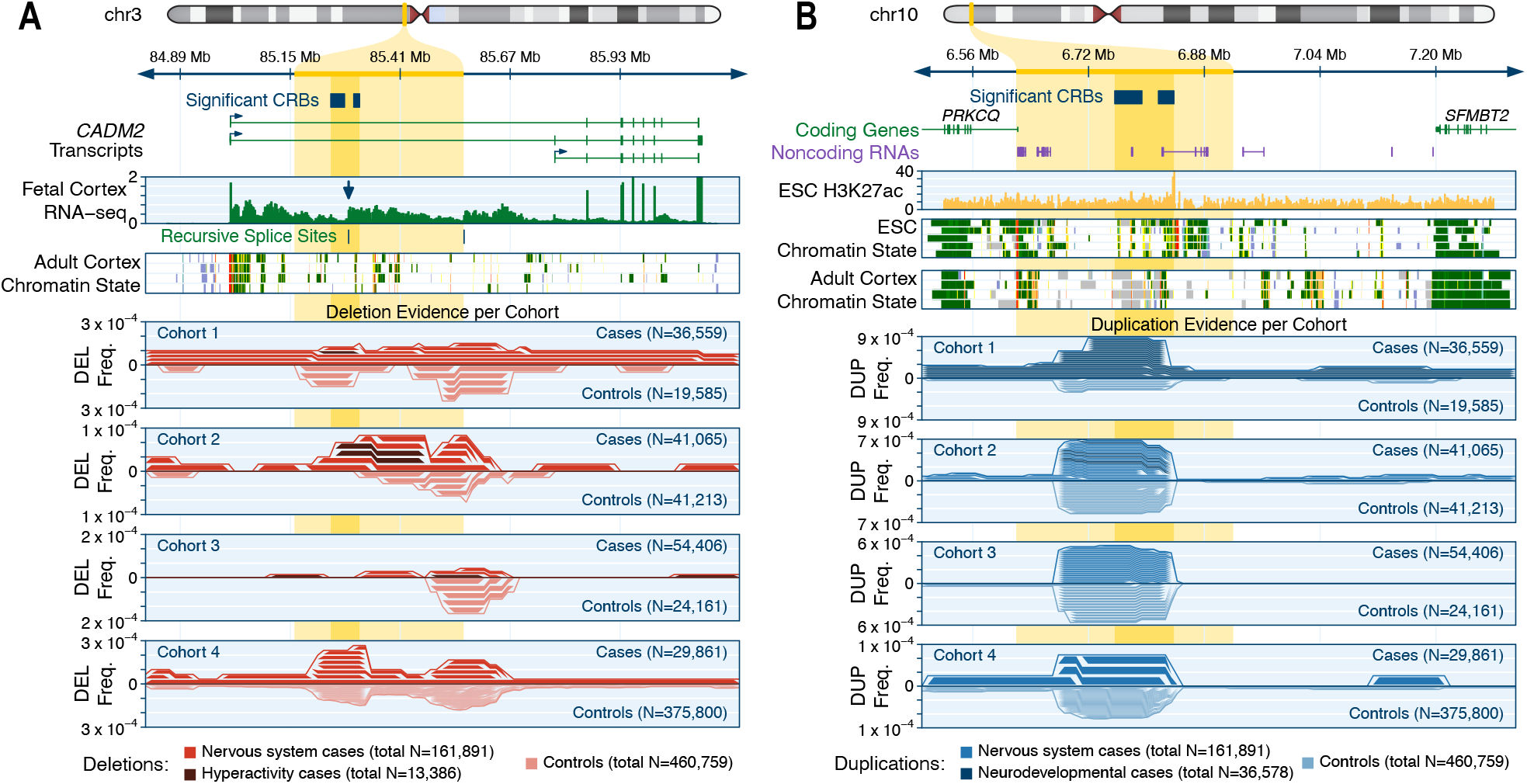
Dosage-sensitive noncoding regulatory regions associated with disease. (**A**) We discovered a genome-wide significant association between hyperactivity and noncoding rare deletions in the first intron of CADM2 on chromosome 3 (OR=42.1; 95% CI=9.2-191.6; light yellow highlight), which involved two significant cis-regulatory blocks (CRBs; dark yellow highlight). These CRBs centered on a molecularly validated recursive splice site conserved across vertebrates responsible for CADM2 isoform switching in the brain.^76,80,81^ (**B**) We discovered a genome-wide significant association between neurodevelopmental disorders and rare duplications at an intergenic locus on chromosome 10 (OR=5.8; CI=5.0-6.6), which also involved two significant CRBs. These CRBs overlap a cluster of three lincRNAs and several annotated enhancers active predominantly in embryonic stem cells (ESCs) and largely inactive in most adult somatic tissues, including adult brain cortex.

### The dosage sensitivity of human genes

The rCNV association analyses conducted in this study provided a dosage sensitivity map of large genomic segments associated with disease, yet they were inherently limited to stringent genome-wide significance thresholds and coarse phenotypes. While disease association studies and clinical interpretation of short variants have been revolutionized by gene-level metrics that estimate intolerance to coding mutations,^50^ there are few comparable metrics that reflect gene-level intolerance to dosage alterations from CNVs. Over the last decade, studies have developed methods for prioritizing likely pathogenic CNVs and estimating selection against CNVs of individual genes,^33,38,82-84^ but there are no widely adopted frameworks to evaluate both haploinsufficiency and triplosensitivity for every human gene. Thus, we reasoned that a catalog of dosage sensitivity scores for all genes—even if imperfect—would provide important insights into the general principles of dosage sensitivity and represent a potentially useful tool for human genetic research and clinical CNV interpretation.

Taking advantage of the increased sample size of our aggregated rCNV dataset relative to previous studies, we developed a two-step procedure to computationally predict the probability of haploinsufficiency (pHI) and triplosensitivity (pTS) for every autosomal protein-coding gene. We first used an empirical Bayes approach to compute the likelihood that each gene belonged to one of two manually curated sets of likely dosage sensitive and insensitive genes based solely on the summary statistics from our gene-level association meta-analyses (**Figure S15A-D**), with slight modifications and optimized parameters such as enriching for focal whole-gene deletions and duplications (**Note S3**). We then trained a machine learning model to predict these per-gene likelihoods based on 129 gene-level features and protected against overfitting with cross-validated out-of-sample prediction and ensemble averaging across seven different model architectures. This model produced pHI and pTS scores for 17,263 autosomal genes (**Figure 7A**; **Table S12**), which easily separated known dosage sensitive and insensitive genes with high precision and recall (**Figure S15E-F**; precision-recall area under curve [AUC]: pHI=0.929, pTS=0.950). Both pHI and pTS were correlated with gene-level constraint metrics derived from point mutations (**Figure S16**) despite our approach modeling the likelihood that changes in gene dosage will result in severe disease, whereas many existing point mutation-derived constraint metrics infer purifying selection based on a lack of variation observed in healthy individuals. Finally, given that the effects of deletions are typically stronger than duplications, we computed standardized cutoffs for pHI and pTS where the average effect sizes of deletions or duplications were as strong as deletions of genes known to be constrained against truncating point mutations (average OR≥2.1).^50^ Applying these cutoffs defined 3,006 haploinsufficient (pHI≥0.84) and 295 triplosensitive (pTS≥0.993) genes with empirical rCNV effect sizes as strong as deletions of gold-standard constrained genes (**Figure 7B**).

**Figure 7.**
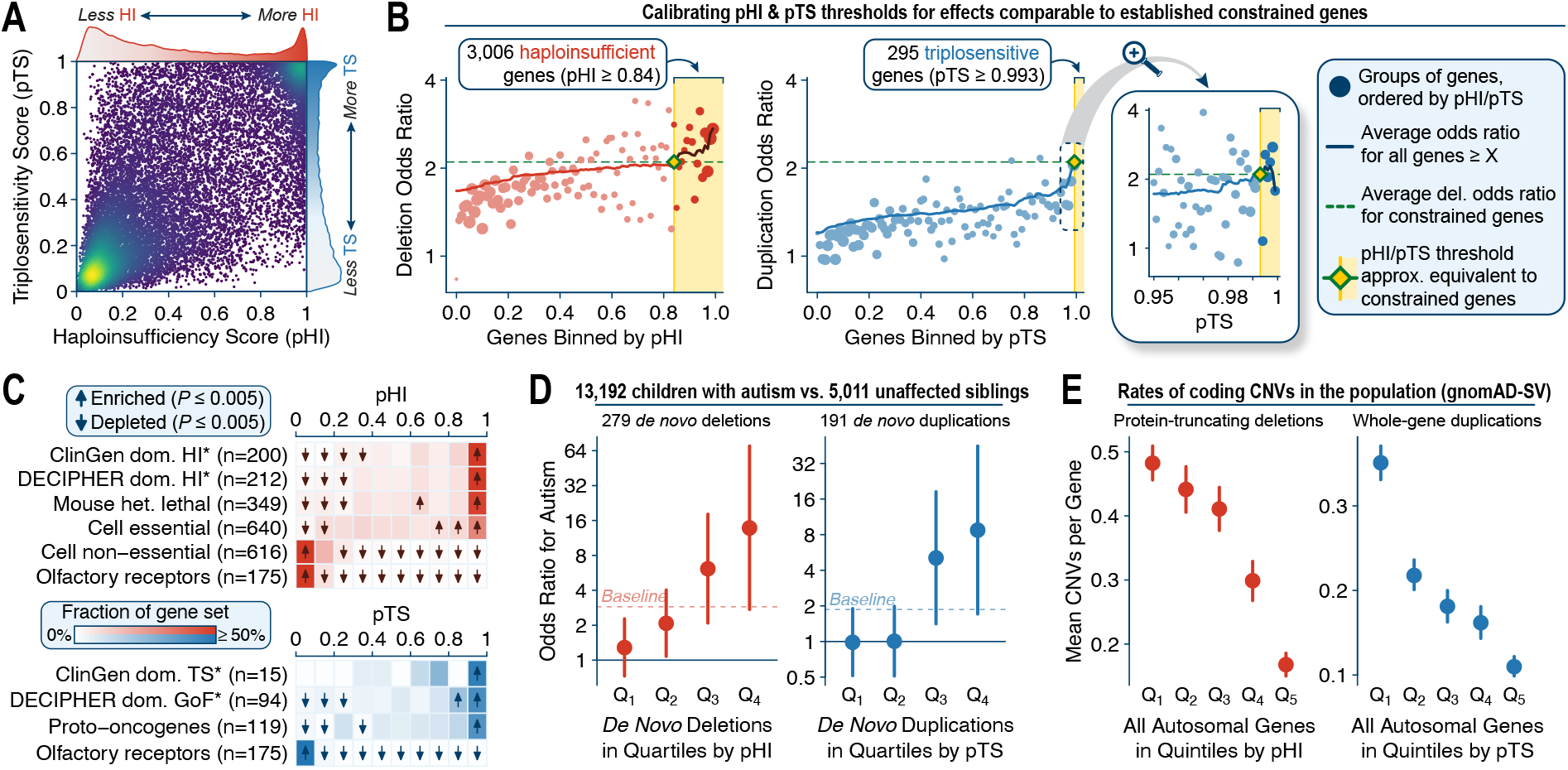
Ensemble machine learning predicts dosage sensitive genes. We developed a machine learning model to predict the probability of haploinsufficiency (pHI) and triplosensitivity (pTS) based on 129 gene-level features for 17,263 autosomal protein-coding genes. (**A**) pHI and pTS were moderately correlated (Pearson R^2^=0.48). (**B**) We calibrated thresholds for pHI and pTS to define 3,006 haploinsufficient and 295 triplosensitive genes where the effect sizes of deletions or duplications were comparable to an established set of genes constrained against truncating point mutations.^50^ These thresholds were set to the lowest pHI or pTS score where the average odds ratio for deletions or duplications computed from our rCNV dataset was at least as strong as the average deletion odds ratio for constrained genes. (**C**) We observed clear shifts in the distributions of pHI and pTS across gene sets with prior biological evidence as being dosage sensitive or insensitive. Gene sets marked with asterisks were considered as criteria when building training sets and thus are not completely independent test sets. (**D**) pHI and pTS stratified risk for autism spectrum disorder conferred by de novo protein-truncating deletions and whole-gene duplications outside of GDs as identified from exome sequencing in 13,192 affected children and 5,011 unaffected siblings (**Note S4**).^52^ Baseline indicates the average odds ratio across all deletions or duplications without stratifying on pHI or pTS. (**E**) pHI and pTS were inversely correlated with rates of protein-truncating deletions and whole-gene duplications in the general population.^27^

We assessed the quality and practical value of these new dosage sensitivity scores using four orthogonal approaches. First, pHI and pTS were predictive of genes with biological evidence for being haploinsufficient or triplosensitive independent of our training criteria, including proto-oncogenes (P<10^−100^, two-sided Kolmogorov-Smirnov [KS] test of pTS),^85^ genes essential in human cell culture (P<10^−100^, KS test of pHI),^86^ or genes whose homologs are embryonically lethal when heterozygously inactivated in mice (P<10^−100^, KS test of pHI) (**Figure 7C**).^87^ Second, pHI and pTS stratified risk for autism conferred by *de novo* CNVs outside of established GDs in 13,192 affected children and their 5,011 unaffected siblings: for example, the top quartile of *de novo* deletions and duplications when ranked by pHI and pTS conferred an order of magnitude greater risk for autism when compared to the bottom quartile (deletions=10.8-fold increased risk; duplications=8.9-fold increased risk; **Figure 7D**; **Figure S17A-C**; **Note S4**).^52^ Third, both pHI and pTS were inversely correlated with rates of protein-truncating deletions and whole-gene duplications in the general population based on an independent catalog of CNVs from genome sequencing (**Figure 7E**; **Figure S17D-F**).^27^ Fourth, genes with high pHI and pTS scores had significant excesses of damaging DNMs and chromosomal rearrangements in individuals with developmental disorders, while we observed no such enrichments for damaging DNMs in unaffected individuals (**Figure S17G-J**).^52,53,88^ These four analyses collectively indicated that pHI and pTS were well-calibrated and predicted dosage sensitive genes throughout the human genome. Furthermore, we repeatedly observed that pTS was more effective than pHI and many existing gene-level metrics when specifically classifying triplosensitive loci (**Figure S15G-H; Figure S17A-C**,**F**), indicating that pTS may provide *in silico* support to the challenges of interpreting duplications in clinical genetics.^89^

Satisfied with their technical quality, we leveraged these scores to understand the general properties governing the dosage sensitivity of human genes. Consistent with prior analyses of population-scale sequencing,^27,84^ we found that haploinsufficiency and triplosensitivity were generally correlated per gene (R^2^=0.482; P<10^−100^, Pearson correlation test). To identify features most predictive of dosage sensitive genes, we designed two complementary elastic net regressions. First, we evaluated the minimum of pHI and pTS per gene to determine the features underpinning bidirectional dosage sensitivity (**Figure 8A**). Not surprisingly, we found that bidirectionally dosage sensitive genes were defined by their evolutionary conservation and constraint against disruptive coding mutations above all other features (**Figure 8B**). Second, to understand the properties distinguishing haploinsufficient and triplosensitive genes, we considered the difference of pHI and pTS scores among the 8,417 genes with some evidence of being dosage sensitive (pHI or pTS ≥ 0.5) (**Figure 8C**). This approach revealed that genes more sensitive to copy loss than gain (*i*.*e*., primarily haploinsufficient genes) tended to be larger, farther from other genes, and with a greater number of conserved enhancers in *cis*, all of which are classic hallmarks of precisely regulated, developmentally critical genes (**Figure 8D**).^90-92^ Conversely, genes more sensitive to gain than loss (*i*.*e*., primarily triplosensitive genes) were generally shorter, G/C-rich genes located in gene-dense, highly active regions not particularly enriched for conserved enhancers (**Figure 8E**). While preliminary, these analyses represent an initial step towards understanding the principles of genic dosage sensitivity and decoupling the mechanisms of haploinsufficiency from triplosensitivity in the human genome.

**Figure 8.**
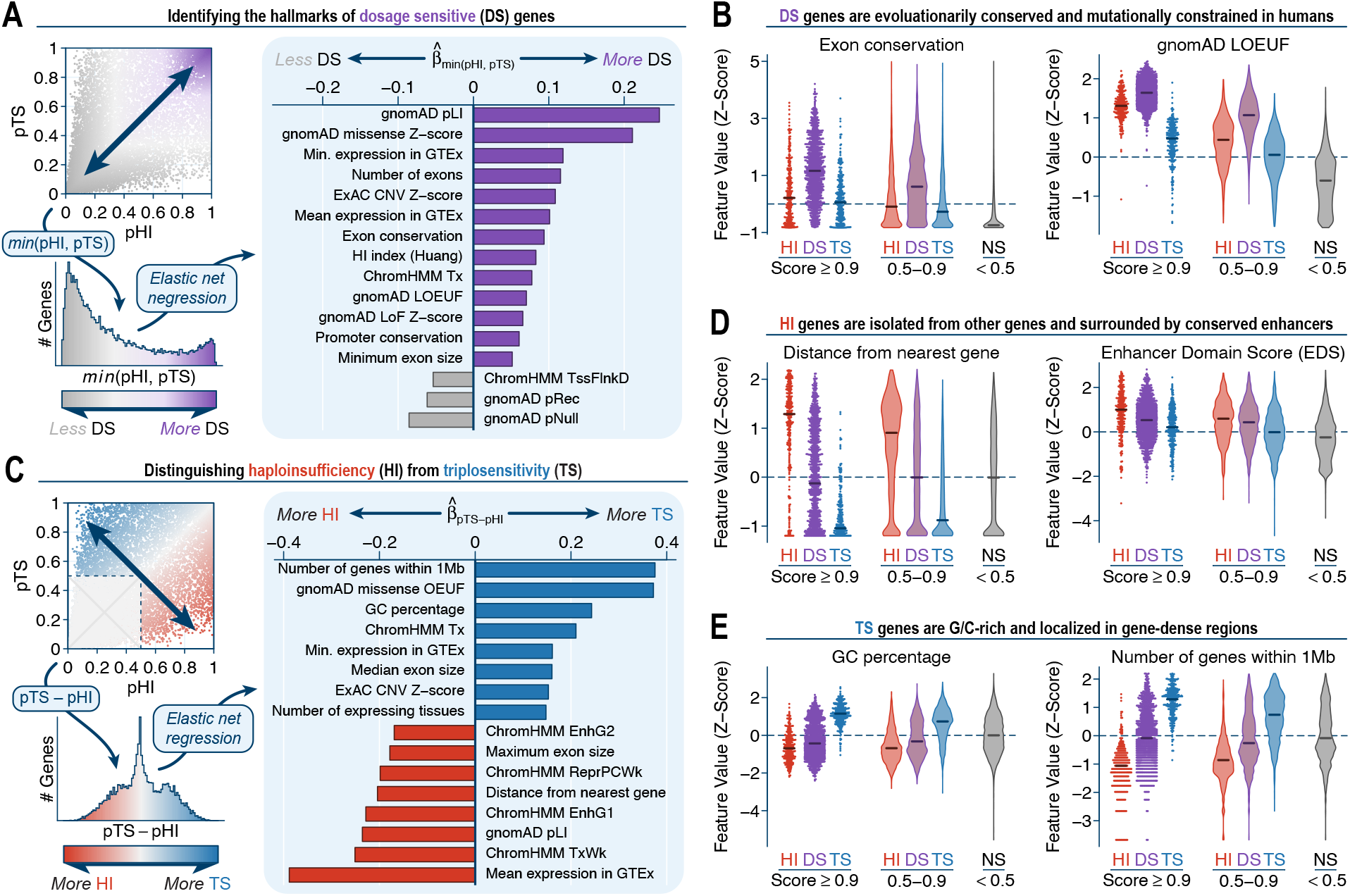
Insights into the dosage sensitivity of human protein-coding genes. **(A)** We identified features predictive of bidirectionally dosage sensitive (DS) genes with a penalized regression of the minimum pHI & pTS value per gene versus all 129 gene-level features. Shown here is an outline of the regression approach and the 16 gene-level features with the largest standardized coefficients in the fitted model. (**B**) Distributions of selected gene-level features for categories of genes classified as haploinsufficient (HI), DS, triplosensitive (TS), and not sensitive (NS) at various thresholds. For clarity, all features have been transformed into Z-scores. (**C**) We also identified features predictive of genes uniquely HI or TS (but not both) using a penalized regression model similar to (A). (**D**-**E**) See (B).

## DISCUSSION

In this study, we systematically assessed the contribution of rCNVs across human disorders by meta-analyzing a large compendium of publicly available biomedical data. Our analyses emphasized the potent and complex roles of rCNVs in disease, expanded existing knowledge of duplications, and enabled initial cross-disorder maps of dosage sensitivity throughout the genome. We built on decades of seminal CNV studies to assemble the largest rCNV dataset to date, which allowed us to produce a genome-wide catalog of rCNV disease-association statistics for all 200kb windows and protein-coding genes across 30 disease phenotypes that can be mined by future studies to test hypotheses and uncover new biological insights. Also included in this catalog was a consensus reference list of large, dosage sensitive genomic segments involved in human disease, including a high-confidence subset of 49 at stringent genome-wide significance. The >40-fold difference in the number of genome-wide significant associations detected by our large segment (N=102 associations) and gene-based (N=85) analyses as compared to our noncoding analysis (N=2) supports the hypothesis that most penetrant disease-associated rCNVs act via direct disruption of one or more protein-coding genes. We further showed that a substantial fraction of all GDs likely harbor at least one dosage-sensitive driver gene based on enrichments of constrained disease genes as well as the nonuniform distributions of damaging DNMs within rCNV segments. Indeed, a general framework of one driver gene per phenotype for many GDs is in agreement with the increased density of constrained genes we observed for pleiotropic rCNVs and matches examples like the 22q11.2 DiGeorge/Velocardiofacial Syndrome deletion where *CRKL* and *TBX1* have been implicated as drivers of kidney and heart abnormalities, respectively.^54,55^ Perhaps more surprising was the lack of any obvious differences among the types or patterns of genes within NAHR-mediated GDs versus those not mediated by NAHR, which may suggest that the pathogenic mechanisms of these two subgroups of rCNVs are more similar than previously appreciated. If true, an important corollary is that it may be feasible to identify driver genes for many unsolved GDs with convergent genomics and/or molecular experiments. Recent breakthroughs in genome-editing technologies like CRISPR/Cas9 may facilitate the molecular dissection of the elements within individual CNVs or GDs,^56,93,94^ although these approaches cannot yet establish *in vivo* physiological relevance for humans. Thus, we anticipate that a combination of experimental and human genetic approaches will be necessary to further clarify the architecture of large rCNVs.

The statistical methods applied in this study highlighted differences between CNV-based association studies and traditional short variant GWAS. There are two major challenges for most conventional GWAS: (1) identifying causal variants among the dozens of linked variants per locus, and (2) predicting the gene(s) affected by each causal variant.^62^ This contrasts sharply with CNV association studies, where the causal variant is obvious—there is typically only one rCNV per locus per individual—and instead the challenge becomes identifying which gene(s) impacted by the rCNV contribute to phenotype expression. Here, we demonstrated that GWAS fine-mapping algorithms can be extended to CNV-based association studies to prioritize individual genes within large rCNV segments across a range of effect sizes and genetic architectures. Unlike conventional gene-based rare variant association studies where each variant usually affects just one gene, the vast size of most rCNVs allowed us to capture strong effect mutation data across most of the genome in a single dataset. In theory, this boosts power for association testing, yet it comes at the cost of the autocorrelation introduced due to many rCNVs overlapping multiple neighboring genes. The patterns we observed by integrating short variant datasets (*e*.*g*., damaging DNMs or mutational constraint) indicated that short variants and rCNVs frequently converge on the same causal genes at disease-associated loci. Therefore, we expect substantial added value from the eventual unification of all classes of genetic variation into comprehensive association frameworks, which will be best accomplished in large-scale sequencing datasets.

The disease association analyses in this study were inherently limited. The large, microarray-based rCNVs we analyzed here will oversimplify complex and multiallelic rCNVs, which play an increasingly recognized role in human traits and diseases.^95-97^ Similarly, this study did not consider smaller (<100kb) rCNVs, which will be an exciting area for future research in exome and genome sequencing studies as sample sizes increase. Sequencing studies will also be better suited to evaluate rCNVs with precise breakpoint coordinates and more nuanced functional predictions. While we implemented multiple safeguards to protect against possible breakpoint imprecision, we have undoubtedly failed to capture all modes by which rCNVs might alter genes or influence disease risk. This oversimplification may be especially true for duplications, the genic consequences of which are diverse.^12^ All of our gene-based duplication analyses were restricted to near-complete gene copy-gain events (*e*.*g*., ≥75% CDS overlap) and thus we did not explicitly consider other forms of gene-disruptive duplications, such as intragenic exonic duplications,^27,98^ which likely have different properties and genic consequences. Lastly, the phenotype standardization procedures we used here were imperfect, but necessary to boost statistical power and succeeded in detecting many known associations. Future studies with access to rich clinical metadata, such as from electronic health records, will likely identify many rCNV-phenotype associations undiscernible with our existing cohorts and methods.

Finally, we leveraged these data to investigate a critical topic in contemporary human genetic research: dosage sensitivity. Dating back to the initial establishment of a haploinsufficiency index to predict the impact of large deletions in the genome,^83^ there have been multiple efforts to predict the functional and/or pathogenic impact of CNVs from microarray and sequencing data.^33,38,82,84^ The vast genetic and functional datasets now publicly available have enabled us to extend from previous approaches and begin to explore predictions of bidirectional dosage sensitivity in disease on an individual gene level with sample sizes at least an order of magnitude larger than prior studies. Although we were limited by the resolution of the large rCNVs aggregated here, we developed a machine learning model that successfully discriminated individual dosage sensitive genes from those tolerant of changes in copy number and defined lists of 3,006 haploinsufficient and 295 triplosensitive genes under strong purifying selection comparable to protein-truncating mutations in gold-standard constrained genes. Our novel dosage sensitivity scores represent an immediately useful tool for gene and variant prioritization in medical genetics, as evinced by their ability to stratify risk for autism conferred by both *de novo* deletions and duplications.^25^ Our triplosensitivity scores in particular may provide a new lens when interpreting some disease-associated missense variants, for which gain-of-function and loss-of-function consequences are challenging to distinguish *in silico*.^99^ Furthermore, while our model did not directly estimate natural selection against dosage changes in genes—as opposed to many existing gene-level constraint metrics derived from short variants, where null mutation rate models are well established—they nevertheless were effective in clarifying some of the biological patterns underlying selection against CNVs in humans. It has been shown that constraint metrics derived from point mutations would also predict genes intolerant of CNVs,^27,50,84^ but the trends distinguishing haploinsufficient and triplosensitive genes were more intriguing. Specifically, our model’s prediction that haploinsufficient genes tend to be larger, farther from other genes, and surrounded by more conserved enhancers is a striking archetypical description of critical developmental genes, which are separated from other genes due to their intricate *cis*-regulatory networks and nuanced regulation across tissues and timepoints.^90-92^ The opposite was true for triplosensitive genes, which appeared to typically be small, G/C-rich genes localized to broadly active, gene-dense regions. While these crude patterns are preliminary, they nevertheless provide an important foothold for future investigations of dosage sensitivity at sequence resolution and for decoupling the principles of haploinsufficiency and triplosensitivity throughout the human genome.

## Data Availability

Code Availability: All code used in this study has been provided in a single repository on GitHub (https://github.com/talkowski-lab/rCNV2). Where applicable, scripts have been provided with documentation and help text. We also have provided a Docker image hosted on DockerHub (https://hub.docker.com/r/talkowski/rcnv) and Google Container Registry (https://gcr.io/gnomad-wgs-v2-sv/rcnv) that contains all dependencies necessary to execute the code identically as presented in this study.
Data Availability: Most data generated in this study, including summary statistics from association tests, have been provided as Supplemental Tables or Supplemental Files. Large Supplemental Data Files have been temporarily hosted in a public Google Cloud Storage Bucket until formal publication in a peer-reviewed journal, as described in the Supplemental Information. Data from existing publications or public resources can be accessed according to their original source, as described in the corresponding Methods section detailing their curation. All other data not otherwise described here or in the Methods will be made available upon request.

https://storage.googleapis.com/rcnv_project/public/collins_medrxiv_2021/sliding_window_sumstats.tar.gz

https://storage.googleapis.com/rcnv_project/public/collins_medrxiv_2021/gene_based_sumstats.tar.gz

## METHODS & SUPPLEMENTAL INFORMATION

Detailed methods and supplemental information for this manuscript has been provided online.

## ACKNOWLEDGEMENTS

We thank Leonid Mirny, Jesse Engreitz, and Joe Nasser for useful discussions and feedback; Claire Redin, Jeremiah Wala, Steven Schumacher, and Stephan Sanders for assistance with data access; and Jake Conway and Matt Stone for advice on code development. We are grateful to all of the families at the participating Simons Simplex Collection (SSC) sites, as well as the SSC principal investigators. These studies were supported by the National Institutes of Health grants HD081256, NS093200, HD096326, and MH106826. R.L.C. was supported by NHGRI T32HG002295 and NSF GRFP #2017240332. H.B. was supported by NIDCR K99DE026824. This work was supported by grants from the Swiss National Science Foundation (31003A_182632 to A.R. and 310030-189147, 32473B-166450 to Z.K.). M.E.T. was supported by Desmond and Ann Heathwood.

## AUTHOR CONTRIBUTIONS

Conceptualization: R.L.C., K.M., X.N., C.L., P.M.B., S.S., H.B., M.E.T.; Methodology: R.L.C., J.U., D.P.H., C.L., J.F., K.E.S., K.K., H.F., B.M.N., Z.K., S.S., H.B., M.E.T.; Software: R.L.C.; Formal analysis: R.L.C., S.E., H.B.; Resources and data curation: R.L.C., J.T.G., E.P., L.N., G.K., D.P.H., S.E., K.M., F.U., J.F.G., J.M., D.L., B.M.N., J.C.H., Z.K., N.K., E.E.D., H.H., H.B., M.E.T.; Writing - original draft: R.L.C., H.B., M.E.T.; Writing - review & editing: R.L.C., J.U., C.L., P.M.B., K.E.S., J.F.G., D.L., J.C.H., A.R., Z.K., E.E.D., H.B., M.E.T.; Supervision, project administration, and funding acquisition: R.L.C., J.F.G., H.F., J.M., D.L., B.M.N., J.C.H., A.R., Z.K., N.K., E.E.D., H.H., S.S., H.B., M.E.T.

## DECLARATION OF INTERESTS

The authors declare no competing interests.

## RESOURCE AVAILABILITY

### Code Availability

All code used in this study has been provided in a single repository on GitHub (https://github.com/talkowski-lab/rCNV2), where it is further organized by analysis aims. Where applicable, scripts have been provided with documentation and help text. All major analysis aim has shell code vignettes with example commands for each script. Furthermore, all association analyses and gene scoring procedures have also been provided in workflow description language (WDL) format, which allows the generalized redeployment of these scripts using the Cromwell execution engine on cloud computing architectures. Finally, we provide a Docker image hosted on DockerHub (https://hub.docker.com/r/talkowski/rcnv) and Google Container Registry (https://gcr.io/gnomad-wgs-v2-sv/rcnv), which provides a controlled container environment containing all dependencies necessary to execute the code identically as presented in this study.

### Data Availability

Most data generated in this study, including summary statistics from association tests, have been provided as Supplemental Tables or Supplemental Files. Large Supplemental Data Files have been temporarily hosted in a public Google Cloud Storage Bucket until formal publication in a peer-reviewed journal, as described in the Supplemental Information. Data from existing publications or public resources can be accessed according to their original source, as described in the corresponding *Methods* section detailing their curation. All other data not otherwise described here or in the *Methods* will be made available upon request.

## Notes

### Competing Interest Statement

The authors have declared no competing interest.

### Author Declarations

This study was approved by the Partners Healthcare Institutional Review Board Protocol #2013P000323. Data from the UK BioBank was accessed via application #50765 (PI: Talkowski), and data from the Simons Foundation for Autism Research Initiative was accessed via SFARIbase application #573206

