## Supplemental Information for "A cross-disorder dosage sensitivity map of the human genome"

**A cross-disorder dosage sensitivity map of the human genome**
Collins *et al., medRxiv*, January 2021 (version 1)

S1. Data sources, sample counts, and cohort metadata
S2. Standardized phenotypes and sample sizes
S3. Locus-level summary statistics for genome-wide significant large segments
S4. Association-level summary statistics for genome-wide significant large segments
S5. Gene set definitions
S6. Master table of all large rCNV segments considered in analyses
S7. Gene-level features used for fine-mapping and dosage sensitivity scoring
S8. Summary statistics for exome-wide significant credible gene sets
S9. Confident fine-mapped gene-phenotype rCNV associations
S10. Sources for genome annotations used in noncoding association tests
S11. Summary statistics from rCNV burden tests of genome annotation classes
S12. Haploinsufficiency and triplosensitivity scores for autosomal protein-coding genes

**Supplemental Notes**
S1. Determination of genome-wide significance for sliding window meta-analyses....28
S2. Depleting rCNVs for strong effects from direct protein-coding alterations ....28
S3. Parameter optimizations for genic dosage sensitivity modeling....29
S4. *De novo* CNV calls from whole-exome sequencing of autism spectrum disorder families 29

**Supplemental Files**....30
S1. Summary statistics from sliding window association meta-analyses
S2. Summary statistics from gene-based association meta-analyses

**Methods**

1. Data curation

### SUPPLEMENTAL FIGURES

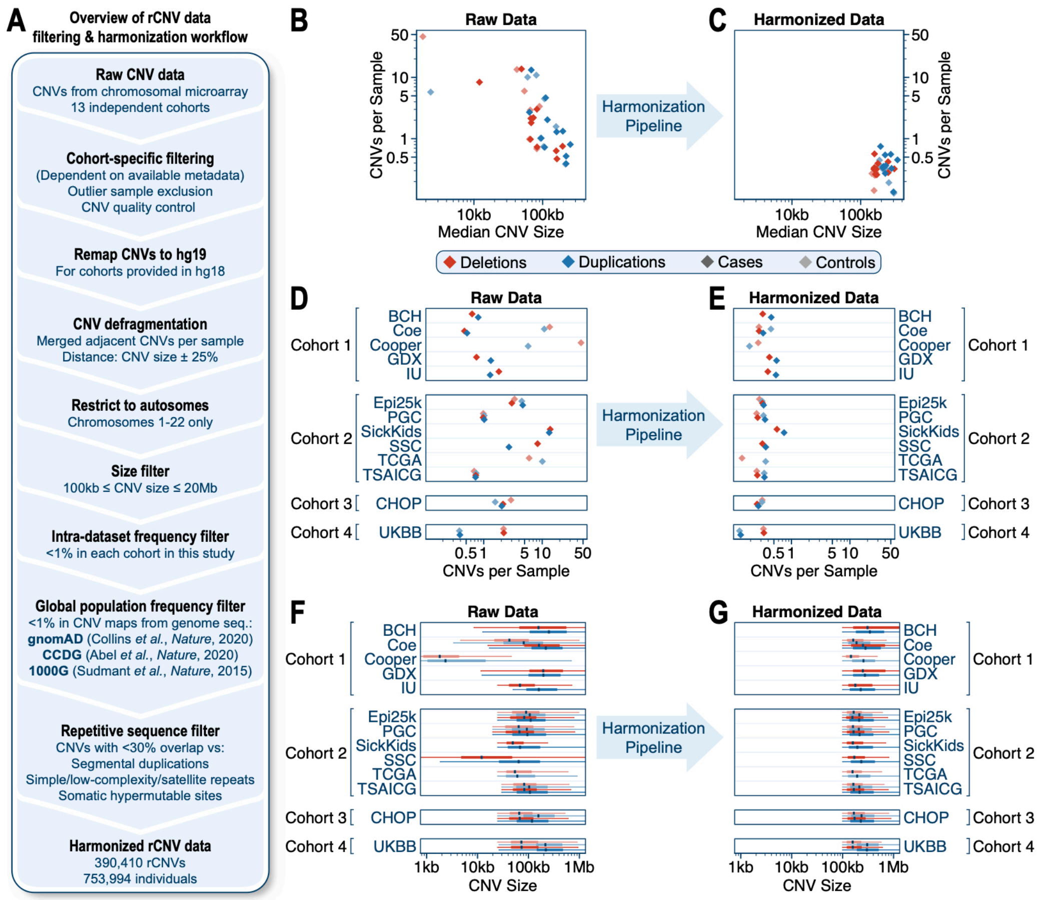

**Figure S1 | rCNV harmonization.** We aggregated & harmonized rCNV data from 753,994 individuals across 13 sources (**Table S1**). (**A**) Overview of rCNV harmonization pipeline. (**B**, **D**, **F**) Selected summary metrics for raw CNV data from each source prior to harmonizing. Sources are grouped by meta-analysis cohort. (**C**, **E**, **F**) Selected summary metrics for rCNV data after harmonization.

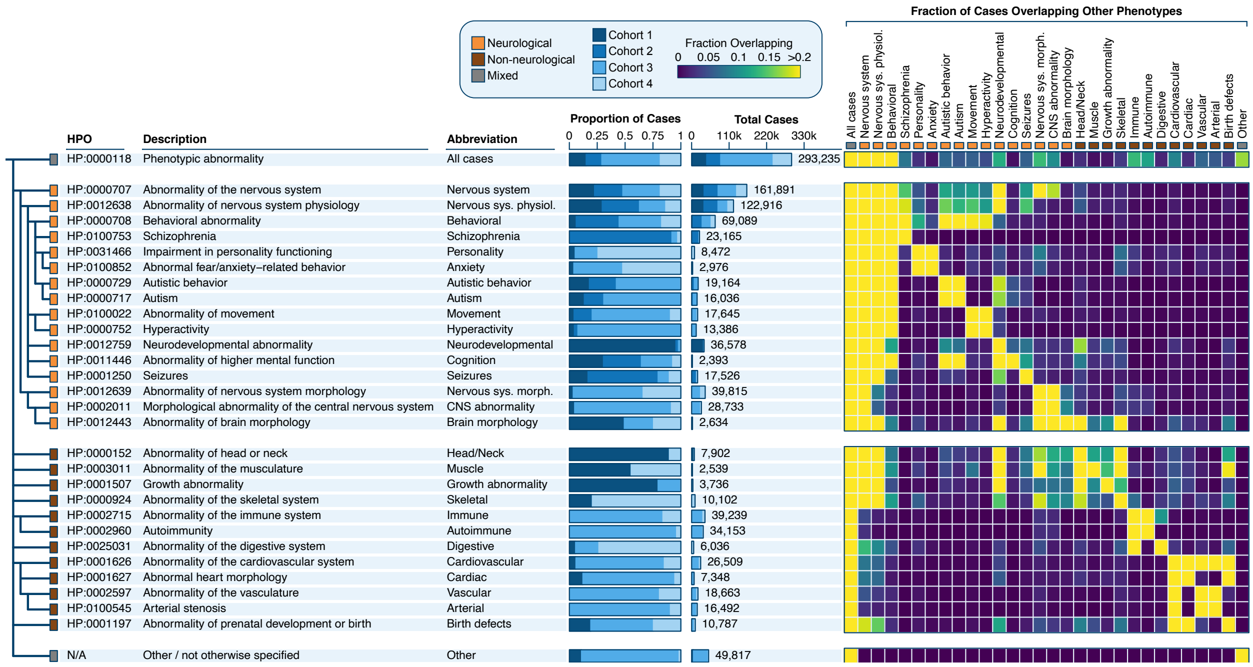

**Figure S2 | Standardized phenotypes.** We standardized phenotype metadata across all cohorts using structured Human Phenotype Ontology (HPO) terms (Kohler et al., 2019). Shown here is the final phenotype hierarchy, split by neurological and non-neurological clades, with the total number of cases per phenotype and the proportion of cases contributed by each of the four cohorts used in meta-analyses. Each phenotype was required to have ≥2,000 total samples, ≥500 samples contributed by two different cohorts, and <50% sample overlap with any other phenotype. Right: asymmetric heatmap of cases overlapping per phenotype. Each row represents the fraction of cases from that phenotype also included in the phenotype indicated on the vertical column axis.

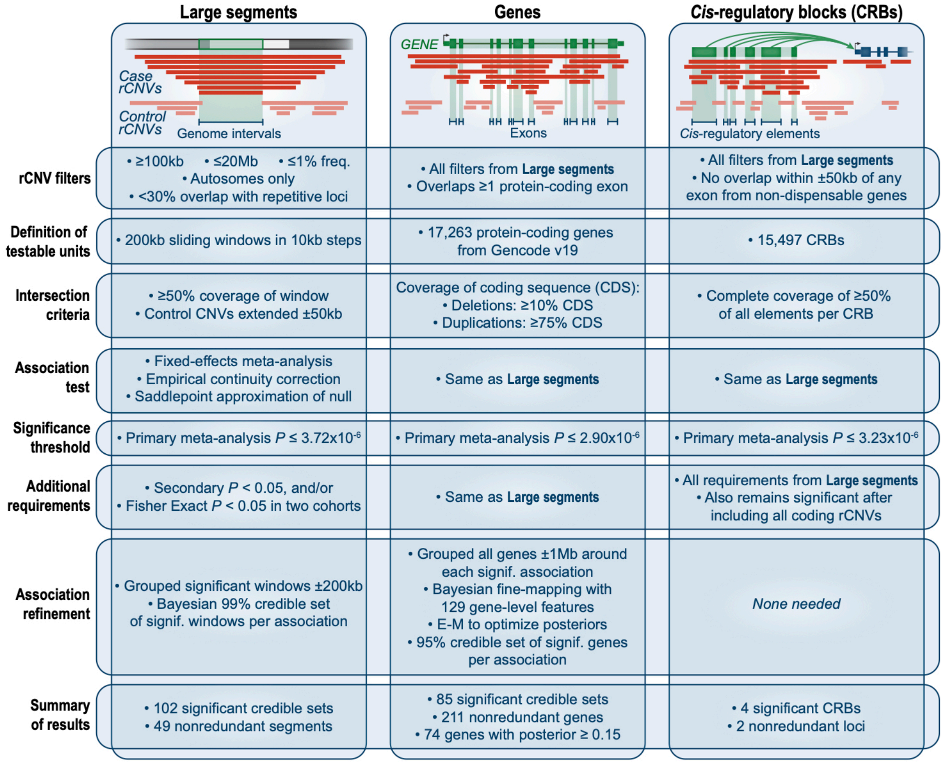

**Figure S3 | Summary of rCNV association models.** We designed three statistical models to search genome-wide for associations between rCNVs and phenotypes. These models are summarized here to highlight their similarities and differences. The predominant difference between the models was their intended test entity: large segments (left column), genes (middle column), or *cis*-regulatory blocks (CRBs; right column). All three models followed similar conceptual frameworks, the steps of which are outlined on the vertical axis.

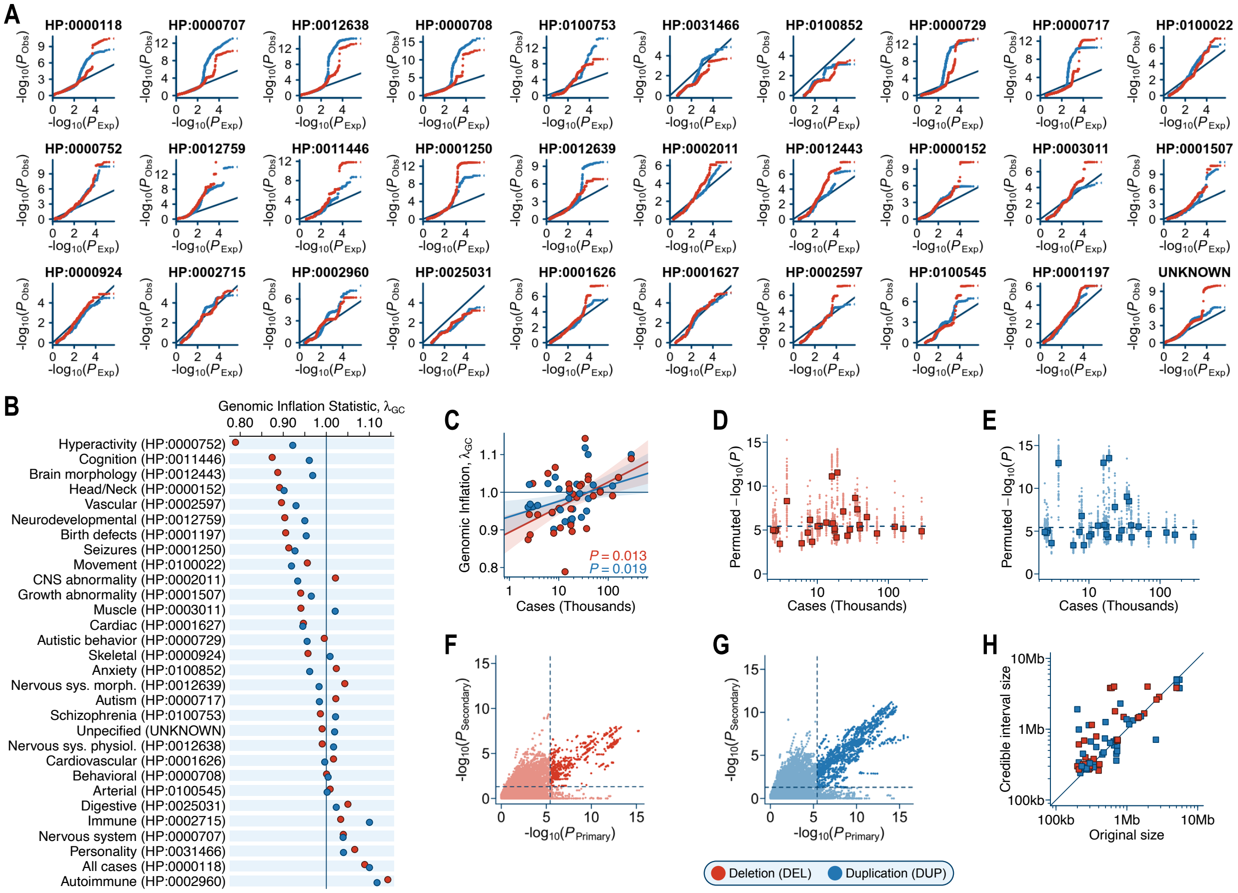

**Figure S4 | Quality control of sliding window association test statistics.** We conducted genome-wide rCNV association meta-analyses for 30 disease phenotypes across all autosomes in 200kb sliding windows in 10kb steps. (**A**) Quantile-quantile plots for meta-analysis test statistics for deletions (red) and duplications (blue) per phenotype. Solid black line denotes the expected distribution if the null hypothesis (*i.e.*, no association) was true for all windows. (**B**) Genomic inflation statistic ($\lambda_{GC}$) for the meta-analysis results from (A), defined as the median observed chi-squared statistic divided by the median chi-squared statistic expected under the null hypothesis. (**C**) Relationship of sample size and $\lambda_{GC}$ following saddlepoint approximation to adjust for case-control imbalance. Trendlines are provided as outlier-robust linear fits for deletions & duplications; shaded areas indicate 95% confidence intervals. (**D**-**E**) We assessed the calibration of our approximated genome-wide significance threshold ($P=3.72\times10^{-6}$; dashed horizontal line) against empirical P-value thresholds derived from 50 random phenotype permutations for deletions (D) and duplications (E) as described in **Note S1**. Small points represent the empirical P-value threshold corresponding to exactly zero false discoveries across all tested windows for a single permutation. Large squares are median P-value thresholds per phenotype across all 50 permutations. (**F**-**G**) Relationship of primary and secondary P-values across all meta-analyses for deletions (F) and duplications (G). Secondary P-values reflect the meta-analysis outcome after excluding the most significant individual cohort per window. Dashed lines indicate thresholds for genome-wide significance, and dark shaded points indicate windows surpassing both thresholds. (**H**) Relationship of original significant locus size versus the size of the 99% credible interval after refinement for each of the 102 final rCNV-phenotype associations at genome-wide significance.

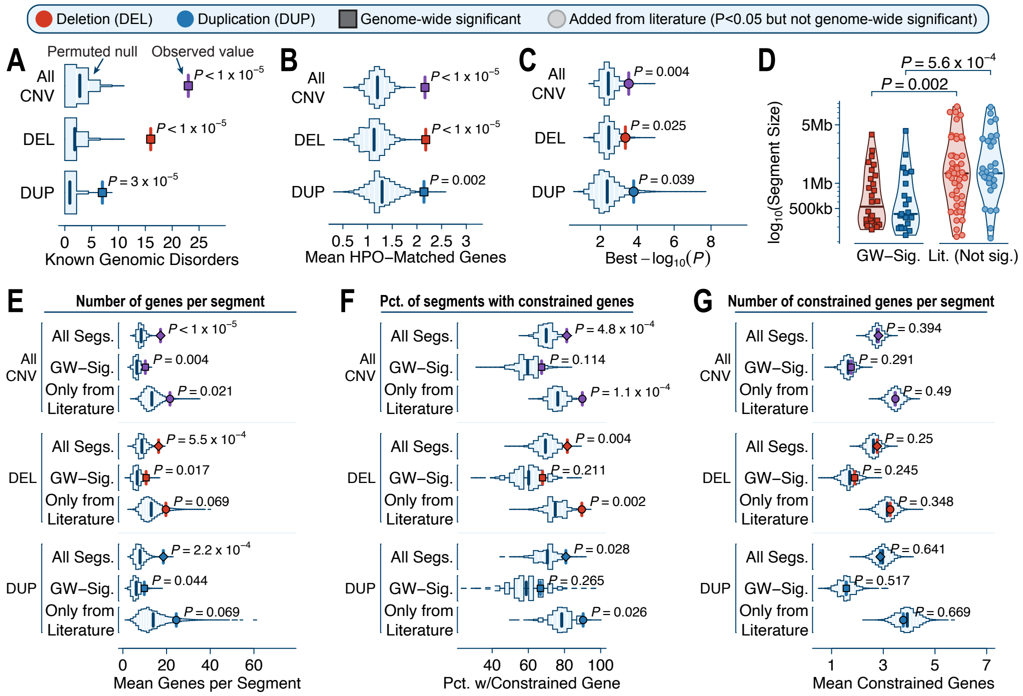

**Figure S5 | Combined analyses of large rCNV segments.** (**A**) We curated a list of 114 GDs reported by at least one of six existing sources (Dittwald et al., 2013; Firth et al., 2009; Girirajan et al., 2012; Owen et al., 2018; Riggs et al., 2012; Stefansson et al., 2014). The genome-wide significant rCNV segments from our discovery analysis overlapped these 114 GDs 8.2-fold more often than expected by chance based on 100,000 random sets of size-matched segments (one-sided permutation test). (**B**) Genome-wide significant rCNV segments overlapped more known disease genes previously implicated in the same phenotypes (Amberger et al., 2015) than expected by chance based on 100,000 random segments while matching on number of genes per segment (one-sided permutation test). (**C**) The 91 previously reported GDs curated from the literature that did not overlap one of our 49 genome-wide significant rCNV segments were significantly enriched for association signals beyond chance expectations from 100,000 random segments (one-sided permutation test, as in A). (**D**) Genome-wide significant segments were smaller on average than literature-reported segments (two-sided Wilcoxon tests). (**E**) rCNV segments overlapped significantly more genes than expected by chance (one-sided permutation test; N=100,000 size-matched permutations, as in B). (**F**) rCNV segments were more likely to overlap at least one mutationally constrained gene than expected by chance (one-sided permutation test; N=100,000 gene-matched permutations, as in B). (**G**) rCNV segments did not overlap a greater total number of mutationally constrained genes than expected by chance (one-sided permutation test; N=100,000 gene-matched permutations, as in B).

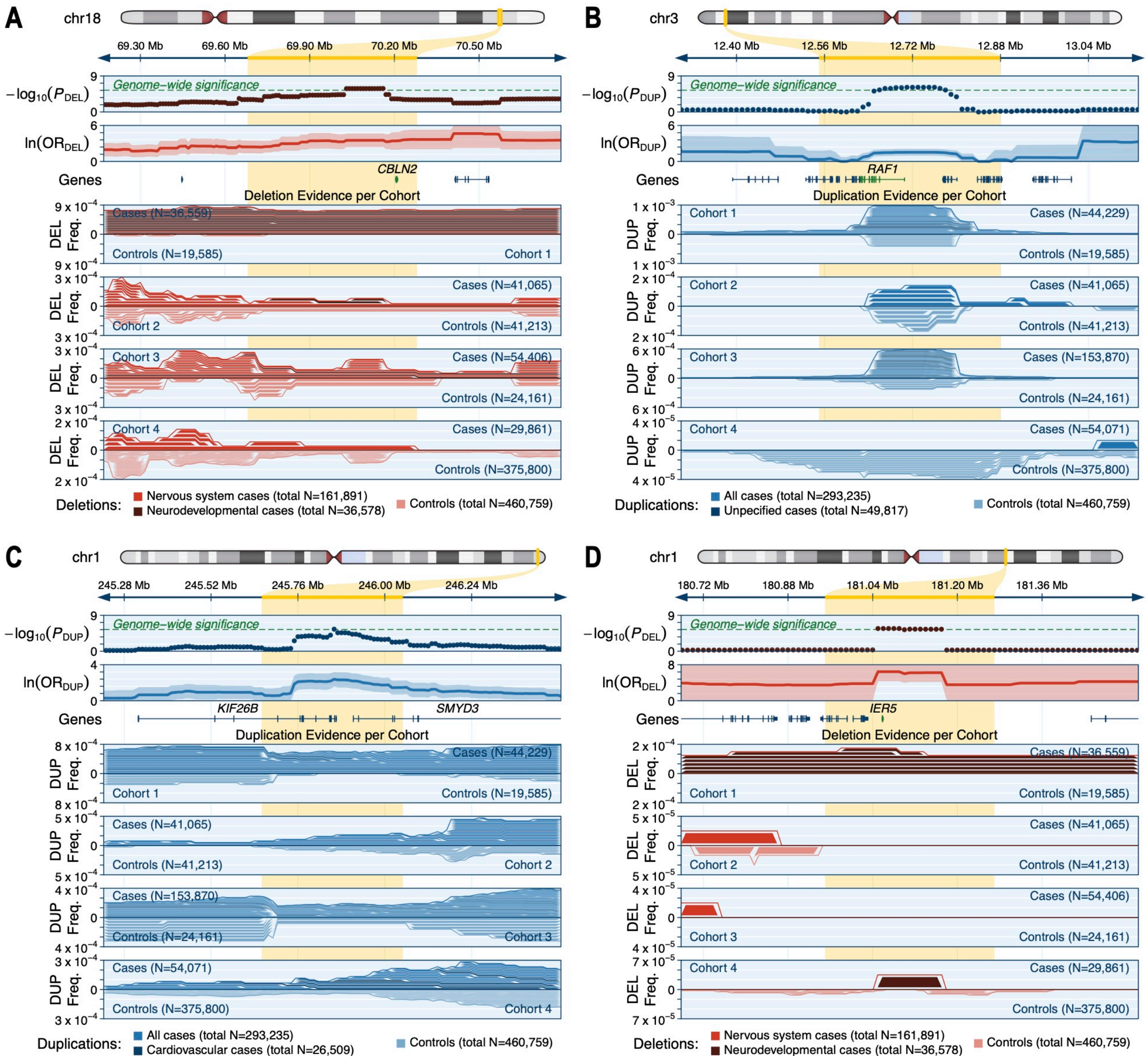

**Figure S6 | Selected loci from large rCNV segment discovery analyses.** (**A**) We discovered a 600kb segment on chromosome 18 where rare deletions were associated with neurodevelopmental disorders. This segment contained just one gene, *CBLN2*, which has recently been implicated in human-specific prefrontal cortex development and neurodevelopmental abnormalities (Shibata et al., 2019). We found multiple gene-specific deletions of *CBLN2* (and no other genes) in affected individuals in our dataset. (**B**) We discovered a 330kb segment on chromosome 3 where rare duplications were associated with disease (*i.e.*, in affected individuals with unspecified phenotypes). This segment encompassed seven genes, including *RAF1*. Heterozygous gain-of-function missense mutations in *RAF1* are a confirmed cause of dominant Noonan and LEOPARD Syndromes (Pandit et al., 2007). (**C**) We discovered a 390kb segment on chromosome 1 where rare duplications were associated with cardiovascular disease. This segment overlapped just two genes: *KIF26B*, which is a mutationally constrained gene implicated in pontocerebellar hypoplasia (Karczewski et al., 2020; Wojcik et al., 2018), and *SMYD3*, which is a histone methyltransferase with confirmed oncogenic roles for several types of cancer (Mazur et al., 2014). (**D**) We discovered a 320kb segment on chromosome 1 where rare deletions were associated with neurodevelopmental disorders. This segment overlapped just three genes, none of which have known genotype-phenotype correlations in disease. However, *IER5* is a promising candidate gene for neurodevelopmental disorders, as it is likely haploinsufficient (zero observed truncating variants and pLI=0.89 in gnomAD v2.1) (Karczewski et al., 2020) and is expressed highest in the brain during the early stages of human fetal neurodevelopment (Miller et al., 2014).

Nominally significant large segments from literature

| Cytoband | Nominal rCNVs | Size | NAHR | Known GDs | All Genes | Genes Constrained Against Truncating SNVs | All OMIM Disease Genes |
|---|---|---|---|---|---|---|---|
| 1p36.21–p36.31 | Recip. | 6,003 kb | No | Recip. | 166 genes | 25 genes | 39 genes |
| 1q21.1 | DEL | 362 kb | Yes | DEL | 15 genes | *ANKRD34A, PIAS3* | *PEX11B, RBM8A* |
| 1q21.1–q21.2 | Recip. | 1,062 kb | Yes | Recip. | 10 genes | No genes | *GJA5, GJA8* |
| 2p25.3 | DUP | 471 kb | No | DUP | *MYT1L* | *MYT1L* | *MYT1L* |
| 2p24.3 | DUP | 477 kb | No | DUP | *DDX1, MYCN* | *DDX1* | *MYCN* |
| 2p15 | DEL | 660 kb | No | DEL | 4 genes | *CCT4* | *FAM161A* |
| 2q11.1–q11.2 | Recip. | 932 kb | Yes | Recip. | 18 genes | 4 genes | 7 genes |
| 2q13 | DEL | 1,713 kb | Yes | Recip. | 9 genes | *ANAPC1* | *ANAPC1, BUB1, MERTK* |
| 2q21.1 | DEL | 700 kb | Yes | Recip. | 7 genes | No genes | No genes |
| 2q23.1 | DEL | 570 kb | No | DEL | *MBD5, ORC4* | *MBD5* | *MBD5, ORC4* |
| 2q32.3–q33.3 | DEL | 8,282 kb | No | DEL | 63 genes | 13 genes | 16 genes |
| 2q37.3 | Recip. | 1,194 kb | No | Recip. | 45 genes | 6 genes | 9 genes |
| 3q29 | DUP | 1,589 kb | Yes | Recip. | 22 genes | 5 genes | 6 genes |
| 4p16.3 | Recip. | 826 kb | No | Recip. | 30 genes | *CTBP1, PCGF3, WHSC1* | 12 genes |
| 5q35.2–q35.3 | Recip. | 1,319 kb | Yes | Recip. | 36 genes | 6 genes | 6 genes |
| 6p25.1–p25.3 | Recip. | 5,853 kb | No | Recip. | 35 genes | 6 genes | 8 genes |
| 7q11.23 | DUP | 1,398 kb | Yes | Recip. | 23 genes | 7 genes | 10 genes |
| 7q36.3 | DUP | 221 kb | No | Recip. | *VIPR2, WDR60* | No genes | *WDR60* |
| 8p23.1 | Recip. | 3,666 kb | Yes | Recip. | 26 genes | *XKR6* | *BLK, CTSB, GATA4, RP1L1* |
| 8q21.11 | DEL | 540 kb | No | DEL | *ZFHX4* | *ZFHX4* | *ZFHX4* |
| 9q34.3 | Recip. | 1,240 kb | No | Recip. | 89 genes | 13 genes | 17 genes |
| 10q11.22–q11.23 | Recip. | 3,739 kb | Yes | Recip. | 34 genes | *MAPK8, WDFY4* | 5 genes |
| 10q22.3–q23.2 | Recip. | 6,970 kb | Yes | Recip. | 28 genes | 4 genes | 6 genes |
| 11p11.2 | DEL | 2,060 kb | No | DEL | 18 genes | *MAPK8IP1, PHF21A, PRDM11* | 6 genes |
| 11q13.2–q13.4 | DEL | 3,473 kb | Yes | DEL | 30 genes | 5 genes | 10 genes |
| 12q14.2–q15 | DEL | 3,574 kb | No | DEL | 18 genes | 5 genes | 6 genes |
| 13q12.12 | DEL | 1,485 kb | Yes | DEL | 9 genes | No genes | *MIPEP, SACS, SGCG* |
| 14q32.2–q32.31 | DEL | 1,110 kb | No | DEL | 13 genes | *BEGAIN, EML1, EVL, YY1* | *EML1, YY1* |
| 15q13.1 | Recip. | 845 kb | Yes | Recip. | 4 genes | *TJP1* | No genes |
| 15q24.1 | Recip. | 1,616 kb | Yes | Recip. | 64 genes | 7 genes | 13 genes |
| 15q25.2 | Recip. | 1,501 kb | Yes | Recip. | 13 genes | 4 genes | *AP3B2, HOMER2* |
| 15q25.2–q25.3 | DEL | 729 kb | Yes | DEL | 8 genes | *ZNF592* | *ALPK3, WDR73, ZNF592* |
| 15q26.3 | DUP | 3,163 kb | No | DUP | 22 genes | 4 genes | 6 genes |
| 16p13.11 | Recip. | 1,156 kb | Yes | Recip. | 11 genes | *KIAA0430, MYH11* | *ABCC6, MYH11, NDE1* |
| 16p12.3 | Recip. | 1,311 kb | Yes | Recip. | *XYLT1* | *XYLT1* | *XYLT1* |
| 16p11.2–p12.2 | DEL | 7,098 kb | Yes | Recip. | 64 genes | 12 genes | 15 genes |
| 17p13.3 | Recip. | 1,230 kb | No | Recip. | 41 genes | 10 genes | 13 genes |
| 17p11.2–p12 | Recip. | 1,114 kb | Yes | Recip. | 23 genes | *NCOR1, PMP22* | 5 genes |
| 17p11.2 | Recip. | 3,403 kb | Yes | Recip. | 52 genes | 5 genes | 11 genes |
| 17q11.2 | Recip. | 1,156 kb | Yes | Recip. | 13 genes | 4 genes | *NF1, RNF135* |
| 17q21.31 | DEL | 460 kb | Yes | Recip. | 5 genes | *KANSL1* | *KANSL1, MAPT* |
| 17q23.1–q23.2 | DEL | 2,133 kb | Yes | Recip. | 15 genes | 5 genes | 5 genes |
| 19p13.11–p13.2 | DEL | 3,620 kb | No | DEL | 77 genes | 13 genes | 12 genes |
| 21q21.3 | DUP | 291 kb | No | DUP | *APP* | No genes | *APP* |
| 22q11.1–q11.21 | DUP | 1,199 kb | Yes | DUP | 13 genes | *CECR2, MICAL3, PEX26* | *ATP6V1E1, IL17RA, PEX26* |
| 22q11.21–q11.23 | Recip. | 1,732 kb | Yes | Recip. | 18 genes | *BCR, GNAZ, MAPK1* | *BCR, MAPK1* |
| 22q13.2–q13.33 | DUP | 8,163 kb | No | Recip. | 88 genes | 13 genes | 14 genes |

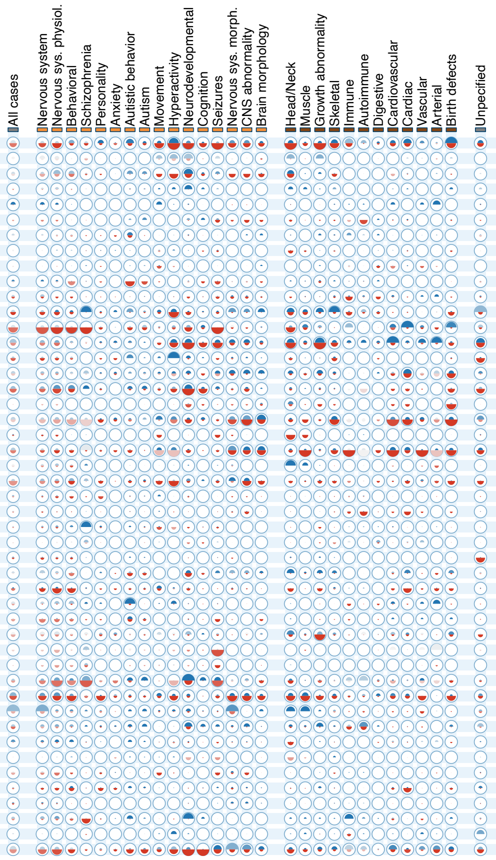

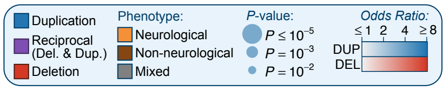

**Figure S7 | Disease-associated rCNV segments curated from existing literature.** In addition to the 49 rCNV segments associated with disease at genome-wide significance (**Figure 2**), we also analyzed 79 segments reported by at least one of six sources (Dittwald et al., 2013; Firth et al., 2009; Girirajan et al., 2012; Owen et al., 2018; Riggs et al., 2012; Stefansson et al., 2014) that did not reach our likely conservative criteria for genome-wide significance ($P=3.72x10^{-6}$) but were at least nominally ($P<0.05$) associated with one or more phenotypes in our study. Details for each of these 79 rCNV segments are summarized here. Overlapping segments—including "reciprocal" loci like 1q21 or 17p11.2—have been collapsed into single rows for clarity. For each locus, we provide segment size, predicted NAHR-mediated mechanism, meta-analysis summary statistics for each of the 30 phenotypes evaluated, and genic content, further partitioned by constraint against truncating point mutations (Karczewski et al., 2020) and previously reported associations with any disease phenotype (Amberger et al., 2015). Summary statistics are represented as one semicircle each for duplication (blue) and deletion (red) shaded by the effect size estimate with radii scaled according to the final meta-analysis $-\log_{10}$ P-value.

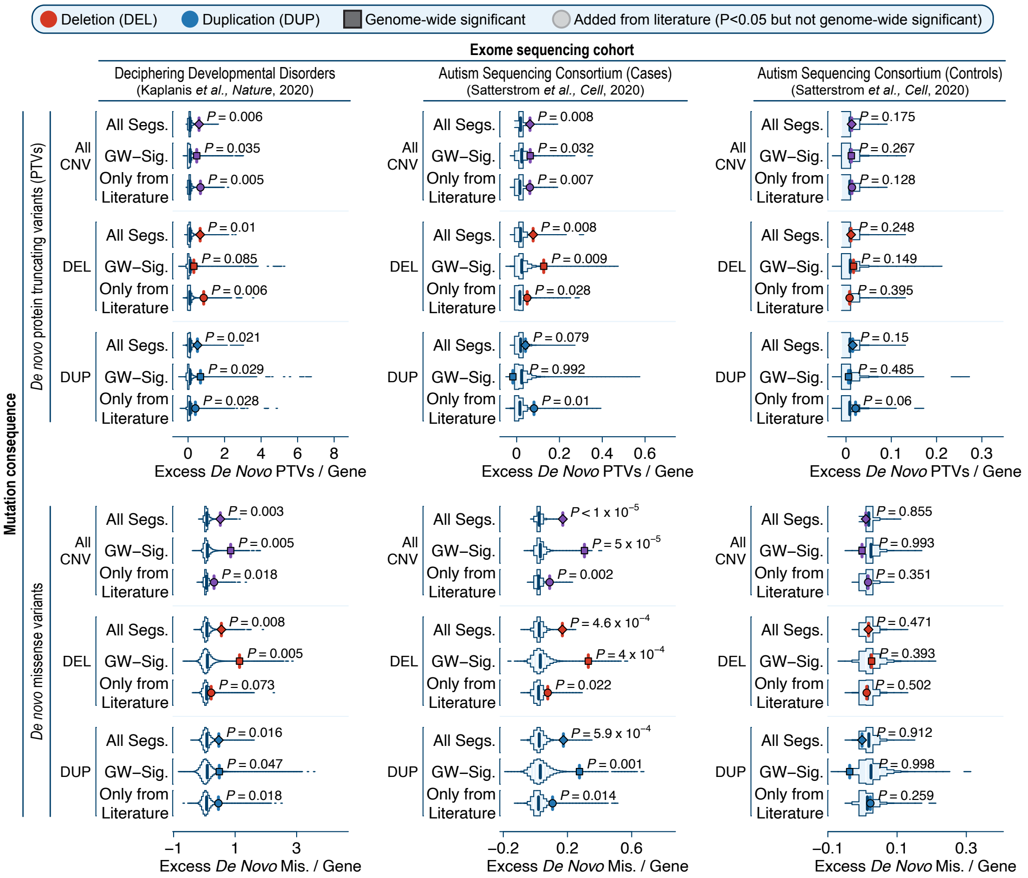

**Figure S8 | Permutation tests of *de novo* coding mutations in large rCNV segments.** The genes within disease-associated rCNV segments were significantly enriched for excesses of damaging *de novo* mutations (DNMs) identified in two independent exome-sequencing studies of developmental disorders (Kaplanis et al., 2020) and autism (Satterstrom et al., 2020). Here, we provide the results from 100,000-fold permutation tests matched on the number of genes per segment for three cohorts (horizontal axis) and two mutational consequences (protein-truncating variants [PTVs] and missense variants; vertical axis). Counts of *de novo* variants were residualized against gene-specific mutation rates of each mutational consequence (PTV or missense) (Karczewski et al., 2020), and subsequently normalized against the rate of synonymous *de novo* mutations to control for any localized mutation rate miscalibration.

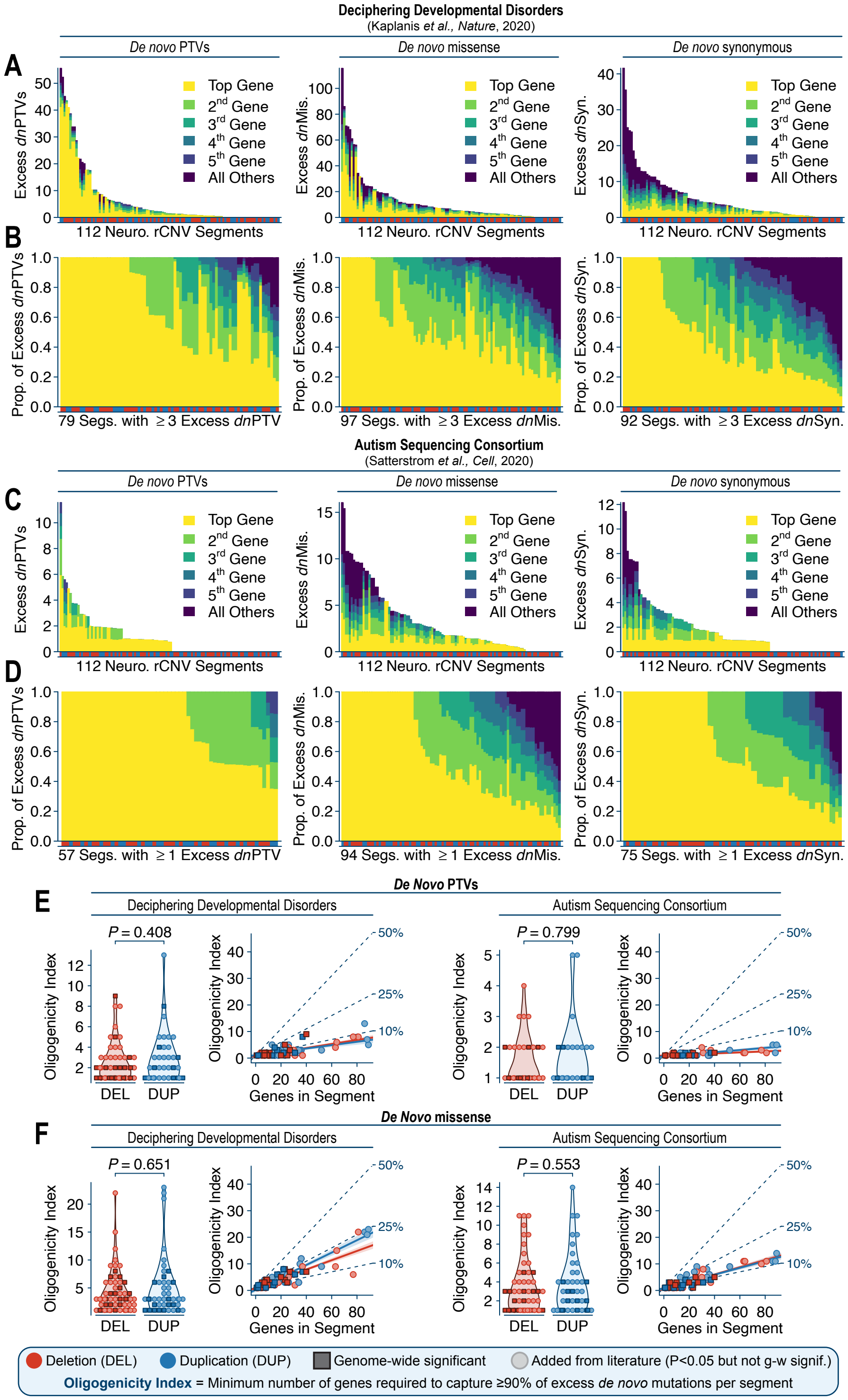

**Figure S9 | Distributions of *de novo* coding mutations within large rCNV segments.** We evaluated the distributions of *de novo* point mutations (DNMs) from two exome sequencing studies of developmental disorders (Kaplanis et al., 2020) and autism (Satterstrom et al., 2020) to better understand the genetic architecture of 128 disease-associated large rCNV segments. (**A**) Distributions of total excess DNMs per segment from a study of 31,058 children with developmental disorders (Kaplanis et al., 2020). Genes within each segment associated with one or more neurological phenotypes (n=112 rCNV segments) were ranked based on their mutation rate-adjusted (Karczewski et al., 2020) excess of DNMs per consequence (protein-truncating variants [PTVs], missense [Mis.], and synonymous [Syn.]) before being tallied in order. The five genes with greatest excess DNMs per consequence were assigned unique colors; excess DNMs in all genes after the fifth gene were summed and assigned the darkest color (purple; labeled "All Others"). Red and blue tabs beneath each segment indicate deletion- and duplication-associated segments, respectively. (**B**) Cumulative distributions of *excess* DNMs per rCNV segment; *i.e.*, data from (A) divided by the total number of *excess* DNMs per segment. (**C**-**D**) Distributions of DNMs per rCNV segment from a study of 5,924 children with autism (Satterstrom et al., 2020). Data are represented identically as in (A-B). (**E**) We compared the distributions of excess *de novo* PTVs from the exome sequencing studies in (A-D) to estimate the number of dosage sensitive genes within each segment, defined here as the minimum number of genes that captures ≥90% of the total excess *de novo* PTVs per segment ("oligogenicity index"). Most excesses of *de novo* PTVs per rCNV segment could be captured by just one or two genes (left panels), although this statistic was proportional to segment size (right panels). There were no significant differences between deletions and duplications (two-sided Wilcoxon tests). Trendlines are outlier-robust linear fits. Dashed lines correspond to expected oligogenicity index values if ≥90% of the total excess *de novo* PTVs could be explained by 50%, 25%, or 10% of all genes per segment on average, respectively. (**F**) Comparisons of excess *de novo* missense mutations per rCNV segment from the studies in (A-B) and (C-D), formatted identically as in (E).

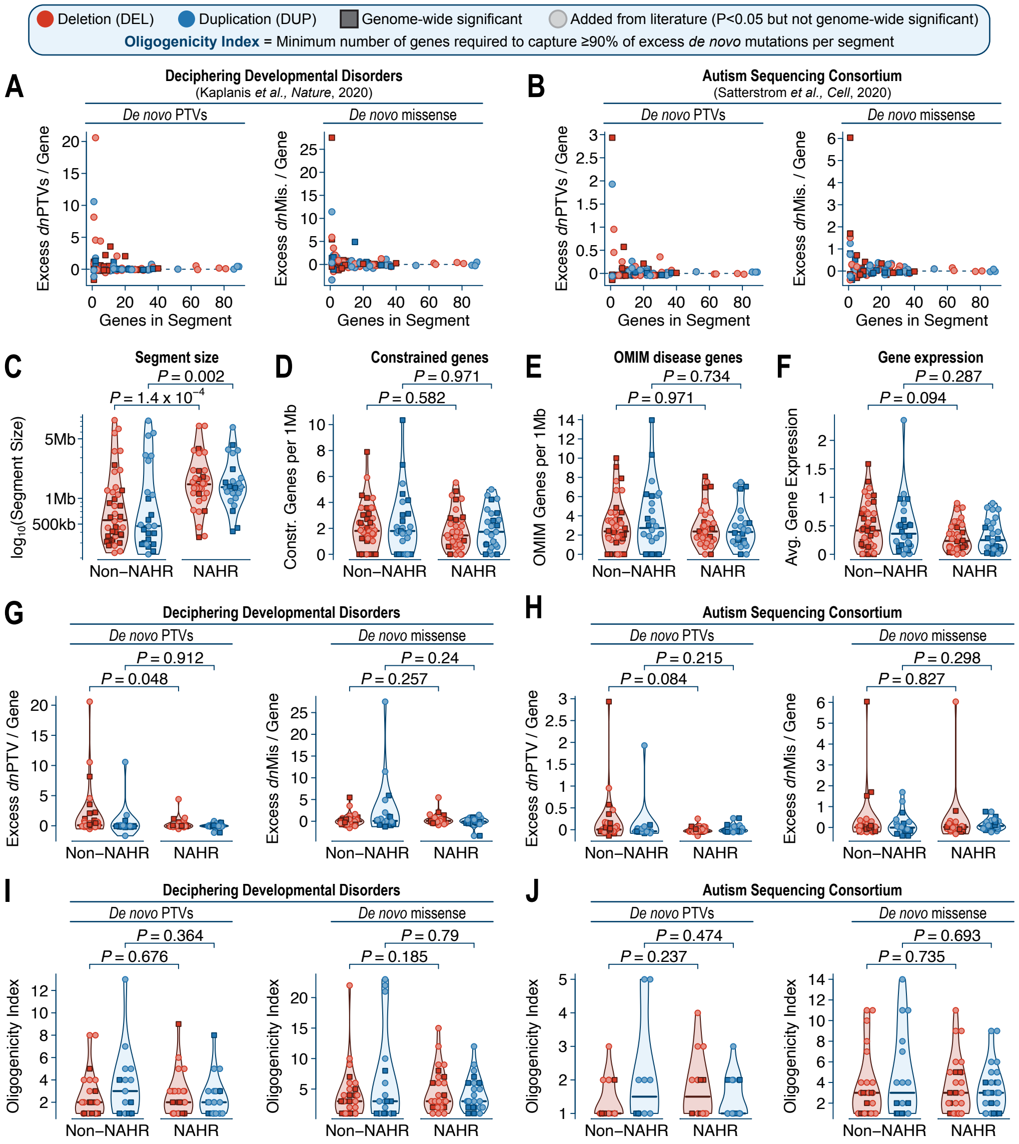

**Figure S10 | Comparisons of NAHR-mediated and non-NAHR-mediated rCNV segments.** (**A-B**) Smaller rCNV segments had greater excesses of damaging *de novo* mutations (DNMs) in large-scale exome sequencing studies of developmental disorders (Kaplanis et al., 2020) and autism (Satterstrom et al., 2020). Excess DNMs were calculated based on the number of genes per segment while adjusted for gene-specific mutation rates. (**C-J**) We classified all 128 rCNV segments based on predicted mechanism (NAHR-mediated vs. non-NAHR-mediated breakpoints) and compared the genomic features of these two subgroups. After accounting for the significantly larger average size of NAHR-mediated segments (C), we found no significant differences between NAHR-mediated and other segments for (D) density of mutationally constrained genes (Karczewski et al., 2020), (E) density of known disease genes (Amberger et al., 2015), (F) harmonic mean of RNA expression of the genes within each segment (scaled as $\log_{10}(TPM + 1)$ in GTEx v7) (Battle et al., 2017), or excesses of *de novo* protein-truncating variants (PTVs) or missense mutations in the same (G) developmental disorder and (H) autism cohorts from (A-B). All comparisons were performed as two-sided Wilcoxon tests and P-values are reported here before any adjustment for multiple comparisons. (**I-J**) We examined the distributions of DNMs among the two exome sequencing studies in (A-B) for evidence favoring a single dominant gene by computing the minimum number of genes that capture ≥90% of the total excess *de novo* mutations across all genes per segment (referred to here as the "oligogenicity index"). We found no significant differences in oligogenicity index between NAHR-mediated and other rCNV segments.

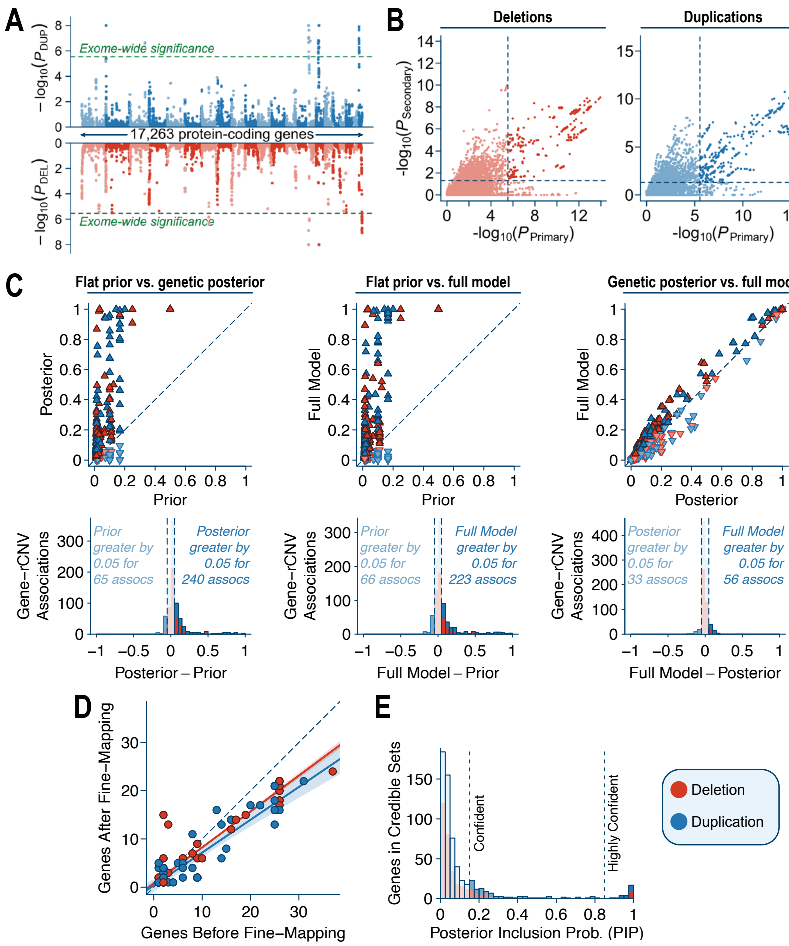

**Figure S11 | Technical details of gene-based association testing and fine-mapping.** (**A**) Example Miami plot of exome-wide rCNV association statistics for one phenotype (neurodevelopmental abnormalities; HP:0012759). (**B**) Relationship of primary and secondary P-values across all meta-analyses. Dashed lines indicate thresholds for exome-wide significance, and dark shaded points indicate genes surpassing both thresholds. (**C**) Comparison of causal probabilities for all exome-wide significant gene-phenotype pairs based on (i) a uniform prior, (ii) fine-mapped posterior after accounting for rCNV evidence alone ("posterior"), and (iii) fine-mapped posterior from the full model including 129 gene-level features ("full model"). Points are colored by CNV type (deletion or duplication) and oriented according to which model produced a higher causal probability (**D**) Comparisons of total number of genes per association before and after fine-mapping. Solid lines are outlier-robust linear fits per CNV type. (**E**) Distribution of PIPs from our final fine-mapping model, colored by CNV type and shaded by fine-mapping confidence. Horizontal lines correspond to "confident" (PIP≥0.15) and "very confident" (PIP≥0.85) fine-mapping thresholds.

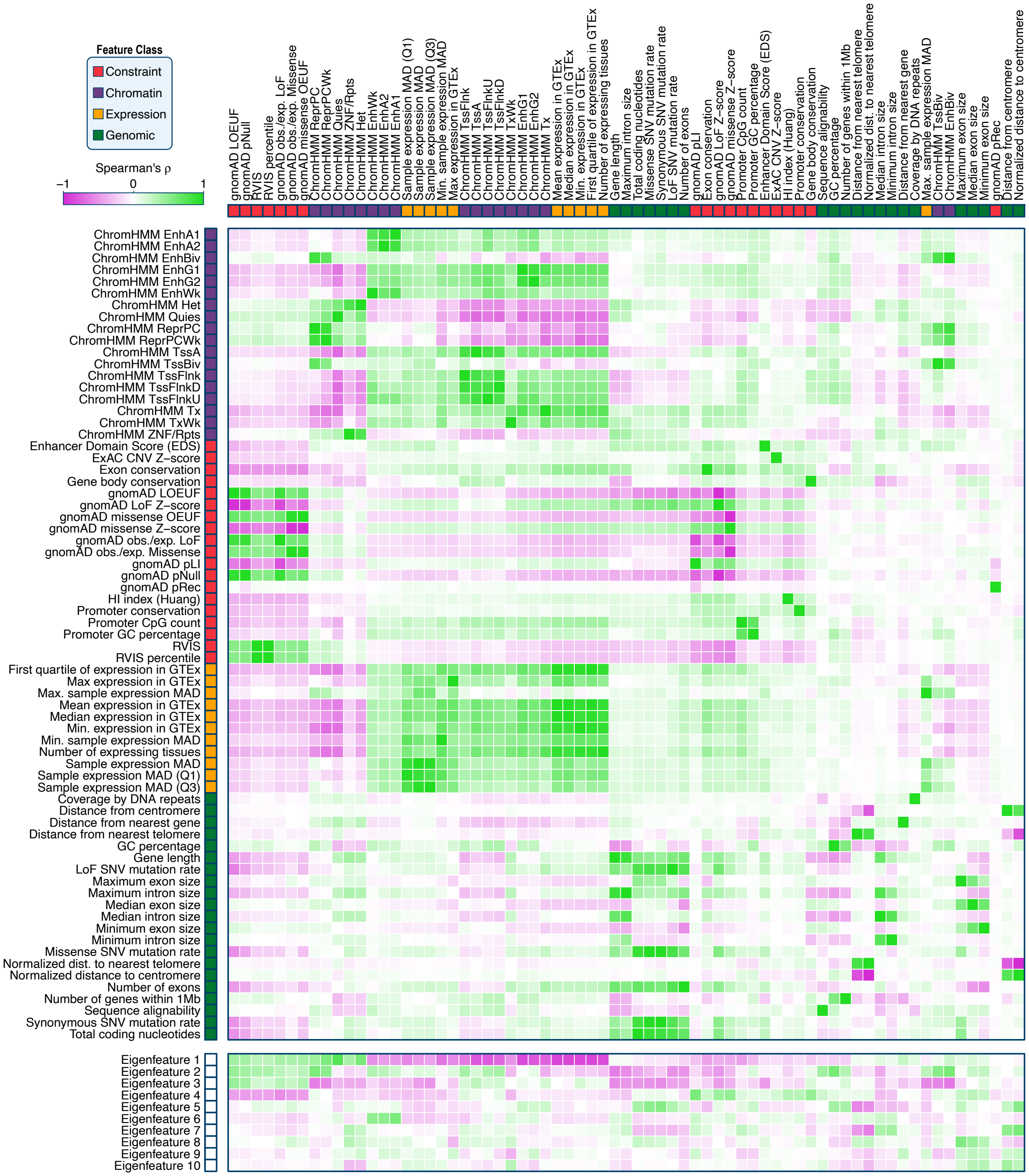

**Figure S12 | Gene-level features used in fine-mapping and dosage sensitivity scoring.** Spearman correlation matrix of 69/129 gene-level features and the top 10 "eigenfeatures." Due to space limitations, 60 derived features are not shown here, including feature standard deviations and principal component-based features.

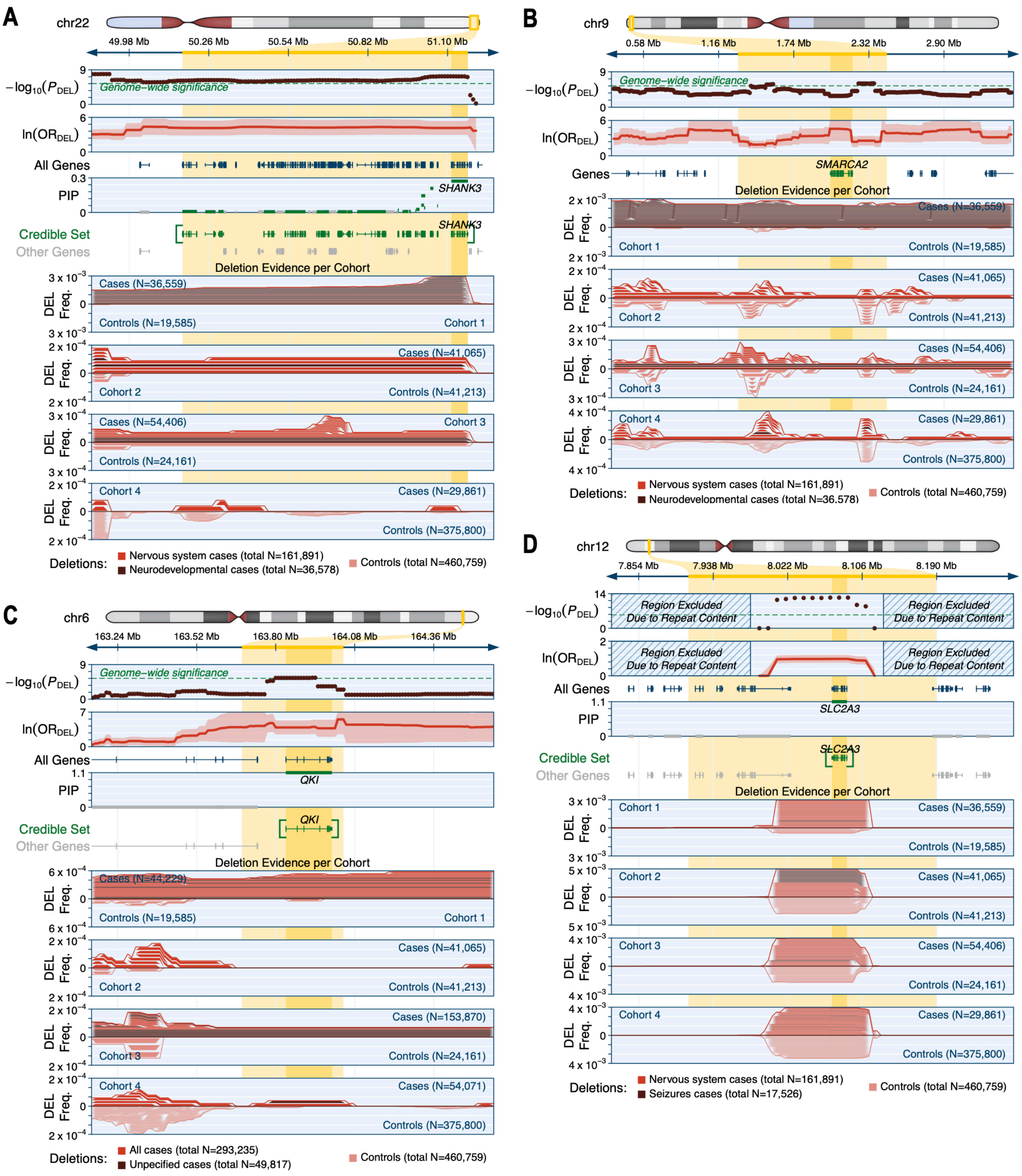

**Figure S13 | Selected genes from rCNV association discovery analyses.** (**A**) We identified a 95% credible set of 19 genes spanning 1Mb on chromosome 22 where rare deletions were associated with neurodevelopmental disorders. Fine-mapping prioritized *SHANK3* as the likely causal gene underlying this association (PIP=0.27), which matches the validated driver gene of 22q terminal deletions in Phelan-McDermid syndrome (Zhou et al., 2019). Meta-analysis P-values and ORs are provided for the more specific (smaller N) of the two phenotypes listed at the bottom of the panel, and ORs are also provided with a 95% confidence interval in lighter shading. (**B**) We identified a 1.2Mb segment of chromosome 9 containing just one gene, *SMARCA2*, where rare deletions were associated with neurodevelopmental disorders. Missense mutations in *SMARCA2* are a known cause of Nicolaides-Baraitser syndrome but the pathogenic mechanism is unknown (Geisheker et al., 2017; Van Houdt et al., 2012). The deletion association we identified here indicates that *SMARCA2* is haploinsufficient, which could help clarify the likely mechanism of the missense mutations observed in Nicolaides-Baraitser patients. (**C**) We discovered a single-gene credible set on chromosome 6 where rare deletions were associated with disease (*i.e.*, in affected cases with unspecified phenotypes). The only gene in this credible set, *QKI* (PIP=1.0), is a mutationally constrained gene with no previously known germline disease associations (Karczewski et al., 2020). We found multiple gene-specific deletions of *QKI* (and no other genes) in affected individuals. (**D**) We identified a credible set of one gene, *SLC2A3*, on chromosome 12 where rare deletions were associated with seizures with a modest effect sizes (OR = 2.7; 95% CI = 2.1-3.3). CNVs of *SLC2A3* have been proposed as risk factors for various neurological phenotypes, including age-of-onset of Huntington's disease and epilepsy (Vittori et al., 2014; Ziegler et al., 2020), and heterozygous *Slc2a3* loss-of-function mice have abnormal neuronal activity patterns (Schmidt et al., 2008). Formatting conventions are identical across all panels.

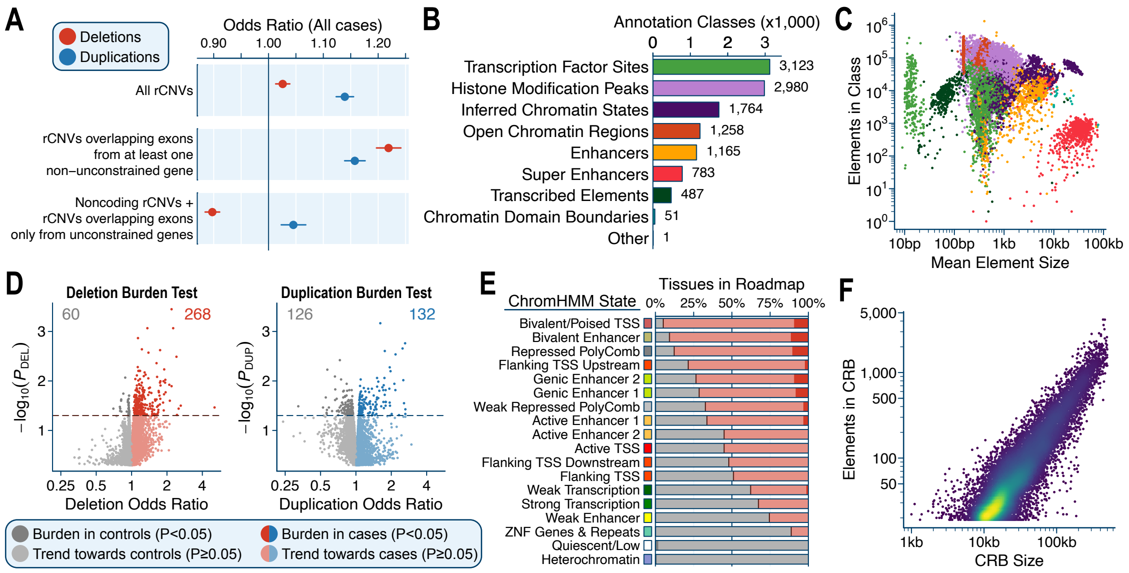

**Figure S14 | Technical details of noncoding annotation prioritization and curation.** (**A**) Fixed-effect meta-analysis odds ratios for rCNVs in all cases vs. controls stratified by genic overlap. "Unconstrained" genes reflect the subset of 4,793 genes meeting criteria for inclusion in noncoding association testing (**Note S2**). Bars are 95% confidence intervals. (**B**) Summary of the 11,612 annotation classes by family considered during the creation of *cis*-regulatory blocks (CRBs) for noncoding rCNV association testing. (**C**) Distribution of mean element size and total number of elements per annotation class from (B). (**D**) Summary statistics of global noncoding rCNV case-control burden testing for all 11,612 annotation classes for deletions (left) and duplications (right). Dashed line indicates a nominally significant (P<0.05) burden of rCNVs in cases (dark red & blue points) or controls (dark grey points). Numbers in top left and right corners indicate total number of annotation classes nominally significant in controls (left) or cases (right). (**E**) To assess whether our global rCNV burden tests were correctly prioritizing annotation classes most likely to be dosage sensitive, we evaluated a matched set of 18 chromatin states across 98 cell types and tissues from the Roadmap Epigenomics Project (Kundaje et al., 2015). For each chromatin state, we computed the proportion of tissues with a burden of rare deletions in cases (red) or controls (grey). Encouragingly, we found that the states most enriched for rare deletions in cases were generally activating elements, like enhancers, whereas the states most enriched for rare deletions in controls were inactive or silent chromatin. Bar colors are consistent with the legend from (D). (**F**) Relationship of CRB size and number of elements per CRB, colored by density.

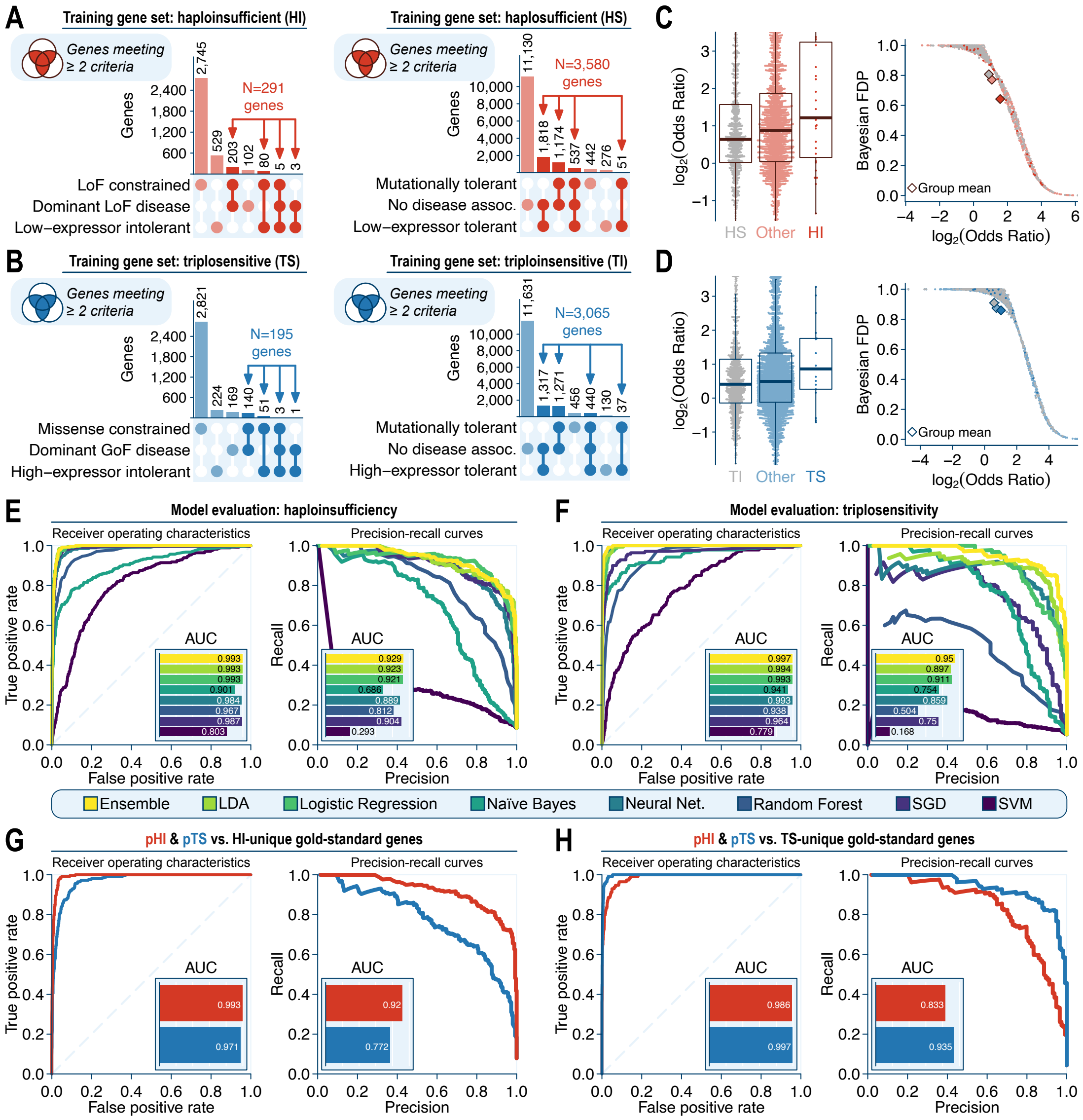

**Figure S15 | Genic dosage sensitivity model development.** (**A-B**) We defined training sets of genes that were likely (A) haploinsufficient & haplosufficient and (B) triplosensitive & triploinsensitive as all genes meeting at least two of the three criteria listed per category. (**C-D**), we computed case:control odds ratios using the same meta-analysis approach as for our gene-based association analyses, with minor modifications for optimized parameters (**Note S3**). We subsequently transformed these odds ratios into Bayesian false discovery probabilities (BFDPs), which reflected the odds that each gene was dosage sensitive based on our model's priors. These BFDPs were used to train a machine learning model to predict dosage sensitivity for every gene. (**E-F**) For our final scores, we used an ensemble averaging model (yellow), which was the most performant as determined by the harmonic mean of area under the curve (AUC) statistics for receiver operating characteristic and precision-recall curves evaluated against the curated likely dosage sensitive gene sets from (A-B) for (E) haploinsufficiency and (F) triplosensitivity. (**G-H**) We assessed the specificity of our final scores, pHI and pTS, against the non-overlapping subsets of our curated likely dosage sensitive training genes that were (G) only haploinsufficient or (H) only triplosensitive, finding that pHI outperformed pTS when predicting haploinsufficiency and pTS outperformed pHI when predicting triplosensitivity.

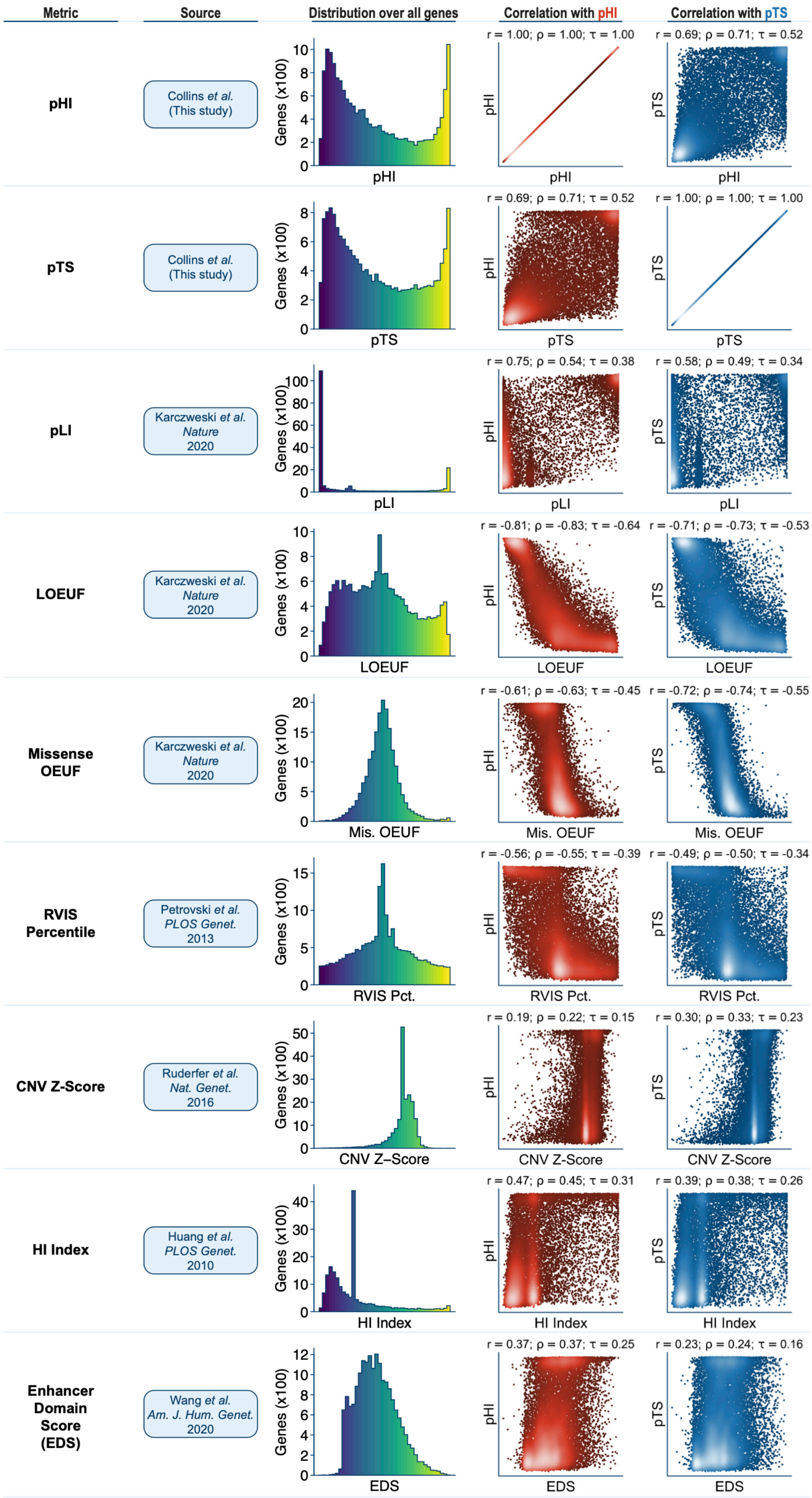
Metric
Source
Distribution over all genes
Correlation with pHI
Correlation with pTS
pHI
Collins et al. (This study)
r = 1.00; ρ = 1.00; τ = 1.00
r = 0.69; ρ = 0.71; τ = 0.52
pTS
Collins et al. (This study)
r = 0.69; ρ = 0.71; τ = 0.52
r = 1.00; ρ = 1.00; τ = 1.00
pLI
Karczweski et al. Nature 2020
r = 0.75; ρ = 0.54; τ = 0.38
r = 0.58; ρ = 0.49; τ = 0.34
LOEUF
Karczweski et al. Nature 2020
r = -0.81; ρ = -0.83; τ = -0.64
r = -0.71; ρ = -0.73; τ = -0.53
Missense OEUF
Karczweski et al. Nature 2020
r = -0.61; ρ = -0.63; τ = -0.45
r = -0.72; ρ = -0.74; τ = -0.55
Mis. OEUF
RVIS Percentile
Petrovski et al. PLOS Genet. 2013
r = -0.56; ρ = -0.55; τ = -0.39
r = -0.49; ρ = -0.50; τ = -0.34
RVIS Pct.
CNV Z-Score
Ruderfer et al. Nat. Genet. 2016
r = 0.19; ρ = 0.22; τ = 0.15
r = 0.30; ρ = 0.33; τ = 0.23
HI Index
Huang et al. PLOS Genet. 2010
r = 0.47; ρ = 0.45; τ = 0.31
r = 0.39; ρ = 0.38; τ = 0.26
Enhancer Domain Score (EDS)
Wang et al. Am. J. Hum. Genet. 2020
r = 0.37; ρ = 0.37; τ = 0.25
r = 0.23; ρ = 0.24; τ = 0.16
EDS
Genes (x100)

**Figure S16 | Comparison of genic dosage sensitivity scores to existing metrics.** Comparisons of the probabilities of haploinsufficiency (pHI) and triplosensitivity (pTS) against seven existing gene-level constraint metrics (Huang et al., 2010; Karczewski et al., 2020; Petrovski et al., 2013; Ruderfer et al., 2016; Wang and Goldstein, 2020). For each metric, we provide the distribution of scores for the 17,263 autosomal protein-coding genes we considered when developing pHI and pTS. Genes lacking metrics for each original source were assigned the per-chromosome mean for that metric. We also provide scatterplot comparisons of each metric against pHI (red) and pTS (blue). Pearson's r, Spearman's rho, and Kendall's tau are provided as measures of correlation for each comparison.

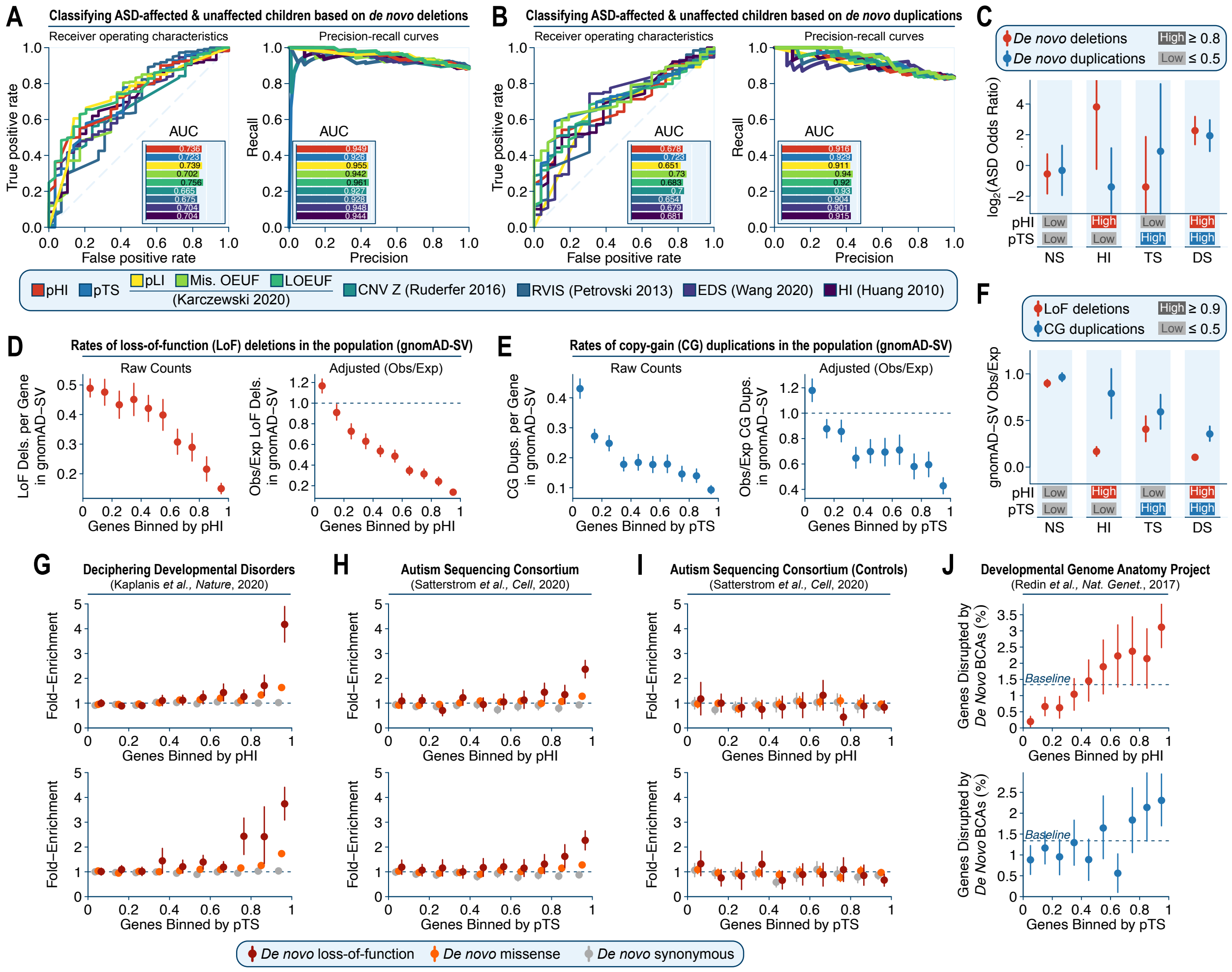

**Figure S17 | Quality assessment of genic dosage sensitivity scores.** (**A-B**) We evaluated our novel dosage sensitivity scores, pHI and pTS, against seven existing gene-level intolerance metrics (Huang et al., 2010; Karczewski et al., 2020; Petrovski et al., 2013; Ruderfer et al., 2016; Wang and Goldstein, 2020) when distinguishing children affected with autism spectrum disorder (ASD) from their unaffected siblings based on the genes overlapped by their *de novo* CNVs alone (**Note S4**) (Satterstrom et al., 2020). For this analysis, we restricted to the 426 children carrying at least one non-GD *de novo* protein-truncating deletion or whole-gene duplication. (A) For *de novo* deletions, pHI outperformed all metrics other than pLI and LOEUF from gnomAD based on the harmonic mean of the area under the curve (AUC) statistics for receiver operating characteristic and precision-recall curves. (B) For *de novo* duplications, pTS similarly outperformed all metrics other than missense OEUF from gnomAD. (**C**) pHI and pTS stratified risk for ASD conferred by *de novo* CNVs preferentially by CNV type (*i.e.*, pHI was more effective for deletions and pTS was more effective for duplications). (**D**) Left: average counts of protein-truncating deletions in gnomAD-SV v2.1 (Collins et al., 2020) for all genes binned by pHI. Bars represent 95% confidence intervals after 1,000-fold bootstrapping. Right: same data from left panel after adjustment for per-gene expected number of LoF deletions using a negative binomial regression as previously described (Collins et al., 2020). (**E**) Comparisons of whole-gene copy-gain duplications from gnomAD-SV v2.1 versus pTS, presented identically to (D). (**F**) pHI and pTS identified genes depleted for LoF deletions and CG duplications in gnomAD-SV v2.1 preferentially by CNV type, as in (C). (**G-H**) We evaluated the relationship of pHI and pTS versus enrichments of coding *de novo* mutations (DNMs) from two exome sequencing studies of developmental disorders (Kaplanis et al., 2020) and autism (Satterstrom et al., 2020). Genes were assigned to ten fixed-width bins based on pHI or pTS score, and the ratio of total observed DNMs to expected DNMs was computed per gene while adjusting for gene-specific mutation rates (Karczewski et al., 2020). We observed enrichments of loss-of-function and missense DNMs in affected children (G-H) but not in their unaffected siblings (I). Synonymous DNMs showed no enrichment in any comparison, as expected. (**J**) Genes with higher pHI and pTS scores were more likely to have been disrupted by *de novo* balanced chromosomal abnormalities (*i.e.*, translocations & large inversions) among 273 individuals with congenital anomalies (Redin et al., 2016). In all panels, error bars correspond to 95% confidence intervals based on 1,000-fold bootstrapping.

### SUPPLEMENTAL TABLES

*Note: all Supplemental tables are provided in a separate file.*

**Table S1 | Data sources, sample counts, and cohort metadata.**
Information on the cohorts included in this study.

**Table S2 | Standardized phenotypes and sample sizes.**
List of HPO terms and sample sizes used in this study.

**Table S3 | Locus-level summary statistics for genome-wide significant large segments.**
Meta-analysis summary statistics for the 49 rCNV segments associated with one or more phenotypes at genome-wide significance. Abbreviations: "freq." = rCNV carrier frequency as an inverse variance-weighted average across all four cohorts; "ln_or" = natural log-odds ratio of association in cases.

**Table S4 | Association-level summary statistics for genome-wide significant large segments.**
Meta-analysis summary statistics for each rCNV-phenotype association corresponding to one of the 49 rCNV segments in **Table S3**. Abbreviations are the same as in **Table S3**.

**Table S5 | Gene set definitions.**
List of gene sets used in this study and their curation criteria.

**Table S6 | Master table of all large rCNV segments considered in analyses.**
Summary of nonredundant set of 128 large rCNV segments derived from our genome-wide discovery analysis and literature curation. Abbreviations: "meta_best_p" = most significant meta-analysis P-value observed within the segment for any phenotype; "meta_best_ln_or" = strongest meta-analysis natural log-odds ratio observed within the segment for any phenotype (which may not necessarily match the phenotype from "meta_best_p"); "gw_sig" = segment met criteria for genome-wide significance in our discovery analysis; "literature_gd" = segment matched a GD previously reported in one of six sources curated from the literature; "nahr" = predicted non-allelic homologous recombination-based CNV mechanism. Gene sets are defined in **Table S5**.

**Table S7 | Gene-level features used for fine-mapping and dosage sensitivity scoring.**
Details for all gene-level features considered during fine-mapping and dosage sensitivity scoring.

**Table S8 | Summary statistics for exome-wide significant credible gene sets.**
Meta-analysis summary statistics for the 85 exome-wide significant gene-based credible sets. Abbreviations are the same as in **Table S3**. Additional abbreviations: "top_gene" = gene with best posterior inclusion probability (PIP) among all genes in the credible set; "vconf_genes" = list of genes with PIP ≥ 0.85; "conf_genes" = list of genes with 0.85 > PIP ≥ 0.15.

**Table S9 | Confident fine-mapped gene-phenotype rCNV associations.**
Meta-analysis summary statistics for all individual gene-rCNV-phenotype associations detected at exome-wide significance and fine-mapped to a posterior inclusion probability (PIP) ≥ 0.15. Abbreviations are the same as in **Table S3**.

**Table S10 | Sources for genome annotations used in noncoding association tests.**

List of genome annotation sources curated in this study, including a description of which annotations were considered per source.

**Table S11 | Summary statistics from rCNV burden tests of genome annotation classes.**
List of all genome annotation tracks evaluated for inclusion in *cis*-reguatory block (CRB) creation, including their summary statistics from global rCNV burden testing in cases vs. controls.

**Table S12 | Haploinsufficiency and triplosensitivity scores for autosomal protein-coding genes.**
List of probability of haploinsufficiency (pHI) and triplosensitivity (pTS) scores for 17,263 genes.

### SUPPLEMENTAL NOTES

#### Note S1 | Determination of genome-wide significance for sliding window meta-analyses.

Conventional genome-wide association studies (GWAS) of common short variants typically use $P < 5 \times 10^{-8}$ as the threshold for genome-wide significance, which corresponds to a Bonferroni adjustment for all common short variants in the human population (Roeder and Wasserman, 2009). This threshold has been rigorously vetted and widely adopted by the human genetics community. However, there has been less investigation of how appropriate this threshold is for association studies of rare variants (Xu et al., 2014), and extremely little discussion for the extension of this threshold for other classes of variants like CNVs. Thus, we evaluated genome-wide significance in our rCNVs sliding window analyses as the Bonferroni-corrected P-value threshold corresponding to the number of independent, non-overlapping 200kb windows, which we calculated by merging all overlapping windows and dividing the sum of their sizes by 200kb (effective N=13,457.4 independent windows; $P=3.72 \times 10^{-6}$). We subsequently assessed the calibration of this P-value threshold via phenotype permutation. For each permutation, we shuffled all sample phenotype labels per cohort while matching on CNV size between cases and controls and reran the full meta-analysis model across all cohorts from this permuted dataset. We conducted 50 independent permutations for all phenotypes (N=30) and CNV classes (N=2), for a total of 3,000 permuted association meta-analyses. Finally, for each phenotype, we computed the median maximum P-value observed across all permutations as an estimate of true genome-wide significance given the properties of our dataset. The average maximum P-value observed for most phenotypes was less significant than our Bonferroni-corrected threshold of $P=3.72 \times 10^{-6}$ (**Figure S4D-E**), indicating that the Bonferroni threshold was appropriate for our data, association model, and study design.

#### Note S2 | Depleting rCNVs for strong effects from direct protein-coding alterations.

We wanted to identify loci where rCNVs were associated with disease through mechanisms unlikely to be explained by direct alterations to protein-coding sequences. Specifically, we were interested in identifying rCNV-associated *cis*-regulatory blocks (CRBs) of non-protein-coding functional elements. We accomplished this by filtering our rCNV data to deplete for predicted coding effects. The most conservative approach would be to exclude any rCNV on the basis of any overlap with any annotated exon; however, this approach was inappropriately strict given our rapidly increasing understanding that *cis*-regulation of gene expression is frequently mediated by sequences that lie within or near other, unrelated protein-coding genes, such as the widespread observation of gene promoters as strong enhancers of other genes in *cis* (Andersson and Sandelin, 2020; Fulco et al., 2019; Jung et al., 2019) or the enrichment of transcribed genes near chromatin domain boundaries (van Steensel and Furlong, 2019). Instead, we aimed to deplete our rCNV dataset for all direct genic effects *except* for those deemed unlikely to make large contributions to disease risk due to rare genetic variation. To accomplish this, we defined a set of 4,793 genes—approximately one-quarter of all autosomal genes—we determined to be unlikely to mediate strong effects in disease due to rare coding alterations. These “likely unconstrained” genes were defined as the intersection of two criteria. First, we required these genes to have no previously reported disease association in OMIM (Amberger et al., 2015). Second, we required these genes to have no population-level evidence for strong selection against coding variants; specifically, genes were required to meet all of the following metrics in gnomAD v2.1: LOEUF ≥ 1, missense OEUF ≥ 1, synonymous Z-score ≥ -3 and ≤ 3, pLI ≤ 0.1, LoF O/E in the upper 50% of all genes, missense O/E in the upper 50% of all genes, observed LoF variants > 0, and observed missense variants > 0 (Karczewski et al., 2020). We excluded all rCNVs overlapping an annotated exon from any gene except for the 4,793 “likely unconstrained” genes defined here for all subsequent CRB association testing. While this approach was imperfect and likely failed to exclude all coding effects relevant to disease or cryptic unannotated exons, we observed that the global burden of rCNVs in cases vs. controls was almost entirely captured by the ~75% of genes not in this set of “likely unconstrained” genes, whereas there was a vastly attenuated burden in cases vs. controls after excluding rCNVs overlapping genes not in this subset (**Figure S14A**).

##### Note S3 | Parameter optimizations for genic dosage sensitivity modeling.

The first step of our genic dosage sensitivity scoring model used rCNV effect sizes per gene as inputs. We estimated effect sizes using a fixed-effects meta-analysis similar to our previous gene-based association testing, as detailed in *Methods*; however, for the purposes of dosage sensitivity scoring, we modified four parameters to enrich for focal strong effects attributable to individual genes. First, we standardized the coding sequence (CDS) overlap requirements across deletions and duplications by requiring all rCNVs to overlap ≥80% of the total CDS per gene before qualifying as a likely gene-disruptive rCNV. Second, we excluded uninformative massive rCNVs by optimizing the maximum permissible number of genes disrupted per rCNV (≤24). We accomplished this by maximizing the difference between cases and controls of rCNVs disrupting a manually curated set of likely dosage sensitive gene as a fraction of all disrupted manually curated likely dosage sensitive and insensitive genes. Third, we identified a subset of phenotypes (N=9) with the strongest enrichments for coding rCNVs by computing the meta-analysis effect size of whole-gene rCNVs over curated likely dosage sensitive genes and retaining only phenotypes with <200,000 total cases that were also nominally significantly enriched (P≤0.05) for both deletions and duplications; this excluded many phenotypes with lower prior likelihoods of strong risk contributions from rCNVs (*e.g.*, immune system or cardiovascular abnormalities). Fourth, we optimized the minimum number of total rCNV disruptions per gene (≥10) required for genes to be included for model training by identifying the threshold at which the average standard error of meta-analysis effect size estimates was <1 for both deletions and duplications.

##### Note S4 | *De novo* CNV calls from whole-exome sequencing of autism spectrum disorder families.

As a quality assessment of our genic dosage sensitivity scores, we evaluated enrichments of *de novo* exonic CNVs in 13,363 children affected with autism spectrum disorder versus (ASD) their 5,011 unaffected siblings. These *de novo* CNVs were generated using GATK-gCNV applied to whole-exome sequencing data and represent an extension of the data described in a recent publication (Satterstrom et al., 2020). Manuscripts describing the GATK-gCNV method and the expanded ASD exome sequencing data are forthcoming, but the generation of this dataset is also summarized here. Briefly, we consolidated 72,410 samples—a subset of which comprised families—with exome sequencing data available from the Autism Sequencing Consortium (ASC), the Simons Simplex Collection (SSC), the Simons Powering Autism Research (SPARK), and other general research use cohorts. We performed standard quality control measures as previously described (Satterstrom et al., 2020), and subsequently processed all samples with GATK-gCNV to detect rare coding CNVs. GATK-gCNV is a Bayesian read depth-based CNV detection tool that adjusts for known and unknown technical confounders of exome capture-based sequencing depth, such as GC content and mappability. In a set of 7,165 samples for which we have matching gold-standard genome sequencing-based CNV calls, we found that GATK-gCNV achieved 90% sensitivity and positive predictive value for rare CNVs with site frequency less than 1%. Furthermore, *de novo* CNVs detected by GATK-gCNV achieved >90% sensitivity and >95% positive predictive value for variants spanning more than 3 exons as compared to gold-standard, molecularly validated de novo CNVs identified in matched genome sequencing (Belyeu et al., 2020; Werling et al., 2018). The GATK-gCNV method is publicly available as part of the GATK software package (McKenna et al., 2010), and all of the results and data referenced here pertaining to the ASD samples will be described in detail in a future manuscript.

### SUPPLEMENTAL FILES

**File S1 | Summary statistics from sliding window association meta-analyses.**
Genome-wide sliding window summary statistics for deletion & duplication association meta-analyses for each phenotype are provided in bgzip-compressed BED format in the gzip-compressed tar archive, "sliding_window_sumstats.tar.gz". Statistics suffixed with "secondary" refer to the outcome of the same meta-analysis model run after excluding the most significant individual cohort per window, as described in the *Materials & Methods* section. This file has been temporarily hosted at the following URL until formal publication in a peer reviewed journal: https://storage.googleapis.com/rcnv_project/public/collins_medrxiv_2021/sliding_window_sumstats.tar.gz

**File S2 | Summary statistics from gene-based association meta-analyses.**
Genome-wide gene-based summary statistics for deletion & duplication association meta-analyses for each phenotype are provided in bgzip-compressed BED format in the gzip-compressed tar archive, "gene_based_sumstats.tar.gz". Statistics suffixed with "secondary" refer to the outcome of the same meta-analysis model run after excluding the most significant individual cohort per gene, as described in the *Materials & Methods* section. This file has been temporarily hosted at the following URL until formal publication in a peer reviewed journal: https://storage.googleapis.com/rcnv_project/public/collins_medrxiv_2021/gene_based_sumstats.tar.gz

### METHODS

#### 1 | Data curation

*Cohorts and raw CNV datasets*

In this study we aggregated microarray-based CNV data across diverse sources, ranging from population-scale research biobanks to clinical genetic testing laboratories. The full list of cohorts included in this study is detailed in **Table S1**. We applied cohort-specific quality control measures to each raw CNV dataset prior to CNV harmonization across all cohorts. Below, we describe each cohort and any cohort-specific steps applied prior to CNV harmonization:

- Boston Children's Hospital (BCH): We obtained CNV calls for 3,591 clinically referred samples from the Children's Hospital of Boston as described in a previous study (Talkowski et al., 2012). We subsequently converted the CNVs from hg18 to hg19 coordinates using the UCSC liftOver tool while requiring 50% of each CNV to remap contiguously to hg19 (Lee et al., 2019).

- Coe *et al.* (Coe): We obtained CNV calls for 29,083 samples with developmental disorders and 11,256 unaffected control samples from NCBI dbVar accession #nstd100 as described in a previous publication (Coe et al., 2014).

- Cooper *et al.* (Cooper): We obtained CNV calls for 8,329 unaffected control samples from NCBI dbVar accession #nstd54 as described in a previous publication (Cooper et al., 2011). While affected samples are also described in Cooper *et al.*, we retained only the unaffected controls as all cases from Cooper *et al.* were reprocessed and included in Coe *et al.* above.

- GeneDX (GDX): We obtained CNV calls for 9,958 clinically referred samples from GeneDX. Samples were profiled for CNVs on a custom proprietary SNP array platform. We converted the subset of CNVs provided in hg18 coordinates to hg19 using the UCSC liftOver tool while requiring 50% of each CNV to remap contiguously to hg19 (Lee et al., 2019).

- Indiana University (IU): Chromosomal microarray analysis was performed on genomic DNA extracted from peripheral blood or buccal swab samples from 1,673 consecutive patients tested in the Indiana University Cytogenetics Laboratory during the period of January 2018 through August 2019. These samples were evaluated using the CytoScan HD Microarray platform, which includes both non-polymorphic and SNP oligonucleotide probes, and the data were analyzed via Chromosome Analysis Suite software version 3.3 (ThermoFisher Scientific, USA). All non-artifact copy number variants (CNVs) identified in these samples were retained, irrespective of clinical significance. Following CNV detection, we excluded samples derived from buccal swab DNA, samples with known aneuploidies or large runs of homozygosity, and samples with no phenotypic indication specified. After filtering, we retained a total of 1,576 samples.

- Epi25 Consortium (Epi25k): We obtained CNV calls for 12,758 samples with epilepsy and 8,478 unaffected control samples from the Epi25 Consortium as described in a recent publication (Niestroj et al., 2020). We subsequently restricted CNVs to ≥10 probes and ≥25kb in size. After CNV filtering, samples with >25 CNV calls were excluded as outliers, retaining a total of 12,053 cases and 8,173 controls.

- Psychiatric Genetics Consortium (PGC): We obtained CNV calls for 21,094 samples with schizophrenia and 20,277 unaffected control samples as described in a previous study

(Psychiatric Genetics Consortium, 2016). We subsequently converted the CNVs from hg18 to hg19 coordinates using the UCSC liftOver tool while requiring 50% of each CNV to remap contiguously to hg19 (Lee et al., 2019).

- SickKids Hospital (SickKids): We obtained CNV calls for 2,691 samples with a variety of neurodevelopmental or neuropsychiatric disorders and 1,769 unaffected relatives from NCBI dbVar accession #nstd173 as described in a previous publication (Zarrei et al., 2019). We retained CNVs ≥25kb in size. After CNV filtering, samples with >80 CNV calls were excluded as outliers. We identified a single locus on chr12 that had CNVs only appearing in ADHD samples at 2.8% frequency; these CNVs were removed from the callset. Finally, given that all unaffected samples were first-degree relatives of the affected probands, we excluded all unaffected relatives from subsequent analyses. After filtering, we retained a total of 2,689 samples.

- Simons Simplex Collection (SSC): We obtained CNV calls for 2,795 samples with autism spectrum disorder and their unaffected relatives as described in a previous publication (Sanders et al., 2015). CNVs were filtered on probability (pCNV) $\leq 10^{-9}$, per recommendation of the authors. We subsequently converted the CNVs from hg18 to hg19 coordinates using the UCSC liftOver tool while requiring 50% of each CNV to remap contiguously to hg19 (Lee et al., 2019). Finally, given that all unaffected samples were first-degree relatives of the affected probands, we excluded all unaffected relatives from subsequent analyses.

- The Cancer Genome Atlas (TCGA): We obtained CNV calls for 8,670 whole-blood derived normal (*i.e.*, non-tumor) samples from donors with non-hematological cancers as described in a previous publication (Zack et al., 2013). We subsequently restricted CNVs to ≥10 probes and ≥25kb in size. Finally, to enrich for true germline CNVs and protect against tumor contamination, we further required deletions and duplications to have $\log_2$ intensities $\leq \log_2(0.5)$ and $\geq \log_2(1.5)$, respectively.

- The Genetic Etiology of Tourette Syndrome Consortium (TSAICG): We obtained CNV calls for 2,434 samples with Tourette Syndrome and 4,093 unaffected controls as described in a previous publication (Huang et al., 2017).

- Children's Hospital of Philadelphia (CHOP): We obtained CNV calls for 342,408 samples analyzed by the Center for Applied Genomics at the Children's Hospital of Philadelphia, a subset of which were described in a recent publication (Li et al., 2020). We subsequently restricted CNVs to quality score ≥40, ≥10 probes, and ≥25kb in size. After CNV filtering, samples with LRR_SD <0.25, >20 CNV calls, or SNP call rate <98% were excluded as outliers, as well as samples genotyped on arrays with <175k SNP probes or samples labeled as cancer or Down's Syndrome patients. Finally, we identified 19 loci with platform-specific artifactual CNV pileups. CNVs covered ≥10% by any of these artifact regions were removed from the final CHOP CNV dataset. After filtering, we retained a total of 178,031 samples.

- UK BioBank (UKBB): We obtained CNV calls for 480,051 samples from the UK BioBank, a subset of which was described in a previous publication (Mace et al., 2017). We subsequently restricted CNVs on quality score ≥17 and ≥25kb in size. After CNV filtering, samples with >10 CNV calls were excluded as outliers as well as any samples with known malignant cancers or chromosomal disorders (*e.g.*, Down's Syndrome or sex chromosome aneuploidies), which retained a total of 429,871 samples.

*CNV harmonization*

After collecting and processing CNV calls for each cohort as described above, we next subjected all CNV calls from each cohort to the same set of global filters. These steps are described below, and the code necessary to execute these steps is provided as vignettes in the "filter_CNV_data.sh" script within the GitHub repository associated with this study (see *Code Availability*). In practice, these steps were parallelized on Google Cloud via the Cromwell execution engine, as is implemented in "filter_CNV_data.wdl". We harmonized CNV calls across cohorts as follows:

1. CNV defragmentation: Many microarray-based CNV calling algorithms infrequently introduce fragmentation (*i.e.*, over-segmentation) of large CNV calls. For the purposes of this study, fragmentation of CNV calls has the potential to bias association tests, as single individuals might be counted multiple times for a given locus or gene. Thus, we applied a standardized defragmentation step to raw CNV calls for all studies where this was possible. We defragmented each raw CNV dataset using "defragment_cnvs.py" with --max-dist 0.25, which merges CNVs of the same type (deletion or duplication) found in the same sample if their breakpoints are within ±25% of the size of their corresponding original CNV calls. We applied this process to all cohorts except four (PGC, TSAICG, and control samples from Coe and Cooper), which were unable to be defragmented due to inadequate sample-level metadata.

2. CNV size & ploidy: We excluded all CNVs not mapped to autosomes from the primary hg19 assembly and further required all CNVs to be ≥100kb and ≤20Mb in size.

3. Maximum frequency from population sequencing: We excluded CNVs that matched reported common CNVs (allele frequency >1%) from any annotated population in three independent CNV reference catalogs derived from genome sequencing (Abel et al., 2020; Collins et al., 2020; Sudmant et al., 2015). The version of gnomAD-SV used for this analysis (gnomAD-SV v2.1, non-neuro) included 8,342 samples without known neuropsychiatric disorders as available from the gnomAD website and described in a previous publication (Collins et al., 2020). Matching CNVs were required to have a matching CNV type (deletion or duplication), ≥50% reciprocal overlap by size as calculated by BEDTools (Quinlan and Hall, 2010), and both coordinates from the CNV breakpoint were required to be within ±100kb of the corresponding genome sequencing-based CNV.

4. Maximum frequency in this study: We excluded CNVs that overlapped other CNVs in at least 1% of samples within the same dataset or in any of the other array CNV datasets. For this filter, we compared each cohort's CNV calls to the raw CNVs (before filtering) of each other cohort pairwise in serial and determined matching CNV calls using the same criteria as described for the population sequencing-based frequency filter (as in step 3, above).

5. Sequence context: We excluded all CNVs with ≥30% coverage by somatic hypermutable sites, segmental duplications, simple/low-complexity/satellite repeats, or N-masked bases of the hg19 reference assembly. Coverage was computed using BEDTools, repeats were obtained from the UCSC Genome Browser, and a list of somatic hypermutable sites was repurposed from a previous study (Collins et al., 2020; Lee et al., 2019; Quinlan and Hall, 2010).

Finally, after applying the CNV harmonization procedures described above to the raw CNV calls from each cohort, we pooled harmonized CNV data across cohorts into four matched groups, dubbed "meta-cohorts," to control for technical differences between individual data sources and cohorts. These meta-cohorts represent the basic unit on which all association testing was performed. Individual cohorts were assigned to meta-cohorts on the basis of similarity between sample recruitment strategies, study or cohort design, microarray platforms, and processing pipelines for each callset. Meta-cohort assignments were as follows:

- *Cohort 1*: mainly aCGH-based CNVs from diagnostic laboratories, including BCH, Coe, Cooper, GDX, and IU.
- *Cohort 2*: mainly SNP array-based CNVs from case-control disease association studies, including Epi25k, PGC, SSC, SickKids, TCGA, TSAICG.
- *Cohort 3*: CHOP.
- *Cohort 4*: UKBB.

<u>Phenotype standardization</u>
Given the variability of phenotypic detail available between different cohorts, we applied a standardized hierarchical phenotype consolidation scheme uniformly across all samples to convert all sample-level phenotype labels into Human Phenotype Ontology (HPO) terms (Kohler et al., 2019). This process is described below.

First, for each sample, we recursively matched all available phenotype information against HPO keywords based on keyword substrings and assigned the corresponding HPO terms to each sample when a match was found. We abided by all HPO phenotype definitions with a single exception: we matched all congenital anomalies to HP:0001197 (Abnormality of prenatal development or birth), despite the description for that term excluding fetal structural anomalies. We made this exception because congenital anomalies were a phenotype of particular interest in this study, and there was no obvious existing HPO code corresponding to these phenotypes. The code to perform this HPO conversion is contained in “convert_phenotypes_to_hpo.sh”.

For the UKBB cohort, all phenotypes were encoded in the UKBB-sanctioned version of ICD-10. Given the scope of analyses in this study, we reduced the 19,153 unique ICD-10 codes to a smaller subset relevant to this study. This was accomplished by (1) automated filtering to isolate ICD-10 codes with a cohort prevalence of at least 0.01% but no greater than 5% using “prune_ukbb_icd10_dictionary.py”, and (2) manual review by a board-certified physician (P.M.B.) to further identify terms unlikely to have a strong genetic component. Afterwards, phenotypes for each sample were converted from ICD-10 to plain-text indications using “icd10_to_indication.py”. Once converted to plain-text indications, the UKBB cohort was subjected to indication-to-HPO conversion as described above.

Second, after assigning HPO terms to each sample, we next reduced the number of HPO terms used in this study to those with appreciable sample size and those that were partially non-overlapping with other related HPO terms. We tabulated the number of samples matching each HPO code and retained all HPO terms with ≥2,000 samples. Next, we compared shared sample memberships between all pairs of hierarchically related HPO terms to determine the optimal subset of non-redundant terms. If <2,000 samples differed between a pair of HPO terms, we retained the more general (*i.e.*, higher-level) term, and excluded the more specific (*i.e.*, lower-level) term. If both terms were equally high-level and siblings (defined as reciprocally sharing at least 50% of their parent terms), we retained the term with the larger sample size. Finally, to help control for batch-specific technical artifacts, we required that all HPO terms be represented with

≥500 samples in at least two different meta-cohorts, as defined above. This process yielded a condensed hierarchical phenotype classification system with 30 distinct HPO terms, each which had ≥2,000 samples in total, ≥500 samples from at least two different meta-cohorts and differed from all other HPO terms by ≥2,000 distinct samples. The final list of HPO terms and samples per term per cohort is provided in **Table S2** and **Figure S2**.

*Previously reported genomic disorders*
We curated lists of previously reported genomic disorders (GDs), defined as genomic intervals where rare CNVs have been linked to one or more diseases. For this purpose, we integrated lists of GDs from six existing publications and public resources (Dittwald et al., 2013; Firth et al., 2009; Girirajan et al., 2012; Owen et al., 2018; Riggs et al., 2012; Stefansson et al., 2014). We extracted GD coordinates from these references as provided and converted them to hg19 using UCSC liftOver where necessary (Lee et al., 2019). We only considered GDs from the ClinGen Pathogenic CNV Regions list that were scored at high- or medium-confidence. After overlapping all GDs from the six sources above, we trimmed overlapping segmental duplications overlapping the boundaries of each GD interval, if any were present. Segmental duplications were downloaded from the UCSC Genome Browser in hg19 coordinates (Lee et al., 2019).

*Genes and gene sets*
Many of the analyses in this study used consensus gene definitions, coordinates, and gene sets. Therefore, we curated a standard reference file of autosomal genes and gene sets. The commands executed to curate gene-based data are contained in “format_genes.sh” and are also described below.

For all analyses in this study, we defined genes strictly as canonical transcripts from autosomal protein-coding genes as defined by Gencode v19 (Harrow et al., 2012). We extracted canonical transcripts from for protein-coding genes using “get_canonical_transcripts.py”, which implements canonical transcript filters according to the official Ensembl definition. We further restricted all canonical transcripts to exons expressed in ≥20% of transcripts in at least one human tissue catalogued by the Genotype-Tissue Expression (GTEx) Project (Battle et al., 2017). To accomplish this, we computed the per-exon maximum proportion expressed across transcripts (“pext”) score across all available tissues (Cummings et al., 2020). Bases from exons with missing pext scores were ignored when calculating mean pext scores per exon, and exons entirely missing pext scores were considered as passing the ≥20% requirement. Genes with no exons remaining after exon-level expression filtering were removed outright from all subsequent analyses.

We also considered various subsets of genes throughout this study. A full list of these gene sets is provided in **Table S5**, along with inclusion criteria and references for external data where necessary. When curating these gene sets, we required unique matches to gene symbols from one of the canonical protein-coding genes curated above.

*Predicted NAHR-mediated CNV regions*
We defined a set of loci where NAHR-mediated CNVs might be predicted to occur based on the genomic properties of the flanking regions. To build this set of loci, we first defined pairs of segmental duplications meeting all of the following criteria:

1. Both members of the pair mapped to the same chromosome;
2. The pair was no closer than 100kb and no farther than 10Mb apart;
3. Both members of the pair were ≥1kb in size;
4. The pair featured strict sequence identity (no indels) ≥95%;

5. The pair mapped with direct orientation of repeats (*i.e.*, same strand);
6. The total intervening sequence between members of the pair had ≤30% coverage by the excluded regions used during CNV harmonization (as described above); and
7. At least 100kb of the intervening sequence remained after subtracting the excluded regions used during CNV harmonization.

After defining candidate pairs of segmental duplications (above), we collapsed overlapping pairs into predicted NAHR-mediated CNV regions while requiring ≥50% reciprocal overlap of intervening sequence per BEDTools as well as both ends of their respective intervals to be within 1Mb of each other (Quinlan and Hall, 2010). For each cluster of segmental duplication pairs, we retained the pair with the smallest intervening (*i.e.*, spanning) distance, and used the innermost coordinates for analysis purposes.

*Gene-level features*
We compiled a master table of gene-level features to be used in gene-based fine-mapping and genic dosage sensitivity scoring. The full list of features is provided in **Table S7**. For each of the canonical protein-coding genes curated for this study (see above), we collected a total of 129 gene-level features across four major categories: genomic, expression, chromatin, and mutational constraint. All expression features were derived from GTEx v7 due to it being the last GTEx version native to hg19, which was a direct match for the Gencode version used in this study (Battle et al., 2017; Harrow et al., 2012). All chromatin features were based on the Roadmap Epigenomics Project (REP) using the expanded 18-state ChromHMM model on 98 tissues as described in a previous publication (Kundaje et al., 2015).

Following annotation of all 129 features per gene, we collapsed correlated annotations to retain the principal components, or "eigenfeatures," that captured at least 99% of inter-gene variance. However, prior to principal components analysis (PCA), we normalized the data in two steps. First, all variables were transformed using Box-Cox power transformations across all genes, except for a small subset of features as noted in **Table S7**. Second, following transformation, all variables were centered (mean=0) and scaled (standard deviation=1) across all genes. We subsequently performed PCA on the transformed & standardized gene feature matrix and used the resulting principal components as features inputs to gene-based fine-mapping and dosage-scoring.

*Genome annotations*
We curated a large database of genome annotation tracks for association testing. This process is described below, but all commands required to curate the datasets referenced below are contained in "curate_annotations.sh". In practice, this curation process was parallelized using "curate_annotations.wdl" on the Google Cloud Platform via Terra, a scientific cloud computing web-based interface (Birger et al., 2017).

For the purposes of this study, we aimed to comprehensively assess all major repositories of genome annotations. In total, we evaluated 11,612 genome annotation tracks across all sources, which are listed in **Tables S9-10**. We subjected each annotation track to an identical set of curation steps, as follows:

1. Element size & ploidy: We excluded all elements not mapped to autosomes from the primary hg19 assembly and further required all elements to be ≥5bp and ≤200kb in size.

2. Sequence context: We excluded any elements covered at least 10% by the same set of excluded loci used during CNV curation (described above). Given the small size of many

annotations and the dozens of transcript isoforms for many T-cell receptor gene clusters, we merged all somatic hypermutable loci using a distance of ±100kb prior to track curation for the purposes of this analysis. Element coverage was assessed with BEDTools (Quinlan and Hall, 2010).

3. Nonredundancy: We collapsed all overlapping elements from the same annotation track using BEDTools (Quinlan and Hall, 2010).

After curation, we computed several summary statistics for each track, including total number of elements, minimum/mean/median/maximum element sizes, and total nonredundant nucleotides covered by all elements in the track. These statistics are provided in **Table S11**.

We included several thousand annotation tracks from ENCODE and REP, for which we applied additional criteria. To be considered in this analysis, ENCODE/REP tracks had to meet all of the following criteria: (1) available for download in BED format, (2) aligned to hg19, (3) sample from an unperturbed human cell type, tissue, or cell line, (4) no ENCODE data audit errors (color code: red) or non-compliances (color code: orange), (5) marked as "released" status (*i.e.*, not archived and/or retracted), and (6) must be peaks-only (no background sample).

#### 2 | Large segment association meta-analyses

We designed a sliding window framework to scan all autosomes for signals of association between rCNVs and any of the disease phenotypes considered in this study. To accomplish this, we meta-analyzed counts of rCNVs in cases vs. controls in parallel for each phenotype. This procedure is detailed below, but the code to execute these analyses is contained in “sliding_window_analysis.sh”. In practice, this analysis was parallelized on FireCloud/Terra using “sliding_window_analysis.wdl” (Birger et al., 2017).

*Sliding window model design*

We generated sliding windows for all autosomes at 200kb resolution with 10kb step size and excluded any windows with ≥30% coverage by N-masked sequences or known somatically hypermutable site, as applied in our CNV harmonization (described above). The window size of 200kb was selected to approximately match the median rCNV size for most cohorts after harmonization. After filtering, we retained a final set of 267,237 windows for analysis.

Next, we intersected rCNVs against all sliding windows separately for cases and controls for each cohort while requiring rCNVs to overlap at least 50% of each window to be counted. We conducted this procedure twice per phenotype group & cohort: once each for deletions and duplications. Given that rCNV breakpoint precision varies by locus, platform, and CNV calling algorithm, we systematically extended the breakpoints of each control rCNV by +50kb. rCNV breakpoints in cases were not extended. We did this to conservatively protect against spurious associations arising from situations where control rCNVs breakpoints might be underestimated compared to case breakpoints.

After CNVs were tallied in cases and controls for each window, we next compared the ratios of CNV carriers between cases and controls per phenotype per cohort using a one-sided Fisher's exact test. While we did not use these single-cohort association statistics for any assessment of significance, and instead used a fixed-effects meta-analysis (as described below), we did use the Fisher’s exact test results for designating the individually most significant cohort per window for each combination of CNV type and phenotype.

<u>*Meta-analysis association test*</u>
We combined rCNV association statistics across metacohorts for each sliding window using a fixed-effects meta-analysis implemented using the metafor R package (Viechtbauer, 2010). We applied two key modifications to this meta-analysis. First, given that rCNV counts per window are (a) sparse, (b) zero-inflated, and (c) had unbalanced case & control sample sizes for most phenotype groups (*e.g.*, frequently >10- to 100-fold more controls than cases), we implemented an empirical continuity correction as proposed in a previous publication (Sweeting et al., 2004). Second, as sample size imbalance between cases and controls has been shown to distort test statistics in genetic association studies, we applied a saddlepoint approximation correction to the test statistics for each phenotype per CNV type to recalibrate our P-values. Our implementation of this procedure was based on a previously proposed algorithm (Dey et al., 2017).

Each phenotype and CNV type were meta-analyzed separately for a total of two genome-wide meta-analyses per phenotype. Further, for each combination of phenotype and CNV type, we conducted two versions of the same meta-analysis: once while including all four cohorts, and once while conditionally excluding the single most significant cohort per window, as described above. We designated the results from the model including all four cohorts as the "primary" statistics and designated the results from the conditional exclusion model as the "secondary" statistics for purposes of defining genome-wide significance (described below).

<u>*Assessing genome-wide significance*</u>
After conducting genome-wide meta-analyses for each combination of phenotype and CNV type, we next wanted to control false discovery rate (FDR) at an approximate equivalent to genome-wide significance. However, estimating the number of independent tests performed across all windows, phenotypes, and CNV types is difficult due to numerous necessary assumptions, such as the independence of samples between phenotypes or the local correlation structure of CNV counts between neighboring windows. Instead, we used a "genome-wide" significance threshold of $P \leq 3.72 \times 10^{-6}$, which corresponds to a Bonferroni correction if applied to the number of non-overlapping 200kb windows tested in our analysis.

We empirically assessed the calibration of this primary P-value threshold using a permutation-based approach, which is summarized in **Note S1**. We repeated the following four steps 50 times for each CNV type and phenotype:

1. Permuted phenotype labels for all rCNVs while matching on size (split by quantile) and CNV type (deletion or duplication);

2. Reran full sliding window rCNV association tests, including meta-analysis, for each phenotype and CNV type combination;

3. Computed the fraction of significant windows for a broad range of primary P-value thresholds (*e.g.*, $-\log_{10}(P)$ ranging from 0-20); and

4. Reported the least significant primary P-value threshold that results in an empirical FDR $\leq 3.72 \times 10^{-6}$ (*i.e.*, zero false positive associations expected by chance).

Finally, we computed the median of the empirically observed P-value thresholds for each of the 50 permutations per CNV type (N=2) per phenotype (N=30) and compared these empirical P-value thresholds to our approximated Bonferroni-equivalent threshold. As is detailed in **Note S1** and **Figure S4**, we found that the Bonferroni approximation appeared to be appropriate for our datasets and study design.

In practice, we considered a window to be genome-wide significant if its primary P-value exceeded the Bonferroni-corrected genome-wide significance threshold, and it additionally satisfied at least one of the following two criteria: (1) its secondary P-value (as described above) was also nominally significant (P<0.05); and/or (2) at least two separate cohorts were nominally significant (P<0.05) per single-cohort Fisher's exact tests. Absent a true replication sample, these *post hoc* filters were required to protect against Winner's Curse (Tam et al., 2019).

*Association refinement and annotation*

For each locus with a significant association between rCNVs and one or more phenotypes, we aimed to identify the minimal interval(s) that contained the causal element(s) with 99% confidence. This procedure is described below and was performed separately for deletions and duplications using "refine_significant_regions.py".

First, per phenotype, all significant windows were collapsed into nonredundant blocks by merging all windows within ±200kb of any significant window. Next, for each block, we executed the following three steps:

1. Computed an approximate Bayes factor (ABF) for each window following the procedure specified by Wakefield *et al.* in prior publications (Wakefield, 2007, 2009);

2. Identified the minimal set of windows that captured at least 99% of the total ABF for the block (*i.e.*, defined the 99% credible set) by ranking all $n$ windows in descending order according to their ABF and subsequently computing the minimum set of $k$ windows as follows:

$$\text{99\% credible set} = \left\{ k \in n : \frac{\sum_{i=1}^{k} \Lambda_i}{\sum_{i=1}^{n} \Lambda_i} \geq 0.99 \right\}$$

   Where $\Lambda_i$ is the ABF per window $i$; and

3. Merged all windows in the 99% credible set (with ±200kb padding) to define the set of intervals comprising the 99% credible set.

The result of this process was a list of 99% credible sets for each genome-wide significant locus identified by our sliding window meta-analysis. After refinement, we computed two summary statistics per credible set:

1. Case and control rCNV frequencies, which were computed as the mean rCNV carrier rate across all windows in the 99% credible set; and

2. Pooled effect sizes, which were computed as the inverse variance-weighted mean across all windows in the 99% credible set.

After credible sets were defined for each association, we collapsed associations across phenotypes to derive a final set of nonredundant disease-associated rCNV segments for downstream analyses. To reduce associations to nonredundant rCNV segments, we clustered any overlapping credible intervals of the same CNV type (deletion or duplication) from different phenotypes. After clustering, we computed two summary statistics for each rCNV segment:

1. Case and control rCNV frequencies, which were computed as the weighted mean rCNV carrier rate per 99% credible set, where the weights corresponded to the square root of the sample size for each phenotype; and

2. Pooled effect sizes, which were computed as the inverse variance-weighted mean across all significant phenotypes.

Following our sliding window meta-analysis and association refinement, we annotated each genome-wide significant rCNV segment based on overlap with exons from any protein-coding gene, any overlap with previously reported GDs, and ≥25% reciprocal overlap by size with any predicted NAHR-mediated CNV interval. We manually polished all *in silico* NAHR mechanism predictions by comparing the segment coordinates to the hg19 segmental duplication, repeat masker, and assembly gap tracks in the UCSC Genome Browser (Lee et al., 2019) and revised any apparent discrepancies with our computational annotations.

*Feature enrichment tests*

We assessed disease-associated rCNV segments for enrichments of various genomic and gene-level features using two different permutation-based approaches. First, for features based on genomic coordinate (*e.g.*, previously reported GDs or all genes), we conducted 100,000 random permutations while matching on rCNV segment size using a custom implementation of BEDTools shuffle (Quinlan and Hall, 2010). Second, for all gene-based features (*e.g.*, proportion of constrained genes or number of disease genes), we conducted 100,000 random permutations while matching on number of genes per rCNV segment. In this gene-matched permutation approach, we randomly seeded each permuted segment by selecting one gene at random from anywhere in the genome, and iteratively added genes based on minimal linear distance (*i.e.*, closest genes first) until the total number of genes was matched to the number overlapped by the original rCNV segment. In this way, we generated randomly permuted lists of rCNV segments matched exactly on the number of genes per segment. In both the size-matched and gene-matched permutation schemes, we applied the same two restrictions: (1) we did not allow shuffled segments of the same CNV type to overlap, and (2) we did not consider any regions not covered by at least one 200kb window assessed in our sliding window meta-analyses.

For most features, we assessed enrichment by computing the average value across all rCNV segments for all 100,000 permutations and comparing the distribution of permuted values to the empirically observed average value of the rCNV segments from our disease-association meta-analyses. The exception was for all features relating to *de novo* loss-of-function and missense point mutations, where we computed the excess of mutations per segment by subtracting the expected number of mutations per segment extrapolated based on gene-specific mutation rates (Karczewski et al., 2020), and further normalized excess *de novo* damaging mutations against *de novo* synonymous mutations by residualizing against an outlier-robust linear fit of excess of damaging *de novo* mutations predicted from the excess of synonymous *de novo* mutations. This step was necessary to control for infrequent instances where we observed regional miscalibration of gene-based mutation rates.

#### 3 | Gene-based association meta-analyses

We designed a gene-based meta-analysis model to identify individual genes where rCNVs were associated with one or more diseases. This procedure is detailed below, but the code to execute these analyses is contained in “gene_burden_analysis.sh”. In practice, this analysis was parallelized on FireCloud/Terra using “gene_burden_analysis.wdl” (Birger et al., 2017).

*Gene-based model design*
Our gene-based association test compared counts of gene-overlapping rCNVs in cases vs. controls per cohort across all 30 phenotypes considered in this study. We begin by enumerating all autosomal protein-coding genes that passed our curation procedure as described above. We further excluded genes if their canonical transcript was ≥30% covered by any of the excluded regions during CNV harmonization (*e.g.*, somatically hypermutable sites, simple repeats, etc.). After all filtering, we retained 170,422 exons from 17,263 genes for these analyses.

Having curated a set of genes and exons to test, we next tallied genic rCNVs in cases vs. controls for each CNV type, cohort, and phenotype. For each rCNV-gene pair, we computed the total fraction of exonic bases (CDS) from the gene overlapped by the rCNV and considered a rCNV to overlap a gene based on its CDS overlap conditional on its CNV type. For deletions, we required ≥10% CDS overlap, since we reasoned that most coding deletions should result in protein truncation but we wanted to impose a liberal minimum overlap to protect against spurious annotations given the coarse resolution of most microarrays. For duplications, we instead required ≥75% CDS overlap. Given that functional annotation of duplications is more challenging than deletions, we imposed a stricter CDS overlap to isolate rCNVs predicted to duplicate nearly all of a gene.

Finally, as in our sliding window analyses, we computed per-cohort association statistics with one-sided Fisher's exact tests for each gene, CNV type, and phenotype, but only used these statistics for determining the individually most significant cohort per gene for each combination of CNV type and phenotype.

*Meta-analysis association test*
We conducted exome-wide disease-association meta-analyses for each phenotype and CNV type using identical methods as for our sliding window analyses (described above), namely a fixed-effects model with empirical continuity correction and saddlepoint approximation to update the null distribution used for significance testing (Dey et al., 2017; Sweeting et al., 2004).

*Assessing exome-wide significance*
We assessed our gene-based rCNV association tests at a Bonferroni-equivalent of exome-wide significance of $P \leq 2.90 \times 10^{-6}$ (*i.e.*, adjusting for 17,263 genes tested). We assessed the calibration of this exome-wide significance threshold identically to our sliding window analyses by performing 50 random phenotype label-swapping permutations for each CNV type and phenotype. Similar to our sliding window analyses, we found that the exome-wide P-value threshold was a good match for the expectation derived by permutations (data not shown, but qualitatively similar to **Figure S4**).

In practice, we considered a gene to be exome-wide significant if its primary P-value exceeded the Bonferroni-corrected exome-wide significance threshold, and it additionally satisfied at least one of the following two criteria: (1) its secondary P-value (as described above) was also nominally significant ($P<0.05$); and/or (2) at least two separate cohorts were nominally significant ($P<0.05$) per single-cohort Fisher's exact tests.

*Fine-mapping genic associations*
Lastly, we aimed to prioritize individual likely causal genes, and to define the minimal set of genes per block that most confidently explains each phenotypic association. To accomplish this, we first clustered all significant genes per phenotype into "significant gene blocks" by grouping all genes within ±1Mb of an exome-wide significant gene per phenotype. We next adapted established fine-

mapping algorithms from conventional genome-wide association studies (GWAS) to our rCNV-based data, as follows:

1. We transformed all association statistics for all genes per block—including non-significant genes—into approximate Bayes factors (ABFs) following the same procedure as in our sliding window analysis (Wakefield, 2007, 2009). For these calculations, we used an empirical Bayes approach to estimate null variance based on the observed variances for (i) all exome-wide significant genes pooled across all phenotypes, (ii) the most significant gene per gene block, and (iii) four manually curated lists of genes with known or likely haploinsufficiency. We applied Bayesian model averaging across these different null variance estimates.

2. We calculated posterior inclusion probabilities (PIPs) for each gene per block based on the ABFs from [1] while assuming a flat prior (*i.e.*, when assuming that each gene per block is equally likely to be causal).

3. We developed an adaptation of E-M algorithms described in two previous studies (Kichaev et al., 2017; Wen et al., 2017) to iteratively update gene priors for each block based on functional enrichments in a logistic generalized linear model (GLM). For this purpose, we used the gene-level principal components derived from all 129 gene features described earlier, applied light L2 regularization with a penalty of 0.1, and assessed convergence based on the square root of the mean squared error of both PIPs and GLM coefficients $\leq 10^{-8}$. In each iteration, we averaged PIPs for individual genes associated with multiple phenotypes where applicable.

4. Finally, we computed a 95% credible set of genes per block based on the cumulative sum of PIPs for genes ranked by causal likelihood. We only retained exome-wide significant genes after calculating each credible set (*i.e.*, genes that were not originally exome-wide significant were included when calculating the 95% credible set but were removed from the credible set after calculation).

For downstream analyses, we considered any gene with PIP ≥ 0.15 to be a "confident" candidate gene, and any gene with PIP ≥ 0.85 to be a "very confident" candidate gene.

#### 4 | Noncoding association meta-analyses

We developed a three-stage procedure for genome-wide association meta-analyses between noncoding rCNVs overlapping predicted *cis*-regulatory elements and the disease phenotypes considered in this study. This process is described below, but is also provided in “noncoding_association_analysis.sh,” and in practice was parallelized on FireCloud/Terra using “noncoding_association_analysis.wdl” (Birger et al., 2017).

*Noncoding rCNV filtering*

For the purposes of association testing, we defined “noncoding” rCNVs as those not predicted to disrupt any coding sequence that might credibly influence disease risk via strong-effect genetic variation, as explained in **Note S2**. Specifically, we retained all rCNVs that did not overlap any annotated exon from any canonical protein-coding gene unless that gene was one of 4,793 genes predicted to be mutationally tolerant in the general population. Prior to rCNV exclusion, we padded all exons from all genes by ±50kb to protect against CNV breakpoint imprecision.

*Prioritization of genome annotation classes for association testing*

To reduce our search space for association testing, we next assessed which of the 11,612 genome annotation classes we curated previously (see above) had evidence of dosage sensitivity in disease. For each track, we counted the number of noncoding rCNVs that completely overlapped at least one element per class, split by cohort, CNV type, and phenotype. After tabulating counts of CNVs per class, we conducted an inverse-variance weighted Z-score meta-analysis for each class with saddlepoint approximation applied across all classes per cohort. We considered any class with $P<0.05$ to have some evidence for possible disease relevance to be included for subsequent association testing.

*Definition of* cis-*regulatory blocks (CRBs)*
Given that many genome annotations are correlated, we next clustered elements into CRBs for association testing. After identifying which genome annotation classes had a global burden of noncoding rCNVs in cases, we clustered all elements across these classes using the density-based DBSCAN algorithm with a ±5kb neighborhood distance, a minimum number of elements per cluster proportional to 5% of the total number of annotation tracks being considered (rounding down), and a minimum number of tracks with at least one element per cluster proportional to 1% of the total number of tracks being considered (rounding down). After clustering, CRBs were assigned the minimum start and maximum end coordinate across all their constituent elements. Finally, pairs of non-overlapping CRBs within ±10kb were merged, and we excluded CRBs meeting any of the following four criteria: (1) >500kb in size; (2) ≥30% covered by the exclusion regions used during rCNV harmonization (see above); (3) within ±100kb of an element from the CNV harmonization exclusion regions that was ≥100kb in size; or (4) did not overlap a hypothetically testable interval (*i.e.*, any contiguous 100kb interval with no protein-coding exons excluded during noncoding rCNV filtering). In total, this procedure generated 15,497 non-overlapping CRBs for association testing.

*CRB-based model design*
Similar to our previous sliding window and gene-based association tests, our noncoding association test compared counts of noncoding rCNVs per CRB in cases vs. controls per phenotype. For each cohort, we tallied noncoding rCNVs against CRBs separately for cases and controls. Specifically, we intersected noncoding rCNVs versus the individual elements from each CRB, and only counted rCNV-element pairs where the rCNV completely (100%) covered the element. After intersecting rCNVs with individual elements, we tallied all rCNV-CRB pairs where the rCNV hit at least 50% of all elements from that CRB. We conducted this procedure a total of two times per phenotype group & metacohort, once each for deletions and duplications. Finally, we computed per-cohort association statistics with one-sided Fisher's exact tests for each CRB, CNV type, and phenotype, but only used these statistics for determining the individually most significant cohort per CRB for each combination of CNV type and phenotype.

*Meta-analysis association test*
We conducted genome-wide disease-association meta-analyses per CRB for each phenotype and CNV type using identical methods as for our sliding window analyses (described above): a fixed-effects model with empirical continuity correction and saddlepoint approximation to update the null distribution used for inference (Dey et al., 2017; Sweeting et al., 2004). Finally, for each CRB, we re-ran an identical meta-analysis while considering all rCNVs (*i.e.*, not restricted to noncoding rCNVs only), and used these statistics during quality control, as described below.

*Assessing genome-wide significance*
We assessed our noncoding rCNV association tests at a Bonferroni-equivalent of genome-wide significance of $P \leq 3.23 \times 10^{-6}$ (*i.e.*, adjusting for 15,497 CRBs tested). We evaluated the calibration of this genome-wide significance threshold identically to our sliding window and gene-based analyses by performing 50 random phenotype label-swapping permutations for each CNV type and phenotype. Like with our previous analyses, we found that this approximate genome-wide P-

value threshold was a good match for the expectation derived by permutations (data not shown, but qualitatively similar to **Figure S4**).

In practice, we considered a CRB to be genome-wide significant if its primary P-value exceeded the Bonferroni-corrected significance threshold for both the meta-analysis restricted to noncoding rCNVs and the meta-analysis when including all rCNVs (*i.e.*, including all gene-disruptive rCNVs), and it additionally satisfied at least one of the following two criteria: (1) its secondary P-value (as described above) was also nominally significant (P<0.05); and/or (2) at least two separate cohorts were nominally significant (P<0.05) per single-cohort Fisher's exact tests.

#### 5 | Gene dosage sensitivity scoring

We developed a model to predict haploinsufficiency and triplosensitivity for each protein-coding gene across all 22 autosomes, which is detailed below. The code to reproduce these analyses is contained in "gene_scoring_analysis.sh". In practice, this analysis was parallelized on FireCloud/Terra using "gene_scoring_analysis.wdl" (Birger et al., 2017).

*Curating training gene sets*

We first curated four sets of genes likely to be dosage sensitive based on existing evidence and used these gene sets for estimating priors and training our model. Starting from the list of canonical protein-coding autosomal genes we curated for this study (described above), we defined four training gene sets as follows:

- *Haploinsufficient genes*: all genes meeting at least two of the following three criteria: (1) constrained against protein-truncating mutations in gnomAD v2.1 (Karczewski et al., 2020), (2) confirmed autosomal dominant causes of disease via loss-of-function or haploinsufficient mechanisms per ClinGen (“high confidence” only) or DECIPHER/DDG2P (“confirmed” only), or (3) intolerant against low-expressor samples in GTEx v7 (Battle et al., 2017). Details for each of these criteria are provided in **Table S5**.

- *Triplosensitive genes*: all genes meeting at least two of the following three criteria: (1) constrained against missense mutations in gnomAD v2.1 (Karczewski et al., 2020), (2) confirmed autosomal dominant causes of disease via triplosensitive, gain-of-function, or other unspecified mechanisms per ClinGen (triplosensitive genes of any confidence rating) or DECIPHER/DDG2P (“confirmed” gain-of-function or “other” mechanism only) (Rehm et al., 2015; Wright et al., 2015), or (3) intolerant against high-expressor samples in GTEx v7 (Battle et al., 2017). Details for each of these criteria are provided in **Table S5**.

- *Haplosufficient genes*: all genes meeting at least two of the following three criteria: (1) mutationally tolerant in gnomAD v2.1 (Karczewski et al., 2020), (2) no reported disease associations per OMIM, ClinGen, or DECIPHER/DDG2P (Amberger et al., 2015; Rehm et al., 2015; Wright et al., 2015), or (3) high rate of low-expressor outlier samples in GTEx v7. Details for each of these criteria are provided in **Table S5**.

- *Triploinsensitive genes*: all genes meeting at least two of the following three criteria: (1) mutationally tolerant in gnomAD v2.1 (Karczewski et al., 2020), (2) no reported disease associations per OMIM, ClinGen, or DECIPHER/DDG2P (Amberger et al., 2015; Rehm et al., 2015; Wright et al., 2015), or (3) high rate of high-expressor outlier samples in GTEx v7. Details for each of these criteria are provided in **Table S5**.

<u>*Parameter optimization*</u>
We optimized four parameters used in model training. This process is described in **Note S3**, with additional technical details provided below:

- *CDS overlap per gene:* the goal of our model was to predict the consequences of whole-gene loss or gain (*i.e.*, 100% of CDS deleted or duplicated). However, given the possibilities of CNV breakpoint imprecision, transcript misannotation, and other technical factors, we required ≥80% CDS overlap for a CNV to count as gene-disruptive for both deletions and duplications.

- *Maximum number of genes per CNV:* given that our dosage sensitivity model aimed to quantify the effects of CNVs on individual genes, we assigned a limit on the maximum number of genes a CNV can disrupt in order to be considered by our model. To determine the optimal threshold of genes per CNV, we maximized the difference in "signal-to-noise" ratio between cases & controls by comparing the rate of CNVs disrupting of curated likely dosage sensitive and insensitive genes. First, we computed the sum of observed disruptions per gene set separately in cases and controls per cohort for CNVs impacting no more than N genes, where N was sequentially incremented from zero to 80. Second, we calculated the signal-to-noise ratio as the quotient of the number of sensitive vs. all sensitive and insensitive disruptions. Third, we computed the numeric difference of signal-to-noise ratios between cases and controls per cohort. Fourth, we conducted a weighted maximization of these signal-to-noise ratio differences across cohorts, which was weighted by the total number of positive disruptions per cohort. These analyses determined that the optimal threshold was ≤24 genes per CNV to maximize the signal-to-noise ratio between cases and controls for subsequent modeling.

- *Phenotypes:* to determine which phenotypes should be included in training, we computed the case-control odds ratio per cohort for whole-gene CNVs that overlapped no more than 24 genes in total and disrupted either likely dosage sensitive versus insensitive genes. After computing these odds ratios per cohort, we combined them using the same fixed-effects meta-analysis approach from our genome-wide association analyses. We retained all phenotypes below the highest level HPO term (*i.e.*, all cases) that were nominally significant for both (i) deletions of curated likely haploinsufficient genes and (ii) duplications of curated likely triplosensitive genes. This process prioritized nine phenotypes to be used in subsequent model training.

- *Minimum number of CNVs per gene:* lastly, we wanted to exclude genes from model training that had insufficient CNV evidence to contribute accurate effect size estimates. To accomplish this, we first computed odds ratios for deletions and duplications of each gene with a fixed-effects meta-analysis using the optimized parameters as described above. From these effect size estimates for all genes, we determined the minimum number of CNVs per gene (≥10) such that the remaining genes passing this threshold had a mean effect size standard error less than one. Note that this threshold was applied to the total number of CNVs observed between cases and controls across all cohorts.

<u>*Definition of training exclusion regions*</u>
We only considered genes that could be accurately modeled when estimating priors and when training the dosage sensitivity classifier. Specifically, we excluded genes meeting any of the following three criteria: (1) genes overlapping any of the 114 known GDs we curated from the literature for previous analyses, (2) genes within ±1Mb of any annotated centromere or telomere, and (3) genes with <10 deletions or duplications meeting all criteria for this analysis (described

above). However, when training the dosage sensitivity classifier, we re-introduced any genes from our curated positive or negative training gene sets, even if they had insufficient CNV evidence, were near a centromere or telomere, or overlapped a known GD locus. Thus, after applying all of these criteria, we retained a total of 5,399 and 5,475 genes for training our deletion and duplication models, respectively.

*Empirical Bayes estimation of priors*
Our model required estimating three priors: (1-2) the CNV effect sizes expected for dosage sensitive and insensitive genes, and (3) the expected fraction of all genes in the genome that are truly dosage sensitive. To estimate the expected effect sizes, we computed the median log-odds ratio across all genes in each training gene set of likely dosage sensitive or insensitive genes. To estimate the expected fraction of truly dosage sensitive genes, we averaged across four statistics derived from existing data: (i) 16.3% of all genes that are known to be constrained against protein-truncating mutations in gnomAD v2.1(Karczewski et al., 2020); (ii) 7.5% of genes from our curated deletion training sets were likely haploinsufficient; (iii) 20.5% of all genes have at least one disease association reported in OMIM (Amberger et al., 2015); (iv) 3.9% of all genes have some evidence for dominant haploinsufficiency per ClinGen and/or DDG2P, when including genes with unknown or unconfirmed mechanisms (Bragin et al., 2014; Rehm et al., 2015). Averaging across all four of these values produced a prior expectation of 12.0% of all genes being truly dosage sensitive.

*Bayesian dosage sensitivity likelihoods*
We next computed the likelihood ratio that each gene was dosage sensitive versus insensitive based on the CNV effect size empirically observed in our dataset. For each gene, $g$, with observed log-odds ratio $\hat{\theta}_g$ and standard error $\hat{V}_g$, we specified null and alternative hypotheses as follows:

$$\begin{cases} H_0 : \hat{\theta}_g \leq \theta_0 \\ H_1 : \hat{\theta}_g \geq \theta_1 \end{cases}$$

Where $\theta_0$ and $\theta_1$ are the effect size priors for dosage insensitive and sensitive genes, respectively, computed on a CNV type-specific basis as detailed above. From these hypotheses, we computed the Bayes factor per gene as:

$$BF_g = \frac{\mathrm{P}\,(\hat{\theta}_g \mid H_0)}{\mathrm{P}\,(\hat{\theta}_g \mid H_1)} = \frac{1 - N(\hat{\theta}_g - \theta_0, 1)}{N(\hat{\theta}_g - \theta_1, \hat{V}_g)}$$

Where $N(m, v)$ denotes the normal distribution and we assumed unit variance ($v$) for $H_0$. Subsequently, we computed the Bayesian false discovery probability (BFDP) per gene based on equation (2) from (Wakefield, 2007) as:

$$BFDP_g = \mathrm{P}\,(H_0 | \hat{\theta}_g) = \frac{PO \times BF_g}{PO \times BF_g + 1}$$

Where the prior odds ($PO$) of each gene being dosage insensitive was:

$$PO = \frac{\pi_0}{1-\pi_0}$$

Where $\pi_0 = 1 - \pi_1 = 0.88$, given the prior of $\pi_1 = 0.12$ as estimated above.

Finally, we manually reassigned BFDP values of 0 or 1 to any gene in our curated training sets of likely dosage sensitive or insensitive gene, respectively.

<u>*Ensemble machine learning strategy & composite architectures*</u>
Using the BFDP values calculated for each gene described above, we next trained and applied an ensemble machine learning framework to predict the probability of a gene being haploinsufficient or triplosensitive based on 129 gene-level features. This process is described below and was conducted independently for deletions to predict haploinsufficiency and for duplications to predict triplosensitivity.

We first partitioned all genes into 11 subsets. Given that many gene-level features and our CNV evidence exhibited autocorrelation among neighboring genes, we did not randomly assign genes into these 11 subsets. Instead, we split all chromosomes by arm (*i.e.*, p or q), and grouped chromosome arms into 11 subsets while balancing the average number of genes per subset. Each of these 11 subsets therefore had roughly the same number of genes but the data for each subset was almost entirely independent from the other 10 subsets, as no rCNVs in our dataset were allowed to span centromeres dividing p and q arms.

We next used cross-validation (CV) to predict dosage sensitivity for all genes per subset. Specifically, for each of the 11 subsets, we used the remaining 10 subsets to train and cross-validate seven different models (described below) by sampling a random 90% of genes from these 10 subsets for training and using the remaining 10% of genes for testing. In each model training step, we attempted to predict per-gene BFDPs from the principal components of the 129 gene-level features as described earlier, and rounded BFDPs to binary values as needed based on each model's architecture. Genes were excluded from training based on the three criteria specified above. Following CV, we selected the best-fit model across all CVs based on the square root of mean squared error of predicted and actual BFDPs for the 10% of genes held out for testing and applied the best-fit model to predict BFDPs for all genes in the 11$^{th}$ subset, which was never included in any CV step. We predicted scores for all genes irrespective of training exclusion criteria. Importantly, this framework ensured that the data from each subset are never used to fit the model used to predict BFDPs for the genes in that subset. As a final step, we computed standardized scores for each gene as the transformation of predicted BFDPs based on the normal cumulative distribution function of predicted BFDPs across all 11 subsets, which scaled all scores to adhere to the range of [0, 1].

We applied this CV framework identically for seven different regression and machine learning models: two logistic GLMs (one with and one without stochastic gradient descent), a linear support-vector machine, a random forest, a latent discriminant analysis, a naïve Bayes classifier, and a three-layer feed-forward neural net with logistic activation. All models were implemented in Python using the SciKitLearn and StatsModels libraries (Pedregosa et al., 2011; Seabold and Perktold, 2010). After the seven models had been trained and applied to predict scores for all genes, we computed the receiver-operating characteristic (ROC) for each model versus their corresponding dosage sensitive and insensitive training genes and determined the ROC-optimal score cutoff as the minimized Euclidean distance between the ROC and hypothetically perfect performance (*i.e.*, 100% true positives and 0% false positives). Finally, we computed an ensemble score for each gene as the average of all seven models weighted by their accuracies corresponding to their model-specific ROC-optimal classification thresholds. We used these final ensemble scores for all subsequent analyses.

To evaluate model performance, we computed ROCs and precision-recall curves (PRCs) for all eight models, including the seven independent architectures and the final ensemble model. We used our training gene sets to define performance for both ROC and PRC analysis, and derived an overall rank of architecture performance based on the harmonic mean of the area under ROCs and PRCs for both deletions and duplications. As expected (Dietterich, 2000), the ensemble model outperformed each of the individual component models, justifying our selection of the ensemble model as a final architecture. To further evaluate the specificity of our deletion- and duplication-derived scores for predicting haploinsufficiency and triplosensitivity, respectively, we defined subsets of genes from our curated dosage sensitive gene sets that were only present in one of the two sets (*e.g.*, curated haploinsufficient training genes that were not also present in the curated triplosensitive training gene set) and subsequently repeated ROC and PRC analyses as described above.

*Score comparisons to* de novo *CNVs in autism families*
We assessed the utility of our pHI and pTS scores in interpreting the disease risk contributed by *de novo* CNVs (dnCNVs) from exome sequencing in 13,192 children affected with autism and their 5,011 unaffected siblings as described in **Note S4**. From this dataset, we conducted two complementary analyses after converting CNV coordinates from hg38 to hg19 using UCSC liftOver (Lee et al., 2019) and subsequently excluding dnCNVs with at least 50% reciprocal overlap versus any of the known GDs curated from the literature. In the first analysis, we scored all dnCNVs based on the highest pHI or pTS score of any gene with a predicted gene-truncating or whole-gene duplication per deletion and duplication, respectively. After scoring each dnCNV with pHI or pTS, we next stratified all dnCNVs into quartiles by score per CNV class and computed the empirical odds ratio per quartile of carrying a qualifying dnCNV between affected and unaffected children. In the second analysis, we labeled all affected and unaffected children carrying at least one dnCNV based on the highest pHI (for deletions) or pTS (for duplications) score across all of their dnCNVs and computed the ROC for classifying affected children purely based on the top score of any gene involved in a dnCNV.

*Score comparisons to gnomAD-SV*
We compared our pHI and pTS scores to the rates of gene-disrupting CNVs in the general population as documented by large-scale genome sequencing. To accomplish this, we counted the number of gene-truncating deletions or whole-gene copy-gain duplications from the subset of 8,342 unrelated samples with no known neurological phenotypes from the publicly available gnomAD-SV v2.1 dataset (Collins et al., 2020). We next separated genes into 10 fixed-width bins across the range of pHI and pTS scores in increments of ±0.1. For each bin of genes, we computed the average number of protein-truncating deletions or copy-gain duplications. We also computed a normalized value adjusting for 21 gene-level genomic features, like gene length and CDS length (see **Table S7** for details). To compute an expected number of CNVs in gnomAD-SV per gene, we first normalized and decomposed all 21 genomic features into their top 15 principal components as described earlier. We next fit a negative binomial regression model to predict the number of either protein-truncating deletions or copy-gain duplications in gnomAD based on these 15 eigenfeatures. To reduce the confounding effects of negative selection for constrained genes, we restricted to the subset of 5,578 likely unconstrained genes where we did not expect strong negative selection against coding variants (see **Table S5** for details) before fitting the model. We applied the fit model to predict an expected number of CNVs for all genes and computed a binwise observed:expected ratio by summing the total number of observed CNVs across all genes per pHI or pTS bin and dividing by the sum of expected CNV counts.

*Elastic net regression to isolate predictive features*
We identified gene-level features most predictive of dosage sensitive genes using two different elastic net regressions of all 129 gene-level features curated in this study versus combinations of

our pHI and pTS scores. In both regressions, all features were standard-normalized (*i.e.*, zero mean and unit variance) and we used 10-fold CV to optimize alpha and lambda hyperparameters for each elastic net. In the first analysis, we regressed the minimum of pHI and pTS per gene against all gene-level features. In the second analysis, we regressed the numerical difference between pTS and pHI against all gene level features. In both analyses, we reported standardized coefficients from the overall model as a measure of feature importance.
